# Prenatal smoking, alcohol and caffeine exposure and maternal reported ADHD symptoms in childhood: triangulation of evidence using negative control and polygenic risk score analyses

**DOI:** 10.1101/2021.03.25.21254087

**Authors:** Elis Haan, Hannah M. Sallis, Luisa Zuccolo, Jeremy Labrecque, Eivind Ystrom, Ted Reichborn-Kjennerud, Ole Andreassen, Alexandra Havdahl, Marcus R. Munafò

## Abstract

**Background and aims:** Studies have indicated that maternal prenatal substance use may be associated with offspring attention deficit hyperactivity disorder (ADHD) via intrauterine effects. We measured associations between prenatal smoking, alcohol and caffeine consumption with childhood ADHD symptoms accounting for shared familial factors.

**Design:** First, we used a negative control design comparing maternal and paternal substance use. Three models were used for negative control analyses: unadjusted (without confounders), adjusted (including confounders) and mutually adjusted (including confounders and partner’s substance use). The results were meta-analysed across the cohorts. Second, we used polygenic risk scores (PRS) as proxies for exposures. Maternal PRS for smoking, alcohol and coffee consumption were regressed against ADHD symptoms. We triangulated the results across the two approaches to infer causality. Setting: We used data from three longitudinal pregnancy cohorts: Avon Longitudinal Study of Parents and Children (ALSPAC) in the UK, Generation R study (GenR) in the Netherlands and Norwegian Mother, Father and Child Cohort study (MoBa) in Norway.

**Participants:** Phenotype data available for children was: N_ALSPAC_=5,455-7,751; N_GENR_=1,537-3,119; N_MOBA_=28,053-42,206. Genotype data available for mothers was: N_ALSPAC_=7,074; N_MOBA_=14,583. Measurements: A measure of offspring ADHD symptoms at age 7-8 years was derived by dichotomising scores from questionnaires and parental self-reported prenatal substance use was measured at the 2^nd^ pregnancy trimester.

**Findings:** The pooled estimate for maternal prenatal substance use showed an association with total ADHD symptoms (odds ratio (OR)_SMOKING_=1.11, 95% confidence interval (CI) 1.00-1.23; OR_ALCOHOL_=1.27, 95%CI 1.08-1.49; OR_CAFFEINE_=1.05, 95%CI 1.00-1.11), while not for fathers (OR_SMOKING_=1.03, 95%CI 0.95-1.13; OR_ALCOHOL_=0.83, 95%CI 0.47-1.48; OR_CAFFEINE_=1.02, 95%CI 0.97-1.07). However, maternal associations did not persist in sensitivity analyses (substance use before pregnancy, adjustment for maternal ADHD symptoms in MoBa). The PRS analyses were inconclusive for an association in ALSPAC or MoBa.

**Conclusions:** There appears to be no causal intrauterine effect of maternal prenatal substance use on offspring attention-deficit hyperactivity disorder symptoms.

## Introduction

Many observational studies have shown that symptoms and diagnosis of attention deficit hyperactivity disorder (ADHD) are associated with maternal prenatal smoking (1, 2) and mixed findings have been reported for association with prenatal alcohol and caffeine exposure (3-8). However, inferring causality from associations between maternal prenatal substance use and offspring ADHD is challenging because the association could be affected by unmeasured shared familial factors that contribute to both maternal prenatal substance use and offspring ADHD. Several studies have shown genetic overlap between substance use and ADHD (9), and maternal genetic risk for ADHD has been associated with smoking during pregnancy (10).

Negative control designs (i.e., parental comparison) have been used to investigate potential causal intrauterine effects for a range of outcomes (11, 12). The main principle of the negative control approach is to compare the association of interest with another related association which is not biologically plausible (11). For example, in a parental comparison, if the maternal exposure-child outcome association is stronger, compared with the paternal exposure-child outcome association, this would suggest a potentially causal intrauterine effect. In contrast, if the magnitude of association is similar, this would argue against a causal intrauterine effect, and instead suggest the association is due to confounding.

Negative control designs have been used in the context of maternal prenatal substance use and offspring ADHD. A study based on the Danish National Birth Cohort using parental comparison found evidence for a potential causal effect between maternal prenatal smoking and offspring ADHD diagnosis and symptoms(13). However, several other studies using negative control and other genetically sensitive designs have concluded that the association between maternal prenatal smoking and offspring ADHD is likely not causal (14, 15). Sibling comparison studies on alcohol exposure based on the Norwegian Mother, Father and Child Cohort Study (MoBa) have found little evidence for a causal effect with ADHD diagnosis (16, 17) although a sibling control analysis (16) suggested some evidence for a potential causal relationship with ADHD symptoms as measured by the Conner’s Parent Rating Scale (CPRS-R). To our knowledge no negative control studies have been published on prenatal caffeine exposure and offspring ADHD.

Although published negative control studies investigating intrauterine effects have improved our knowledge of causal effects, they may still be biased because of unmeasured and residual confounding. Using genetic variants is an alternative approach that can strengthen causal inference when using observational data. Genetic variants are randomly and independently assigned at conception and should therefore not be associated with factors that normally confound the exposure-outcome relationship. They can therefore provide stronger support for a potential causal effect (18). We used polygenic risk score (PRS) analyses to look at whether a potential causal relationship exists, based on the principles of Mendelian Randomization (MR). Here we test whether a causal effect exists, rather than estimating the size of the causal effect in a formal MR framework. Studies using genetic variants (PRS) as proxy for exposures rely on three main MR assumptions: (1) relevance – the genetic variant must be robustly associated with the exposure of interest; (2) independence – the genetic variant is not confounded with the outcome or related through selection bias and (3) exclusion restriction – the genetic variant is not associated with the outcome by any other path than through the exposure of interest (19). Assumptions 2 and 3 cannot be tested and, therefore, problems with horizontal pleiotropy – where the same genetic variant is directly associated with many phenotypes – confounding of genetic variant’s relationship with the outcome or selection bias cannot be ruled out (18).

Combining multiple methodological approaches that rely on different assumptions and are subject to different sources of bias – known as triangulation – can strengthen causal inference (20). If results from multiple approaches provide convergent results, it is more likely that the observed association reflects a causal effect (21). In the present study we combined the conventional multivariable regression approach, a negative control design using paternal prenatal substance use as a negative control for the intrauterine exposure, and genetic analyses using PRS as a proxy for the exposures of interest. Our aim was to investigate whether there is a causal effect of maternal prenatal substance use on offspring ADHD outcomes at age 7-8 (Figure 1), using data from three large prospective birth cohorts. Considering that some studies have found that maternal prenatal substance use can have a distinct effect on ADHD hyperactivity and inattention symptom domains (22, 23), we were interested in examining whether we observe similar findings.

**Figure 1.**
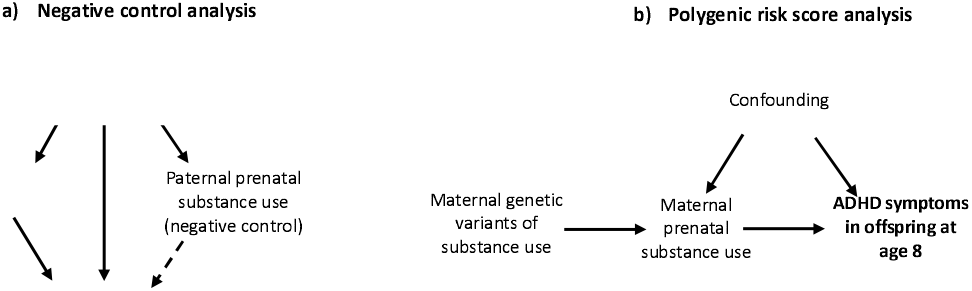
Study design. Note: a) The dashed arrow represents the negative control analysis. Assumption includes: the same confounders influence maternal and paternal prenatal substance use and offspring ADHD symptoms, a causal prenatal (intrauterine) effect only exists for maternal prenatal substance use. b) Polygenic risk score analysis was conducted with maternal genetic variants as proxies for prenatal smoking, alcohol and caffeine consumption (3 separate analyses, with polygenic risk scores specific to the substance used).

## Methods

### Study populations

We used data from three European prospective longitudinal birth cohorts: the Avon Longitudinal Study of Parents and Children (ALSPAC), Generation R (GenR) and the Norwegian Mother, Father and Child Cohort Study (MoBa). ALSPAC is a prospective longitudinal cohort study that recruited 14,541 pregnant women resident in Avon, UK with expected dates of delivery between 1st April 1991 and 31st December 1992 (24-26). GenR is a population-based prospective cohort study in Rotterdam in the Netherlands that recruited 9,778 pregnant women expected to give birth between April 2002 and January 2006 (27). MoBa is a population-based pregnancy cohort study where participants were recruited from all over Norway between 1999 and 2008. The cohort now includes 114,500 children, 95,200 mothers and 75,200 fathers (28). More details about each cohort and the representativeness of these cohorts with respect to smoking and alcohol use are shown in the Supplementary Material.

### Genome-wide genotype data

In ALSPAC, genome-wide data were available for 8,196 mothers. Maternal genetic data was not available for GenR at the time of analyses. In MoBa, genetic data were available for 14,584 mothers. Detailed information about the genotyping is presented in the Supplementary Material.

### Measures

#### Exposures

We used data assessed in the 2^nd^ pregnancy trimester where information for both maternal and paternal substance use was available. Briefly, mothers and fathers were asked about their average number of cigarettes smoked per day if they were smokers, average amount and frequency of alcohol consumption, and how many cups of caffeinated drinks (coffee, tea, cans of cola) per day on average they consumed during the first pregnancy trimester and mothers also before pregnancy. Overall, exposure assessment was similar across the cohorts, but there were some exceptions in GenR (Supplementary Table S1).

Parental prenatal smoking, alcohol and caffeine consumption (from coffee and tea) were categorized to examine dose-dependent relationships. Smoking was categorized: No smoking; 1-4 cigarettes; 5-9 cigarettes and >10 cigarettes per day. Alcohol consumption was categorized: No drinking; <1 drink a week and 1-6 drinks a week. Only a small number of mothers drank daily, therefore these were combined with the group of weekly drinkers. Furthermore, because the measure of alcohol unit was different in each cohort, meta-analysis across the cohorts was conducted comparing drinkers and non-drinkers. However, in ALSPAC and MoBa we were able to harmonise weekly alcohol consumption from units to grams to create a continuous measure of alcohol consumption (Supplementary Table S1). Caffeine consumption from coffee and tea was transformed and summed to total caffeine consumption in milligrams per day and categorized: 0-49mg; 50-199mg; 200-299mg and >300mg.

#### Outcome

ADHD symptoms were measured using different questionnaires around age 7-8 years in each cohort. In each cohort we used questionnaires that measured total ADHD symptoms, as well as hyperactivity and inattention symptom domains.

As the continuous score of ADHD symptoms was either zero-inflated or skewed, a binary variable was derived for total ADHD, hyperactivity and inattention symptoms using the 85^th^ percentile threshold to indicate a high risk of ADHD symptoms. This approach has been used in previous studies to describe how a child’s score compares with other children within the sample (29). The score within a range of 84^th^ to 90^th^ percentile represents children with higher level of symptoms although not necessarily at a level of diagnosis (30).

Up to 4 missing items were allowed depending on number of items in the questionnaire. More details are shown in Supplementary Table S2.

#### Primary outcome measures

The psychometric scales used for the main outcome measure were: maternal report of the Development And Well-Being Assessment (DAWBA) questionnaire in ALSPAC; maternal report of the revised Conner’s Parent Rating Scale (CPRS-R) in GenR; and maternal report of the Disruptive Behaviour Disorders scale (RS-DBD) in MoBa.

#### Secondary outcome measures

There is evidence of measurement differences of maternal and teacher reported ADHD symptoms in children (31), and some studies have found conflicting results depending on the questionnaire used (16). We therefore included additional questionnaires: teacher report of the DAWBA questionnaire and maternal and teacher report of the Strength and Difficulties Questionnaire (SDQ) hyperactivity subscale in ALSPAC; and maternal and teacher report of the Child Behaviour Checklist (CBCL) attention problems subscale in GenR.

The correlation between CPRS-R and RS-DBD has been reported to be high (r=0.7-0.8) (32), as has the correlation between SDQ and CBCL (r=0.7) (33). The correlation between DAWBA and SDQ scales in ALSPAC was moderate (r=0.6).

#### Covariates

Confounders were identified based on previous studies (34-38), such as child’s sex, ethnicity, parental age, education, depression and anxiety problems, financial difficulties, marital status and smoking, alcohol and caffeine use. The list of confounders as used in the analyses is shown in Table 1.

**Table 1.**
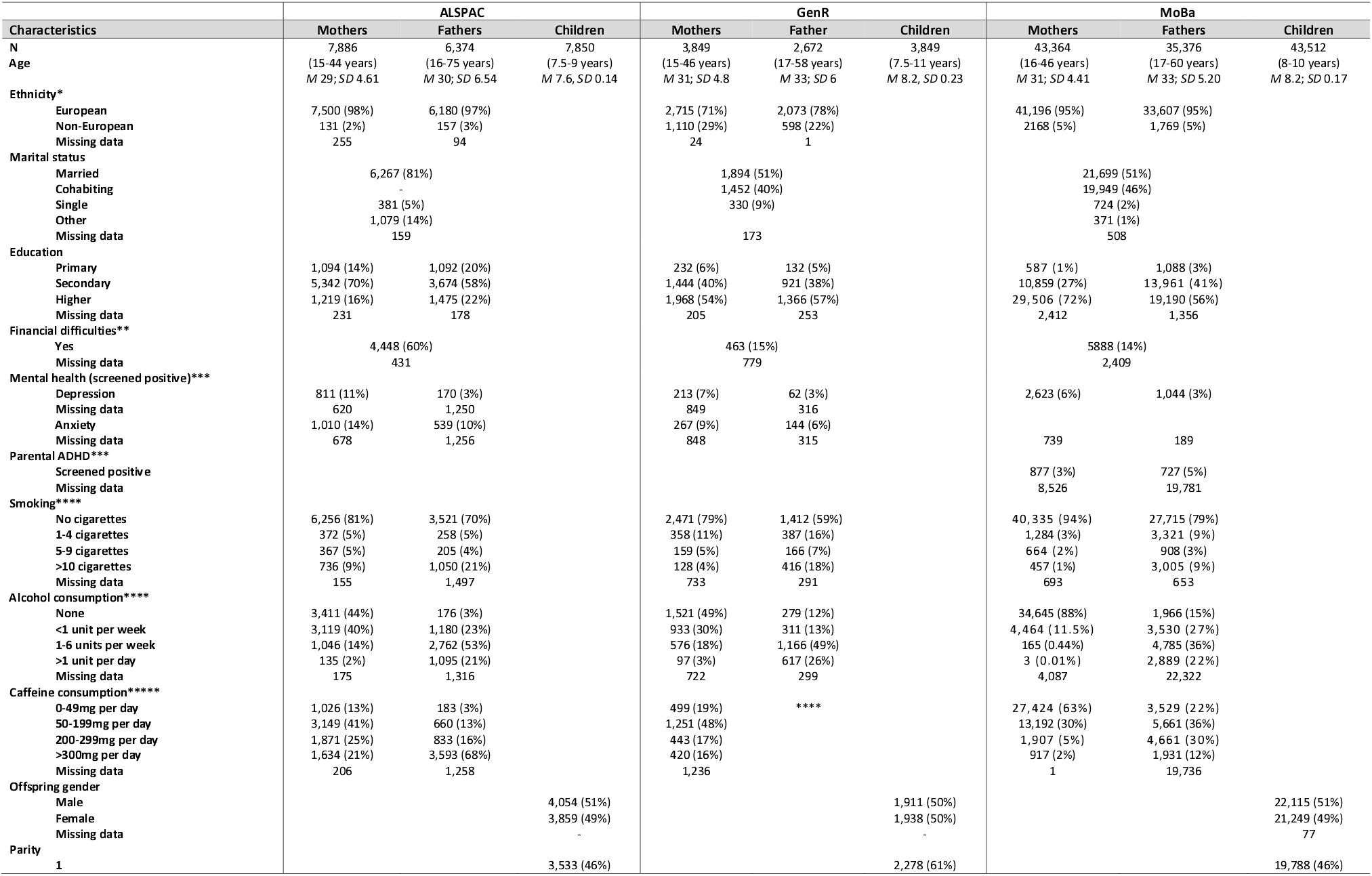
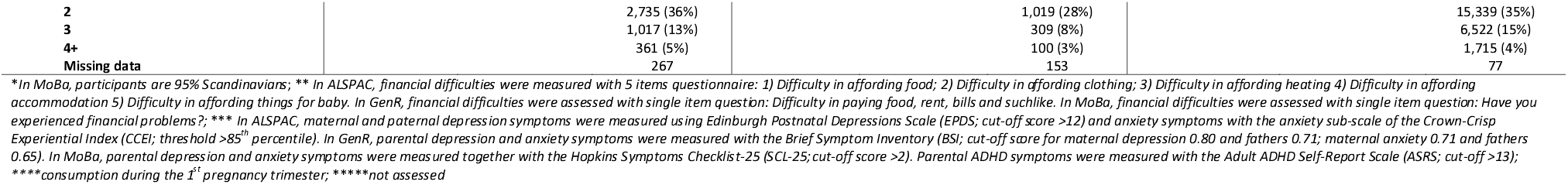
Overview of cohorts.

### Polygenic risk scores

PRS for mothers in ALSPAC and MoBa were calculated using genome-wide significant variants (p<5×10^−8^) and weighted by effect estimates (betas as our exposures were continuous traits) as reported in recent genome-wide association studies (GWAS) of tobacco, alcohol (39) and coffee consumption (40) using PLINK v1.90. More details about the phenotypes and SNPs discovered in these GWAS are shown in Supplementary Table S3. PRS for smoking heaviness was calculated using 49 single nucleotide polymorphisms (SNPs) available in ALSPAC and 51 SNPs available in MoBa and the sample was restricted to smokers during pregnancy. PRS for alcohol consumption was calculated with 90 SNPs available in ALSPAC and 92 SNPs available in MoBa and the sample was restricted to mothers who drank during pregnancy. PRS for caffeine consumption was calculated with 8 SNPs available in ALSPAC and 7 SNPs available in MoBa. There was some overlap between the SNPs included in the PRS for alcohol and caffeine, but no overlap between PRS for smoking and alcohol or caffeine. The correlation between these PRS were low (Supplementary Table S4).

### Statistical analyses

All analyses were performed using Stata (v15: ALSPAC, GenR; v16: MoBa), (41, 42). Analyses were performed as described in our pre-registered protocol (43). Analyses were conducted separately in each cohort and results from the primary outcome measure (maternal reported ADHD symptoms) were meta-analysed across the cohorts using a random effects model. This model takes into account the variance in the exposure and outcome assessment across the cohorts. The sample in each cohort was restricted to singletons in ALSPAC and GenR, whereas in MoBa a robust cluster variance estimator was used to account for the presence of siblings. No further clustering (i.e geographical clustering) was used in any of the cohorts. In ALSPAC and GenR paternal analyses were restricted to individuals who were reported as biological fathers. MoBa only includes fathers registered as the child’s parent. An overview of the analysis sample is shown in Table 1. We based our inference on consideration of whether the direction of effect observed is in the predicted direction, and the strength of statistical evidence against the null. We avoid claiming statistically significant effects, and instead treat the p-value as a continuous measure of statistical evidence (44).

#### Negative control analyses

Associations between maternal and paternal (negative control) exposures and high risk of offspring ADHD symptoms were tested using logistic regression analyses. We used three models: unadjusted (without including potential confounders); adjusted (including confounders) and mutually adjusted (adjusted for confounders and for partner’s substance use). Mutually adjusted models account for assortative mating, and there is evidence of this for health behaviours such as smoking and alcohol use (12, 45-47). In MoBa, because of the longer recruitment period, analyses were additionally adjusted for birth year. We also tested the difference between maternal and paternal associations for each exposure in ALSPAC and MoBa and for smoking and alcohol use in GenR.

#### PRS analyses

We investigated the association between maternal PRS and: 1) maternal exposure phenotypes to validate the PRS during pregnancy; 2) risk of offspring ADHD symptoms. PRS analyses in ALSPAC and MoBa were performed with adjustment for 10 ancestry-informative principal components. In MoBa, PRS analyses were additionally adjusted for birth year and genotyping batch. To explore potential pleiotropic effects, we also tested the association between the PRS and each confounder included in the negative control analysis.

### Sensitivity analyses

#### Negative control analyses

If an association was observed between maternal prenatal substance use and high risk of offspring ADHD symptoms, we further tested our hypothesis of a potential intrauterine effect by comparing maternal prenatal substance use with substance use before pregnancy. Given that ADHD is highly heritable, it is plausible that any observed associations between maternal PRS and offspring ADHD symptoms could be explained by genetic transmission. In MoBa, a measure of maternal ADHD symptoms was available, enabling us to test whether maternal ADHD symptoms could explain the observed associations between maternal substance use and offspring ADHD symptoms. Finally, we also performed complete case analyses to explore the impact of missing data by restricting unadjusted and adjusted analyses to the sample in the mutually adjusted model for each exposure. Using a complete case approach ensured that the same participants were included within each model and therefore enabled us to directly compare the effect estimates across analyses, to help evaluate any impact of missingness. The amount of missing data for each covariate is shown in Table 1. In order to avoid over-interpretation of results where there was no clear evidence of association, we focused our sensitivity analyses on those where there was some evidence of association in the main analyses.

#### PRS analyses

Unweighted PRS were calculated to test the association with each exposure phenotype, given that SNPs selected based only on the genome-wide significance level may be biased upwards (the so-called Winner’s Curse) (48). In addition to the PRS for smoking heaviness, we included a PRS for lifetime smoking. The lifetime smoking measure was derived by Wootton and colleagues and combines information on smoking initiation, duration, heaviness and cessation into a single phenotype (49). A GWAS of this composite lifetime smoking phenotype identified 126 independent SNPs at the genome-wide level of significance (p<5×10^−8^), of which 123 were available in ALSPAC and 121 in MoBa. This measure can be used without stratifying on smoking status. Finally, given that longitudinal studies may be subject to selection bias (50), we tested associations between PRS for smoking, alcohol and caffeine use and whether mothers returned the questionnaire at child age 7-8 in ALSPAC and MoBa.

## Results

Overall, the negative control analyses comparing maternal and paternal substance use associations with high risk of offspring ADHD symptoms showed mixed results across the cohorts. Stronger maternal associations were observed in MoBa, where mothers had lower prenatal smoking, alcohol and caffeine consumption compared to mothers in ALSPAC and GenR. The results of the meta-analysis and comparisons between maternal and paternal associations for the exposures in each cohort are shown in Figure 2. When testing for a difference between maternal and paternal substance use in each cohort individually, we found some evidence of a difference in the association between alcohol use (ALSPAC and MoBa) and caffeine use (MoBa only) and our outcomes. These findings were in line with those we observed in the meta-analysis. Our PRS analyses in ALSPAC and MoBa did not provide clear evidence for a causal effect of maternal prenatal substance use on offspring risk of ADHD symptoms. Furthermore, PRS analyses for lifetime smoking indicated pleiotropic associations with socio-demographic and mental health traits, as well as with returning questionnaires.

**Figure 2.**
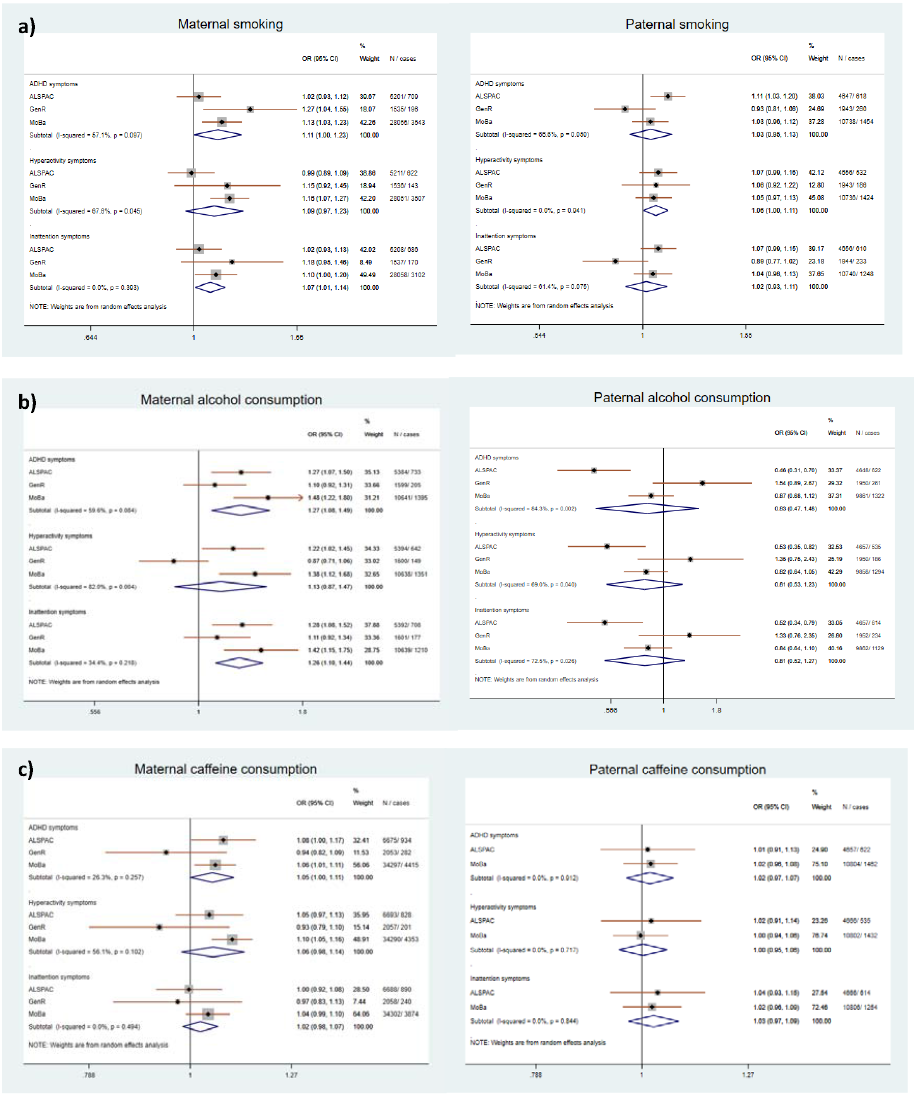
Meta-analysis of maternal and paternal prenatal smoking, alcohol and caffeine consumption across the cohorts. Note: Meta-analysis of smoking (a) and alcohol consumption (b) are based on mutually adjusted model. Meta-analysis of caffeine consumption (c) is based on adjusted model, because paternal caffeine consumption was not assessed in GenR. Heterogeneity between the cohorts is shown by computing I^2^ (see Methods and Supplementary Table S1 for more details). The statistical difference between maternal and paternal associations for smoking exposure in ALSPAC was (P_ADHD_=0.90; P_HYP_=0.91; P_INA_=0.34), in MoBa (P_ADHD_=0.14; P_HYP_=0.04; P_INA_=0.22) and in GenR (P_ADHD_=0.07; P_HYP_=0.79; P_INA_=0.10). The statistical difference between maternal and paternal associations for alcohol exposure in ALSPAC was (P_ADHD_=0.001; P_HYP_=<0.001; P_INA_=0.006), in MoBa (P_ADHD_=0.001; P_HYP_=0.005; P_INA_=0.002) and in GenR (P_ADHD_=0.75; P_HYP_=0.10; P_INA_=0.63) and the statistical difference between maternal and paternal associations for caffeine exposure in ALSPAC was (P_ADHD_=0.88; P_HYP_=0.99; P_INA_=0.68), in MoBa (P_ADHD_=0.05; P_HYP_=<0.001; P_INA_=0.47).

### Smoking

#### Negative control analyses

The pooled estimate for maternal smoking in the mutually adjusted model provided weak evidence of an association with high risk of ADHD total and inattention symptoms (OR_ADHD_=1.11, 95%CI 1.00, 1.23; OR_INA_=1.07, 95%CI 1.01, 1.14). A wide confidence interval was observed for hyperactivity symptoms (OR_HYP_=1.09, 95%CI 0.97, 1.23). For paternal smoking, there was some evidence of an association with high risk of hyperactivity symptoms (OR_HYP_=1.06, 95%CI 1.00, 1.11), but not with other ADHD outcomes (OR_ADHD_=1.03, 95%CI 0.95, 1.13; OR_INA_=1.02, 95%CI 0.93, 1.11). The results showing the dose-dependent relationship using non-smoking as baseline across unadjusted, adjusted and mutually adjusted models in each cohort are shown in Supplementary Tables S5-S7.

#### Sensitivity analyses

In MoBa, additional adjustment for maternal ADHD symptoms attenuated the association with high risk of offspring ADHD inattention symptoms, but there remained evidence of an association with high risk of ADHD total and hyperactivity symptoms (Supplementary Table S8), as well as between maternal smoking before pregnancy and high risk of hyperactivity symptoms (Supplementary Table S9).

Analyses using teacher report of DAWBA and SDQ scales in ALSPAC and TRF in GenR found no clear evidence of an association between maternal prenatal smoking and high risk of offspring ADHD symptoms (Supplementary Table S10). Although there was some evidence of an association when measuring total symptoms using the CBCL in GenR. This association was not observed for maternal smoking before pregnancy (Supplementary Table S11). The pattern of results was similar when restricting to complete cases (Supplementary Tables S12-S14).

#### PRS analyses

In each of the PRS analyses we report the results based on the assumptions described in the introduction.

First, the weighted and unweighted PRS for smoking heaviness and lifetime smoking were associated with smoking behaviour in pregnancy in ALSPAC and MoBa (all p<0.01). These PRS explained 1-3% of variance in smoking phenotypes in ALSPAC and MoBa (Supplementary Tables S15-S18).

Second, in ALSPAC, we did not find any strong evidence for an association between PRS for smoking heaviness and confounders included in the negative control analyses (Supplementary Table S19). However, in MoBa, we found evidence of an association between the PRS for smoking heaviness and lower parity (β=-0.41 95%CI −0.732, −0.092; Supplementary Table S20). The PRS for lifetime smoking was associated with younger maternal age (β=-2.64, 95%CI −3.688, −1.586), lower education (β=-1.00, 95%CI −1.286, −0.711), more financial difficulties (β=1.12, 95%CI 0.317, 1.912), higher likelihood of being single (OR=0.24, 95%CI 0.138, 0.415) and having more severe anxiety symptoms (OR=1.98, 95%CI 1.035, 3.801) in ALSPAC (Supplementary Table S21). Similarly, in MoBa, the PRS for lifetime smoking showed evidence of an association with lower maternal education (β=-0.27, 95%CI −0.356, − 0.191) and higher likelihood of having more severe depression and anxiety symptoms (OR=1.98, 95%CI 1.052, 3.705; Supplementary Table S22).

Third, in ALSPAC, we did not find clear evidence of an association between the PRS for smoking heaviness and high risk of maternal or teacher reported offspring ADHD symptoms (Supplementary Table S23 & S24). Similarly, in MoBa, there was no conclusive evidence of an association between the PRS for smoking heaviness and high risk of offspring ADHD symptoms (Supplementary Table S25). In contrast, we found no clear evidence of an association between the PRS for lifetime smoking and high risk of maternal reported offspring ADHD symptoms in ALSPAC (Supplementary Table S26), but we did find some evidence of an association with high risk of teacher reported ADHD total symptoms measured with both the DAWBA (OR_DAWBA_ = 2.70, 95%CI 1.026, 7.079) and the SDQ (OR_SDQ_ = 3.00, 95%CI 1.034, 8.688; Supplementary Table S27). There was no clear evidence of an association between maternal PRS for lifetime smoking and high risk of maternal reported ADHD symptoms in MoBa (Supplementary Table S28).

### Alcohol

#### Negative control analyses

The pooled estimate of maternal alcohol consumption in the mutually adjusted model showed some evidence of an association with high risk of ADHD total and inattention symptoms (OR_ADHD_=1.27, 95%CI 1.08, 1.49; OR_INA_=1.26, 95%CI 1.10, 1.44), but not with hyperactivity symptoms (OR_HYP_=1.13, 95%CI 0.87,1.47). The strongest associations were observed in ALSPAC and MoBa, in GenR the estimates were in opposite direction for high risk of hyperactivity symptoms. Meta-analysis of paternal alcohol consumption did not show clear evidence of an association with high risk of ADHD symptoms (OR_ADHD_=0.83, 95%CI 0.47, 1.48; OR_HYP_=0.81, 95%CI 0.53,1.23; OR_INA_=0.81, 95%CI 0.52,1.27), but there was high heterogeneity and confidence intervals were wide. The results across unadjusted, adjusted and mutually adjusted models in each cohort are shown in Supplementary Tables S29-S31.

#### Sensitivity analyses

We were not able to report dose-dependent results after adjustment for maternal ADHD symptoms in MoBa due to low numbers of cases (Supplementary Table S32). Additional sensitivity analyses (alcohol use before pregnancy and weekly alcohol use in grams) in ALSPAC and MoBa, and secondary outcome measures in ALSPAC did not find conclusive evidence for an association between maternal prenatal alcohol use and high risk of offspring ADHD symptoms (Supplementary Tables S33-S37). The pattern of results was similar when restricting to complete cases (Supplementary Tables S38-S40).

#### PRS analyses

First, in ALSPAC, the PRS for alcohol consumption was associated with prenatal alcohol consumption (Supplementary Table S15 & S17). However, in MoBa, the PRS for alcohol consumption did not predict alcohol consumption during pregnancy (β=-0.65, 95%CI −0.757, 2.055), although it was associated with alcohol consumption before pregnancy (β=1.06, 95%CI 0.258, 1.859) (Supplementary Table S16 & S18). The alcohol PRS explained 2% of variance in alcohol phenotype during pregnancy in ALSPAC and 0.7% variance in alcohol phenotype before pregnancy in MoBa.

Second, the PRS for alcohol consumption was associated with higher maternal education (β=0.52, 95%CI 0.058, 0.983) and a higher likelihood of having more severe depression symptoms (OR=3.42, 95%CI 1.058, 11.047) in ALSPAC (Supplementary Table S41). However, no clear evidence for an association between the PRS for alcohol consumption and confounders was found in MoBa (Supplementary Table S42).

Third, we found no conclusive evidence of an association between maternal PRS for alcohol consumption and either high risk of maternal or teacher reported offspring ADHD symptoms in ALSPAC, or with maternal reported ADHD symptoms in MoBa (Supplementary Tables S43-S45).

### Caffeine

#### Negative control analyses

The pooled estimate of maternal caffeine consumption in the adjusted model showed some evidence of an association only with high risk of offspring ADHD total symptoms (OR_ADHD_=1.05, 95%CI 1.00, 1.11; OR_HYP_=1.06, 95%CI 0.98, 1.14; OR_INA_=1.02, 95%CI 0.98, 1.07), whereas the meta-analysis of paternal caffeine consumption in ALSPAC and MoBa did not (OR_ADHD_=1.02, 95%CI 0.97, 1.07; OR_HYP_=1.00, 95%CI 0.95, 1.06; OR_INA_=1.03, 95%CI 0.97, 1.09). Cohort specific results are shown in Supplementary Tables S46-S48.

#### Sensitivity analyses

Sensitivity analyses in ALSPAC and MoBa did not find clear evidence for an association between maternal prenatal caffeine consumption and high risk of offspring ADHD symptoms (Supplementary Tables S48-S51). The pattern of results was similar when restricting to complete cases (Supplementary Tables S52-S54).

#### PRS analyses

First, both the weighted and unweighted PRS for caffeine consumption were associated with total caffeine consumption derived from coffee and tea in ALSPAC and MoBa. The caffeine PRS explained 0.3-0.4% of variance in caffeine phenotype in ALSPAC and MoBa (Supplementary Tables S15-S18).

Second, we found no clear evidence of an association between the PRS for caffeine consumption and the confounders in ALSPAC or MoBa (Supplementary Tables S55-S56).

Third, we found no clear evidence of an association between maternal PRS for caffeine consumption and either high risk of maternal or teacher reported offspring’s ADHD symptoms in ALSPAC or with maternal reported ADHD symptoms in MoBa (Supplementary Tables S57-S59).

Results from the PRS meta-analysis in ALSPAC and MoBa are shown in Supplementary Table S60.

### Associations between PRS for substance use and participation at age 8 years

We found evidence of an association between the PRS for lifetime smoking and lower likelihood of returning the questionnaire at age 7-8 years in ALSPAC and MoBa (OR_ALSPAC_ = 0.49, 95%CI 0.311, 0.757; OR_MOBA_= 0.59, 95%CI 0.427, 0.801). Furthermore, in MoBa the PRS for smoking heaviness was associated with higher likelihood of returning the questionnaire (OR_MOBA_ = 2.10, 95%CI 1.01, 4.359), but a similar association was not observed in ALSPAC (OR_ALSPAC_=0.95; 95% CI 0.561, 1.607) (Supplementary Tables S61-S62).

## Discussion

We investigated whether maternal smoking, alcohol and caffeine use during pregnancy are likely to be causally associated with high risk of offspring ADHD symptoms. We applied a triangulation approach using negative control and PRS analyses in three longitudinal birth cohorts. Overall, the results did not provide evidence for a potential causal effect between maternal prenatal substance use and high risk of offspring ADHD symptoms although some inconsistencies were observed across the cohorts and instrument used for ADHD assessment. In contrast to some previous studies our results also did not find evidence for distinct effect of maternal prenatal substance use on ADHD symptom domains.

Our smoking results did not show robust evidence for a causal effect, which is in line with previous findings (37, 51-53). Although in GenR and MoBa, we found suggestive evidence for a causal relationship of maternal prenatal smoking on high risk of maternal reported ADHD symptoms, but when comparing the findings across the cohorts, reporters and questionnaires, the evidence was weak and inconsistent. Additionally, our PRS analyses with lifetime smoking PRS in ALSPAC and MoBa indicated pleiotropic associations which are consistent with recent findings in ALSPAC (54). There is also a large body of evidence showing pleiotropy between smoking, impulsivity and sensation-seeking type of personality (55, 56) which could confound observed phenotype associations in the present study.

Similarly, our findings on prenatal alcohol and caffeine exposure do not show evidence of a causal effect on offspring ADHD symptom risk. Although a previous study in MoBa found some evidence for a potential causal effect of maternal prenatal alcohol consumption when ADHD symptoms were measured with CPRS-R (16), other studies suggest that observed associations between maternal moderate prenatal alcohol consumption and offspring ADHD symptoms may not reflect causal effects (3, 57). Our results on caffeine exposure are in line with previous studies which have concluded that there is likely no causal effect of prenatal caffeine consumption on offspring ADHD symptom risk (6, 58, 59).

Several studies have reported low to moderate parent-teacher agreement on ADHD symptoms assessment (31, 60). It has been suggested that parents and teachers may measure different aspects of child’s behaviour as ADHD symptoms may be more visible at school which is a more structured environment (31). Furthermore, it has been proposed that parent-teacher ratings may differ because of the informant’s perception and individual characteristics (61). It has been shown that mothers with mental health problems or more harsh parenting behaviour overestimate their child’s mental health problems (62, 63). Given that we observed more associations with maternal report than with teacher report, it is possible that observed associations may be confounded by maternal characteristics.

Besides reporter-related discrepancies, we observed different findings depending on the scales used for ADHD assessment. Previous studies investigating the association between maternal prenatal substance use and offspring ADHD have reported Inconsistent findings depending on which scale was used for ADHD symptoms assessment. For example, a study using the SDQ scale reported association between maternal prenatal smoking and ADHD symptoms in children regardless of the reporter (64). Another study using maternal and teacher reported CPRS-R, CBCL, TRF and combined score of CBCL/TRF found some evidence for a potential causal relationship between maternal prenatal smoking and ADHD symptoms only with maternal reported CPRS-R (65). Similarly, a study on prenatal alcohol exposure found some evidence for a causal effect when ADHD symptoms were assessed with maternal reported CPRS-R but not with CBCL (16). Although all the scales for our main outcome measure (DAWBA, CPRS-R, RS-DBD) are based on the Diagnostic and Statistical Manual of Mental Disorders (DSM-IV) criteria for ADHD, we observed inconsistent associations between maternal prenatal substance use and ADHD symptom risk across different scales. It is possible that different scales capture somewhat different aspects of the construct of ADHD.

### Strengths and limitations

The major strength of the current study is the triangulation approach using both observational and genetic analyses, as well as including multiple questionnaires reported by mother and teacher. Using data from three large longitudinal birth cohorts strengthens evidence towards causal interpretation.

However, our study has also limitations. First, outcome assessment varied across the cohorts which may contribute to noise and inconsistent findings. Although all the questionnaires have good psychometric properties, there still may be a risk of measurement error. Second, maternal prenatal substance use was based on self-reports, which most likely lead to underreporting of substance use. In addition, in some cases we observed stronger associations with maternal reported ADHD symptoms than teacher reported ADHD symptoms. Since our exposures were also maternally reported, it is possible that the associations with maternal reported outcomes were inflated due to common method bias. Third, our negative control analyses were based on the assumption that confounders affecting maternal and paternal substance use follow similar patterning, but this assumption may in fact be more likely for some of the substances than others. However, considering that we included cohorts with different cultural backgrounds, as well as applied a PRS approach which is subject to different sources of bias, we have more confidence that our results are less affected by violation of the assumption of the same confounding in maternal and paternal substance use. Fourth, the range of alcohol consumption was restricted in our sample, and therefore we cannot interpret our findings outside of this range. There may be effects of heavier consumption that we were not able to detect. Fifth, in our sample ADHD symptoms were assessed around ages 7-8 years and it is possible that some ADHD symptoms are not fully expressed by that age. This may lead to misclassification of these children as not having high risk of ADHD symptoms. Sixth, our PRS for smoking heaviness and alcohol consumption were calculated based on summary statistics from the latest GWAS which included ALSPAC. However, the contribution of ALSPAC (∼1%) was small and the risk of bias because of the sample overlap is likely to be minimal (66). Seventh, our PRS analyses were likely underpowered. Compared to the variance explained by each PRS reported in GWAS (smoking heaviness PRS ∼4%; alcohol PRS ∼2.5%; caffeine PRS 1.3%), in our sample it was much smaller. Eighth, our sample in the smoking and alcohol PRS analyses was restricted to users as in the discovery GWAS, but this may introduce bias given that non-users were excluded. Nineth, the sample sizes in our fully adjusted models were reduced due to missing data in the included confounders which could introduce bias into our estimates. Although, we repeated all analyses restricting to individuals in our mutually adjusted models and effect estimates remained consistent, there still may be a bias because of study selection and attrition. Tenth, longitudinal cohort studies may suffer from selection bias as socioeconomic and individual characteristics may affect initial and continued participation in the study (67, 68). A study in MoBa found that bias due to self-selection and loss to follow-up can influence exposure-outcome associations (69). Another study in ALSPAC showed that common genetic variants of various phenotypes are associated with participation in the study and these associations differ in the sample with full genetic data and more selected subsamples (50). Given that attrition in our study samples was around 50% and we also observed association between PRS for lifetime smoking and decreased likelihood returning the questionnaire at child’s age 7-8 years, it is plausible that our results may be subject to selection bias. Eleventh, our results should be interpreted in the context of the number of statistical tests performed. Depending on the cohort, we performed 9 to 15 tests for each exposure in each cohort. We avoid using a p-value threshold to claim statistically significant effects, and instead treat the p-value as a continuous measure of statistical evidence.

## Conclusion

Combining both observational and genetic analyses from three longitudinal birth cohorts our study did not find support for a causal effect of maternal smoking, alcohol and caffeine consumption during pregnancy on high risk of offspring ADHD symptoms. Although in this study we did not find strong evidence for a causal effect of maternal prenatal substance use on offspring high risk of ADHD symptoms, prenatal substance use can still have deleterious effects on other child health outcomes. Considering that even small effects can be important at a population level, pregnant women should abstain from smoking and alcohol use during pregnancy and keep their caffeine consumption minimal.

## Data Availability

The informed consent obtained from ALSPAC participants does not allow the data to be made freely available through any third party maintained public repository. However, data used for this submission can be made available on request to the ALSPAC Executive. The ALSPAC data management plan describes in detail the policy regarding data sharing, which is through a system of managed open access. In MoBa, the consent given by the participants does not open for storage of data on an individual level in repositories or journals. Researchers who want access to data sets for replication should submit an application to. Access to data sets requires approval from The Regional Committee for Medical and Health Research Ethics in Norway and an agreement with MoBa. In GenR, to ensure participant privacy and law compliance, individual-level data cannot be made publicly available without explicit informed consent, which is not available. For new analyses or individual-level data access, please contact Generation R data management

http://www.bristol.ac.uk/alspac/researchers/access/

http://www.bristol.ac.uk/alspac/researchers/our-data/

https://generationr.nl/researchers/collaboration/

## Acknowledgements

We are extremely grateful to all the families who took part in this study, the midwives for their help in recruiting them, and the whole ALSPAC team, which includes interviewers, computer and laboratory technicians, clerical workers, research scientists, volunteers, managers, receptionists and nurses. The UK Medical Research Council and Wellcome (Grant ref: 217065/Z/19/Z) and the University of Bristol provide core support for ALSPAC. A comprehensive list of grants funding is available on the ALSPAC website (http://www.bristol.ac.uk/alspac/external/documents/grant-acknowledgements.pdf). GWAS data was generated by Sample Logistics and Genotyping Facilities at Wellcome Sanger Institute and LabCorp (Laboratory Corporation of America) using support from 23andMe.

The Norwegian Mother, Father and Child Cohort Study is supported by the Norwegian Ministry of Health and Care Services and the Ministry of Education and Research. We are grateful to all the participating families in Norway who take part in this on-going cohort study. We thank the Norwegian Institute of Public Health (NIPH) for generating high-quality genomic data. This research is part of the HARVEST collaboration, supported by the Research Council of Norway (NRC) (#229624). We also thank the NORMENT Centre for providing genotype data, funded by NRC (#223273), South East Norway Health Authority and KG Jebsen Stiftelsen. Further we thank the Center for Diabetes Research, the University of Bergen for providing genotype data and performing quality control and imputation of the data funded by the ERC AdG project SELECTionPREDISPOSED, Stiftelsen Kristian Gerhard Jebsen, Trond Mohn Foundation, NRC, the Novo Nordisk Foundation, the University of Bergen, and the Western Norway health Authorities (Helse Vest).

The general design of Generation R Study is made possible by financial support from the Erasmus Medical Center, Rotterdam, the Erasmus University Rotterdam, the Netherlands Organization for Health Research and Development (ZonMw), the Netherlands Organisation for Scientific Research (NWO), the Ministry of Health, Welfare and Sport and the Ministry of Youth and Families. The Generation R Study is conducted by the Erasmus Medical Center in close collaboration with the School of Law and Faculty of Social Sciences of the Erasmus University Rotterdam, the Municipal Health Service Rotterdam area, Rotterdam, the Rotterdam Homecare Foundation, Rotterdam and the Stichting Trombosedienst & Artsenlaboratorium Rijnmond (STAR-MDC), Rotterdam. We gratefully acknowledge the contribution of children and parents, general practitioners, hospitals, midwives and pharmacies in Rotterdam. The generation and management of GWAS genotype data for the Generation R Study were done at the Genetic Laboratory of the Department of Internal Medicine, Erasmus MC, The Netherlands. We would like to thank Karol Estrada, Dr. Tobias A. Knoch, Anis Abuseiris, Luc V. de Zeeuw, and Rob de Graaf, for their help in creating GRIMP, BigGRID, MediGRID, and Services@MediGRID/D-Grid, (funded by the German Bundesministerium fuer Forschung und Technology; grants 01 AK 803 A-H, 01 IG 07015 G) for access to their grid computing resources. We thank Mila Jhamai, Manoushka Ganesh, Pascal Arp, Marijn Verkerk, Lizbeth Herrera and Marjolein Peters for their help in creating, managing and QC of the GWAS database. Also, we thank Karol Estrada for their support in creation and analysis of imputed data.

We would like to thank Sonja Swanson for her valuable ideas when designing the study and Claudia Kruithof for her contribution.

