## Supplementary material for "Prenatal smoking, alcohol and caffeine exposure and maternal reported ADHD symptoms in childhood: triangulation of evidence using negative control and polygenic risk score analyses"

### Supplementary methods.

### Study populations

In **ALSPAC**, the initial number of pregnancies enrolled is 14,541 and of these initial pregnancies, there was a total of 14,676 fetuses, resulting in 14,062 live births and 13,988 children who were alive at 1 year of age. When the oldest children were approximately 7 years of age, an attempt was made to bolster the initial sample with eligible cases who had failed to join the study originally, resulting in an additional 913 children being enrolled. The total sample size for analyses using any data collected after the age of seven is therefore 15,454 pregnancies, resulting in 15,589 fetuses. Of these 14,901 were alive at 1 year of age.

The ALSPAC sample was broadly representative of Great Britain (GB) at the time of data collection; however, parents in ALSPAC were less likely to live in rented accommodation, and fathers were less likely to have a manual occupation compared with the rest of GB (Golding et al., 2001). Regarding prenatal alcohol use, 79% of women in UK drank alcohol during pregnancy (O’Keeffe et al., 2015), whereas in our sample it was 56% at the sample available for analyses. For smoking, it has been shown that in 1986 the prevalence of prenatal smoking in UK was 37% (Madeley et al., 1989), whereas in our baseline sample 25% of mothers reported smoking during pregnancy.

The ALSPAC study was approved by the ALSPAC Ethics and Law Committee and the Local Research Ethics Committees and informed consent for the use of data collected via questionnaires and clinics was obtained from participants. The study website contains details of all the data that is available through a fully searchable data dictionary and variable search tool: http://www.bristol.ac.uk/alspac/researchers/our-data/

**GenR** is a population-based prospective cohort study in Rotterdam in the Netherlands designed to investigate environmental and genetic causes of health and development from fetal life until young adulthood. GenR recruited in total 9,778 pregnant women who were expected to give a birth between April 2002 and January 2006 in Rotterdam. Of all eligible children at birth, 61% agreed to participate in the study. The final baseline sample size included 9,749 children and 80% of the sample has been followed up until age 13 years (Kooijman et al., 2016). The GenR is a multi-ethnic cohort and besides Dutch ethnicity other larger ethnic groups are Surinamese, Turkish and Moroccan.

In GenR, at baseline sample, parents had a higher socio-economic status compared with the general population in the study area (Rotterdam) (Jaddoe et al., 2008). In the Netherlands the prevalence of prenatal alcohol use in 2002-2005 was 36-37% (Bakker et al., 2010), whereas in our baseline sample 43% of mothers reported drinking during pregnancy. Regarding smoking, the prevalence of prenatal smoking in 2002-2005 was about 10% and was highest among mothers with low level of education (18-22%) (Lanting et al., 2012), whereas in our baseline sample 20% of pregnant women reported smoking.

The study was approved by the local Medical Ethical Committee (MEC 198.782/2001/31). Written informed consent was obtained from all participating women.

**MoBa** is a population-based pregnancy cohort study conducted by the Norwegian Institute of Public Health. Participants were recruited from all over Norway from 1999-2008. The women consented to participation in 41% of the pregnancies. The cohort now includes 114.500 children, 95.200 mothers and 75.200 fathers.

In MoBa, according to the Medical Birth Registry of Norway in 2000-2006, women <25 years, those living alone, and smokers were underrepresented (Nilsen et al., 2009). It has been shown that in 1999-2001 the prevalence of prenatal smoking was 17% and in 2002-2004 13%, and prenatal smoking was highest among multiparous (3+), teenage and single mothers and among those with lower education (Kvalvik et al., 2008). In our sample in 1999-2001 14% and in 2002-2004 11% of pregnant women reported smoking during pregnancy. In addition, in 2005-2007 the smoking rate in our sample was 7% and in 2008-2009 5%. Furthermore, it has been reported that average prenatal alcohol consumption in Norway in year 2011-2012 was 4% (Mårdby et al., 2017). A case-control study conducted in 1996-2001 showed that among control mothers 30% reported drinking during the first pregnancy trimester (DeRoo et al., 2008). In our sample in 1999-2001 25% of mothers reported drinking during the first pregnancy trimester and in 2008-2009 the rate was 6%. Additionally, in 2002-2004 the rate of prenatal alcohol use was 16% and in 2005-2007 10%.

The current study is based on version 12 of the quality-assured data files released for research on January 2019. The establishment of MoBa and initial data collection was based on a license from the Norwegian Data Protection Agency and approval from The Regional Committees for Medical and Health Research Ethics. The MoBa cohort is now based on regulations related to the Norwegian Health Registry Act. The current study was approved by The Regional Committees for Medical and Health Research Ethics (2016/1702).

### Genome-Wide Data and Quality Control

**Avon Longitudinal Study of Parents and Children (ALSPAC)**

DNA samples were collected from 11,343 children and 10,015 mothers.

ALSPAC children were genotyped using the Illumina HumanHap550 quad chip genotyping platforms. Individuals were excluded on the basis of gender mismatches; minimal or excessive heterozygosity; disproportionate levels of individual missingness (>3%) and insufficient sample replication (IBD < 0.8). SNPs with a minor allele frequency of < 1%, a call rate of < 95% or evidence for violations of Hardy-Weinberg equilibrium (p < 10^-7^) were removed. Related subjects that passed all other quality control thresholds were retained during subsequent phasing and imputation. 9,115 children and 500,527 SNPs passed these quality control filters.

ALSPAC mothers were genotyped using the Illumina human660W-quad array at Centre National de Génotypage (CNG) and genotypes were called with Illumina GenomeStudio. SNPs were removed if they displayed more than 5% missingness or a Hardy-Weinberg equilibrium (p < 10^-6^). Additionally, SNPs with a minor allele frequency of less than 1% were removed. Samples were excluded if they displayed more than 5% missingness, had indeterminate X chromosome heterozygosity or extreme autosomal heterozygosity.

Related subjects that passed all other quality control thresholds were retained during subsequent phasing and imputation. 9,048 mothers and 526,688 SNPs passed these quality control filters.

Population stratification in mothers and children were compared with Hapmap II (release 22) European descent (CEU), Han Chinese, Japanese and Yoruba reference populations; all individuals with non-European ancestry were removed.

After combining genotype data in the mothers and the children, SNPs with genotype missingness above 1% were removed due to poor quality (11,396 SNPs removed) and a further 321 subjects were removed due to potential ID mismatches. This resulted in a dataset of 17,842 subjects. Imputation of the target data was performed using Impute V2.2.2 against the 1000 genomes reference panel (Phase 1, Version 3) (all polymorphic SNPs excluding singletons), using all 2186 reference haplotypes (including non-Europeans).

The final dataset included 8,237 children and 8,196 mothers.

More details about the genotyping and quality control procedure can be found (Paternoster et al., 2011; Taylor et al., 2018).

**Norwegian Mother, Father and Child cohort (MoBa)**

Approximately 17,000 trios from the Norwegian Mother, Father and Child cohort were genotyped in three batches. The first batch, comprising 20,664 individuals and 542,585 SNPs was genotyped at the NTNU Genomics Core Facility (Trondheim, Oslo) using the Illumina HumanCoreExome (Illumina, San Diego, USA) genotyping array, version 12 1.1. The second batch, comprising 12,874 individuals and 547,644 SNPs was genotyped at the NTNU Genomics Core Facility (Trondheim, Oslo) using the Illumina HumanCoreExome (Illumina, San Diego, USA) genotyping array, version24 1.0. The third batch, comprising 17,949 individuals and 692,367 SNPs, was genotyped at ERASMUS MC (the Netherlands) using the Illumina Global Screening Array (Illumina, San Diego, USA) version 24 1.

Individuals were excluded if they had a genotyping call rate below 95% or autosomal heterozygosity greater than four standard deviations from the sample mean. SNPs were excluded if they were ambiguous (A / T and C / G), had a genotyping call rate below 98%, minor allele frequency of less than 1%, or Hardy-Weinberg equilibrium P-value less than 1 × 10^-6^. Relatedness was assessed by flagging one individual from each pairwise comparison of identity-by-descent with a pi-hat greater than 0.1.

Population stratification was assessed, using the HapMap phase 3 release 3 as a reference by principal component analysis using EIGENSTRAT version 6.1.4. Visual inspection identified a homogenous population of European ethnicity and individuals of non-European ethnicity were removed.

Duplicate samples were removed, and each genotyping batch was split into parents and offspring. Quality control was then conducted by genotyping array in parents and offspring’s separately.

The parents and offspring’s datasets were then merged into one dataset per genotyping batch; keeping only the SNPs that passed quality control in both datasets. All individuals passing the genotyping call rate and autosomal heterozygosity measures were included in the merged datasets. Therefore, the merged datasets included individuals previously excluded or flagged as a duplicate, ethnic outlier, having a sex discrepancy, or high level of relatedness. Concordance checks were then conducted on validated duplicates. Duplicate, tri-allelic and discordant (any discordance between the validated duplicates) SNPs were excluded. Individuals and SNPs with a genotyping call rate below 98% in the merged datasets were excluded. The duplicate sample that was removed before the start of the quality control was then excluded. Mendelian errors identified by the assessment of duos and trios were then recoded to missing. Insertions and deletions were also excluded.

After QC the Human Core Exome 12 batch comprised 20,231 individuals and 384,855 SNPs, the Human Core Exome 24 batch comprised 12,757 individuals and 396,189 SNPs, and the Global Screening Array batch comprised 17,742 individuals and 568,275 SNPs. Imputation was conducted separately for each genotyping batch by using the Haplotype reference consortium (HRC) release 1-1 as the genetic reference panel.

Post imputation quality control was performed by removing individuals if they had a genotyping call rate less than 99% or were of non-European ethnicity. After quality control, a core homogeneous sample of European ethnicity (based on PCA of markers overlapping with available HapMap markers), unrelated (within generation, defined as accumulated identity-by-descent <0.015 and overall identity-by-descent PI_HAT <10%) individuals across all batches and arrays were available for use in analysis (N_children_ = 15,208; N_mothers_ = 14,804; N_fathers_ = 15,198).

More details about the genotyping and quality control procedure has been reported elsewhere (Helgeland et al., 2019).

### Supplementary Table S1. Exposure Assessment

|  | **ALSPAC** | **GenR** | **MoBA** |
| --- | --- | --- | --- |
| **Exposure** | 18 weeks of gestation | 18-25 weeks of gestation | 17 weeks of gestation |
| **Smoking** | Mothers were asked if they were smokers and how many cigarettes per day, they smoked before pregnancy and first three months of the pregnancy  (“*Have you ever been a smoker”; “Number of cigarettes smoked before pregnancy, first 3 months of pregnancy and past 2 weeks”)*  Fathers were asked about their average daily smoking at the start of their partner’s pregnancy  *(“Number of cigarettes smoked at the start of their partner’s pregnancy and in the last 2 weeks”)* | Mothers were asked about their average quantity of daily smoking before pregnancy and in the past 2 months  *(“Number of cigarettes smoked in the past 2 months”*)  Fathers were asked about their quantity of smoking 2 months prior partner’s pregnancy  *(“Number of cigarettes smoked 2 months before their partner’s pregnancy”)* | Mothers were asked if they had been smoking before and during pregnancy and about their average quantity of daily and weekly smoking.  *(“Number of cigarettes smoked per day and week before pregnancy and after pregnancy was known”)*  Fathers were asked about their smoking during their partner’s pregnancy  *(“Number of cigarettes smoked after their partner become pregnant”)* |
| **Alcohol** | Mothers were asked about their average amount and frequency of alcohol consumption before and during first three months of the pregnancy (*“Amount of alcohol consumption (in units) before current pregnancy, during first 3 months of pregnancy and at the time when they first felt the baby move, per day/week”*)  Fathers were asked about their alcohol consumption in past three months.  (“*Amount of alcohol consumption first 3 months of their partner’s pregnancy, units per day/week”)*  *1 drink corresponds to 8 grams of pure alcohol (Alati et al., 2013) | Mothers were asked about their alcohol use and frequency before pregnancy and in the past 3 months.  (“*Number of glasses of alcohol consumed in the past 2 months (per week/month”)*  Fathers were asked about their average alcohol consumption 2 months prior their partner’s pregnancy.  *(“Number of glasses of alcohol consumed per week or day 2 months before their partner’s pregnancy”)*  *1 alcoholic drink corresponds to 12 grams of pure alcohol (Bakker et al., 2010) | Mothers were asked about their frequency and amount of alcohol use before and during the current pregnancy.  *(“Frequency and number of units of alcohol consumed 3 months before pregnancy and during pregnancy per week/month”)*  Fathers were asked about their alcohol use during their partner’s pregnancy  *(“Frequency and number of units of alcohol consumed 6 months before partner’s pregnancy and after partner become pregnant per week/month”)*  *1 alcoholic drink corresponds to 12.8 grams of pure alcohol (Knudsen et al., 2014) |
|  | Additionally, in ALSPAC a continuous measure of alcohol use was available at 8 weeks of gestation. To harmonise weekly alcohol consumption from units to grams, a measure of units was multiplied with corresponding grams in ALSPAC and MoBa to create a continuous measure of alcohol consumption | | |
| **Caffeine** | Mothers and fathers were asked about type (caffeinated, decaffeinated or both) and amount of coffee, tea and cola consumption per day, during the week and weekend during pregnancy  (“*Number of cups of coffee, tea and cola consumed per day during the week and weekend”;* “*Type of drink consumed (caffeinated, decaffeinated or both)”*)  *1 cup of coffee = 75mg; 1 cup of tea = 40mg and 1 can of cola = 33mg of caffeine (Treur et al., 2016) | Mothers were asked about type and amount of coffee and tea consumption per day.  *(“Number of cups of coffee and tea consumed daily before pregnancy and in the past 2 months of pregnancy”; “Type of drink consumed (caffeinated, decaffeinated or both”)*  Caffeine consumption in fathers was not assessed.  *1 cup of coffee = 90mg; 1 cup of decaffeinated coffee 3mg; 1 cup of tea = 45mg of caffeine (Voerman et al., 2016) | Mothers were asked about their daily amount of coffee, tea and soft drinks consumption before and during pregnancy.  *(“Number of cups/glasses of caffeinated and decaffeinated coffee, tea and cola consumption per day”)*  Fathers were asked about their average daily amount of caffeine consumption.  *(“How often consumed coffee, tea and cola weekly and daily”)*  *1 cup of filtered coffee = 85mg; 1 cup of espresso coffee = 60mg; 1 cup of tea = 50mg and 1 can of soft drink = 30mg (Papadopoulou et al., 2018) |

### Supplementary Table S2. ADHD assessment

| **Cohort** | **Instrument** |
| --- | --- |
| **ALSPAC** | **Maternal and teacher report of The Development And Well-Being Assessment (DAWBA)** at age 7.5 years. The DAWBA was designed to generate psychiatric diagnoses based on questionnaires and interviews and brings together different sources of information for predicting psychiatric problems in children and adolescence based on DSM-IV and ICD-10 criteria (Goodman et al., 2000). It consists of 18 items that are measuring ADHD and separately inattentive and hyperactive/impulsive symptom domains. The items are assessed in the 3-point Likert scale with a degree of symptom: 0=no more than other; 1=a little more than others and 2=lot more than others. In ALSPAC, a questionnaire was used.  **Maternal and teacher report of The Strengths and Difficulties Questionnaire (SDQ)** at age 7.5 years. The SDQ is a screening questionnaire for measuring children’s behaviour, emotions and relationships (Goodman, 1997). The hyperactivity scale consists of 5 items. The items are assessed in the 3-point Likert scale in the presence of symptoms: 0=not true; 1=somewhat true, and 2=certainly true |
| **GenR** | **Maternal report of the revised Conner’s Parent Rating Scale (CPRS-R)** at age 7.5 years. The CPRS-R was developed for screening and assessing child’s behavioural problems based on parental report and has been found to be a good instrument for distinguishing ADHD symptom domains (inattention and hyperactive/impulsive) (Conners et al., 1998). It consists of 9 items measuring hyperactivity/impulsivity and 12 items measuring cognitive problems (focusing on attention problems) which correspond to the DSM-IV criteria. The items are assessed in the 4-point Likert scale (0 for not at all true to 3 for very much true).  **Maternal report of The Child Behaviour Checklist (CBCL)** at age 6 years. The CBCL provides information on child’s behavioral problems and social competencies (Achenbach & Rescorla, 2001). Attention problems subscale consists of 5 items.  **Teacher Report Form of CBCL (CBCL-TRF)** at age 7 years. TRF consists of 26 items.  The items in CBCL and TRF are assessed in the 3-point Likert scale in the presence of symptoms: 0=not true, 1=somewhat true and 2= very true. |
| **MoBA** | **Maternal report of the Parent/Teacher Rating Scale for Disruptive Behaviour Disorders (RS-DBD)** at age 8. The RS-DBD has a parent and teacher version for measuring disruptive behaviour disorders in children which closely linked with DSM criteria. It consists of 18 items that are measuring ADHD symptoms and separately inattentive and hyperactive/impulsive symptom domains (Silva et al., 2005). The items are rated in 4-point Likert scale: 1 = not at all; 2 = just a little; 3 = pretty much and 4 = very much.  In MoBa, currently maternal report is available. |
|  | All these scales have shown good psychometric properties (Conners et al., 1998; Goodman et al., 2000; Silva et al., 2005; Goodman, 2001; Rishel et al., 2005), but SDQ and CBCL do not distinguish ADHD symptom domains. |

### Supplementary Table S3. Phenotyping in Genome-wide association studies

|  | **Smoking** | **Alcohol** | **Coffee** |
| --- | --- | --- | --- |
| **GWAS** | The GWAS and Sequencing Consortium of Alcohol and Nicotine use (GSCAN)  N=1.2 million | | The Coffee and Caffeine Genetics Consortium  N=91,462 |
|  | Smoking heaviness was defined as number of cigarettes individual smoked per day. Quantitative measure of cigarettes per day was binned to categories: 1-5; 6-15; 16-25; 26-35 and 36+ cigarettes per day. Studies with pre-defined bins were left the same. | Alcohol consumption was defined as number of drinks per week an individual consumed. As reporting of weekly alcohol consumption varied across the studies, binned response ranges were used (e.g. for 1-4 drinks per week, a midpoint was used). Phenotype was left-anchored at 1 and log-transformed before performing analysis to avoid effect from outliers | Coffee data was collected categorically. Median value of each category was taken (e.g. for 2-3 cups per day, a 2.5 cups per day) for primary phenotype (“phenotype 1”).  Additionally, high/infrequent and non-coffee consumers were compared (“phenotype 2”) |
| **SNPs** | 55 SNPs were discovered to be independently associated with smoking heaviness at the genome-wide significance level (5x10^-8^) | 99 SNPs were associated with alcohol consumption at the genome-wide significance level | 8 SNPs were independently associated with cups of coffee consumed per day at the genome-wide level of significance* |
|  |  | 2 overlapping SNPs between alcohol and coffee SNPs | |

*** *These SNPs have been also validated in caffeine consumption from other sources of caffeine besides coffee (Treur et al., 2016 & McMahon et al., 2014***)**

**Supplementary Table S4. Correlation between PRS in ALSPAC and MoBa**

|  | **Alcohol PRS** | **Smoking heaviness PRS** | **Lifetime smoking PRS** |
| --- | --- | --- | --- |
| **ALSPAC** |  |  |  |
| **Smoking heaviness PRS** | -0.005 |  |  |
| **Lifetime smoking PRS** | 0.029 | 0.209 |  |
| **Caffeine PRS** | 0.121 | -0.007 | -0.009 |
| **MoBA** |  |  |  |
| **Smoking heaviness PRS** | -0.007 |  |  |
| **Lifetime smoking PRS** | 0.043 | 0.217 |  |
| **Caffeine PRS** | 0.128 | -0.013 | 0.001 |

### Supplementary Table S5. Associations of maternal and paternal prenatal smoking on high risk of maternal reported offspring ADHD symptoms in ALSPAC

|  |  |  | **Unadjusted model** | | |  |  | **Adjusted model*** | | |  |  | **Mutually adjusted model**** | | |
| --- | --- | --- | --- | --- | --- | --- | --- | --- | --- | --- | --- | --- | --- | --- | --- |
|  | **N** | **n** | **OR** | **95% CI** | **p-value** | **N** | **n** | **OR** | **95% CI** | **p-value** | **N** | **n** | **OR** | **95% CI** | **p-value** |
| **Maternal** | | | | | | | | | | | | | | | |
| **ADHD (DAWBA)** | 7731 | 1114 |  |  | <0.001 | 6675 | 934 |  |  | 0.034 | 5201 | 709 |  |  | 0.654 |
| No cigarettes (ref) | 6256 | 822 | - | - |  | 5483 | 701 | - | - |  | 4319 | 542 | - | - |  |
| 1-4 cigarettes | 372 | 67 | 1.452 | 1.104,1.911 |  | 314 | 51 | 1.031 | 0.747,1.424 |  | 244 | 38 | 0.843 | 0.576,1.232 |  |
| 5-9 cigarettes | 367 | 68 | 1.503 | 1.144,1.976 |  | 301 | 60 | 1.264 | 0.922,1.734 |  | 228 | 44 | 1.068 | 0.734,1.553 |  |
| >10 cigarettes | 736 | 157 | 1.793 | 1.481,2.170 |  | 577 | 122 | 1.265 | 0.990,1.617 |  | 410 | 85 | 1.074 | 0.797,1.449 |  |
| **Hyperactive** | 7751 | 984 |  |  | <0.001 | 6693 | 828 |  |  | 0.157 | 5211 | 622 |  |  | 0.839 |
| No cigarettes (ref) | 6274 | 726 | - | - |  | 5499 | 628 | - | - |  | 4329 | 486 | - | - |  |
| 1-4 cigarettes | 373 | 59 | 1.436 | 1.076,1.916 |  | 315 | 43 | 0.970 | 0.689,1.368 |  | 245 | 32 | 0.837 | 0.559,1.254 |  |
| 5-9 cigarettes | 366 | 64 | 1.619 | 1.223,2.144 |  | 300 | 54 | 1.266 | 0.913,1.754 |  | 227 | 35 | 0.977 | 0.652,1.463 |  |
| >10 cigarettes | 738 | 135 | 1.711 | 1.398,2.094 |  | 579 | 103 | 1.159 | 0.896,1.501 |  | 410 | 69 | 0.982 | 0.715,1.348 |  |
| **Inattentive** | 7743 | 1058 |  |  | <0.001 | 6688 | 890 |  |  | 0.098 | 5208 | 686 |  |  | 0.651 |
| No cigarettes (ref) | 6266 | 810 | - | - |  | 5494 | 690 | - | - |  | 4326 | 541 | - | - |  |
| 1-4 cigarettes | 372 | 54 | 1.144 | 0.849,1.541 |  | 314 | 44 | 0.940 | 0.670,1.321 |  | 243 | 31 | 0.731 | 0.487,1.099 |  |
| 5-9 cigarettes | 368 | 62 | 1.365 | 1.029,1.811 |  | 302 | 52 | 1.200 | 0.863,1.670 |  | 228 | 39 | 1.063 | 0.721,1.568 |  |
| >10 cigarettes | 737 | 132 | 1.470 | 1.201,1.799 |  | 578 | 104 | 1.224 | 0.947,1.581 |  | 411 | 75 | 1.092 | 0.802,1.488 |  |
| **ADHD (SDQ)** | 7994 | 892 |  |  | <0.001 | 6946 | 737 |  |  | 0.029 | 5405 | 561 |  |  | 0.159 |
| No cigarettes (ref) | 6421 | 639 | - | - |  | 5672 | 545 | - | - |  | 4459 | 415 | - | - |  |
| 1-4 cigarettes | 386 | 57 | 1.568 | 1.170,2.101 |  | 323 | 45 | 1.261 | 0.899,1.767 |  | 249 | 35 | 1.172 | 0.792,1.732 |  |
| 5-9 cigarettes | 380 | 51 | 1.403 | 1.033,1.905 |  | 317 | 38 | 0.967 | 0.669,1.397 |  | 238 | 28 | 0.878 | 0.568,1.358 |  |
| >10 cigarettes | 807 | 145 | 1.982 | 1.627,2.414 |  | 634 | 109 | 1.367 | 1.060,1.762 |  | 459 | 83 | 1.303 | 0.963,1.764 |  |
| **Paternal** | | | | | | | | | | | | | | | |
| **ADHD (DAWBA)** | 5841 | 815 |  |  | <0.001 | 4657 | 622 |  |  | 0.001 | 4647 | 618 |  |  | 0.005 |
| No cigarettes (ref) | 3977 | 474 | - | - |  | 3288 | 382 | - | - |  | 3284 | 381 | - | - |  |
| 1-4 cigarettes | 296 | 47 | 1.395 | 1.007,1.933 |  | 237 | 35 | 1.214 | 0.825,1.787 |  | 235 | 34 | 1.184 | 0.799,1.755 |  |
| 5-9 cigarettes | 254 | 46 | 1.634 | 1.171,2.281 |  | 185 | 29 | 1.293 | 0.846,1.978 |  | 184 | 28 | 1.239 | 0.803,1.912 |  |
| >10 cigarettes | 1314 | 248 | 1.719 | 1.453,2.034 |  | 947 | 176 | 1.440 | 1.159,1.790 |  | 944 | 175 | 1.378 | 1.097,1.730 |  |
| **Hyperactive** | 5851 | 711 |  |  | <0.001 | 4666 | 535 |  |  | 0.042 | 4656 | 532 |  |  | 0.105 |
| No cigarettes (ref) | 3983 | 427 | - | - |  | 3295 | 341 | - | - |  | 3291 | 340 | - | - |  |
| 1-4 cigarettes | 297 | 38 | 1.222 | 0.857,1.743 |  | 237 | 29 | 1.157 | 0.764,1.753 |  | 235 | 28 | 1.112 | 0.728,1.700 |  |
| 5-9 cigarettes | 254 | 29 | 1.073 | 0.720,1.601 |  | 184 | 16 | 0.730 | 0.427,1.248 |  | 183 | 16 | 0.727 | 0.424,1.248 |  |
| >10 cigarettes | 1317 | 217 | 1.643 | 1.377,1.961 |  | 950 | 149 | 1.313 | 1.043,1.652 |  | 947 | 148 | 1.262 | 0.991,1.606 |  |
| **Inattentive** | 5849 | 478 |  |  | <0.001 | 4666 | 614 |  |  | 0.065 | 4656 | 610 |  |  | 0.089 |
| No cigarettes (ref) | 3979 | 45 | - | - |  | 3293 | 394 | - | - |  | 3289 | 393 | - | - |  |
| 1-4 cigarettes | 299 | 41 | 1.298 | 0.932,1.807 |  | 239 | 37 | 1.237 | 0.847,1.806 |  | 237 | 36 | 1.236 | 0.841,1.817 |  |
| 5-9 cigarettes | 254 | 215 | 1.410 | 0.996,1.996 |  | 185 | 27 | 1.149 | 0.743,1.776 |  | 184 | 26 | 1.128 | 0.723,1.760 |  |
| >10 cigarettes | 1317 | 779 | 1.429 | 1.200,1.702 |  | 949 | 156 | 1.225 | 0.981,1.530 |  | 946 | 155 | 1.219 | 0.966,1.538 |  |
| **ADHD (SDQ)** | 6030 | 646 |  |  | 0.099 | 4804 |  |  |  | 0.094 | 4793 |  |  |  | 0.023 |
| No cigarettes (ref) | 4059 | 373 | - | - |  | 3358 | 289 | - | - |  | 3353 | 289 | - | - |  |
| 1-4 cigarettes | 307 | 37 | 1.354 | 0.945,1.940 |  | 241 | 30 | 1.421 | 0.942,2.144 |  | 239 | 29 | 1.322 | 0.869,2.013 |  |
| 5-9 cigarettes | 268 | 32 | 1.340 | 0.912,1.968 |  | 196 | 23 | 1.262 | 0.795,2.004 |  | 195 | 23 | 1.215 | 0.762,1.937 |  |
| >10 cigarettes | 1396 | 204 | 1.691 | 1.409,2.030 |  | 1009 | 145 | 1.479 | 1.171,1.868 |  | 1006 | 144 | 1.326 | 1.036,1.699 |  |

Note: DAWBA – Development and Well-Being Assessment; SDQ – Strengths and Difficulties Questionnaire; N – total sample size; n – number of ADHD cases; OR – odds ratio; 95% CI – 95% confidence intervals; *adjusted for child’s gender, ethnicity, parity, parental age, marital status, education, financial difficulties, depression and anxiety symptoms, prenatal alcohol and caffeine use; **adjusted additionally for partners’ prenatal smoking

### Supplementary Table S6. Associations of maternal and paternal prenatal smoking on high risk of maternal and teacher reported offspring ADHD symptoms in GenR

|  |  |  | **Unadjusted model** | | |  |  | **Adjusted model*** | | | |  |  | **Mutually adjusted model**** | | |
| --- | --- | --- | --- | --- | --- | --- | --- | --- | --- | --- | --- | --- | --- | --- | --- | --- |
|  | **N** | **n** | **OR** | **95% CI** | **p-value** | **N** | **n** | **OR** | | **95% CI** | **p-value** | **N** | **n** | **OR** | **95% CI** | **p-value** |
| **Maternal** | | | | | | | | | | | | | | | | |
| **ADHD (CPRS-R)** | 3116 | 452 |  |  | <0.001 | 2053 | 282 |  | |  | 0.023 | 1535 | 196 |  |  | 0.018 |
| No cigarettes (ref) | 2471 | 330 | - | - |  | 1623 | 209 | - | | - |  | 1219 | 145 | - | - |  |
| 1-4 cigarettes | 358 | 62 | 1.359 | 1.009,1.829 |  | 226 | 32 | 1.016 | | 0.666,1.548 |  | 165 | 21 | 1.009 | 0.592,1.720 |  |
| 5-9 cigarettes | 159 | 29 | 1.447 | 0.952,2.200 |  | 113 | 18 | 1.132 | | 0.649,1.974 |  | 82 | 13 | 1.260 | 0.639,2.484 |  |
| >10 cigarettes | 128 | 31 | 2.073 | 1.361,3.158 |  | 91 | 23 | 1.965 | | 1.144,3.375 |  | 69 | 17 | 2.480 | 1.293,4.755 |  |
| **Hyperactive** | 3119 | 345 |  |  | 0.007 | 2057 | 201 |  | |  | 0.076 | 1536 | 143 |  |  | 0.217 |
| No cigarettes (ref) | 2475 | 260 | - | - |  | 1628 | 152 | - | | - |  | 1221 | 107 | - | - |  |
| 1-4 cigarettes | 358 | 41 | 1.102 | 0.777,1.563 |  | 226 | 20 | 0.931 | | 0.560,1.548 |  | 165 | 14 | 0.771 | 0.413,1.440 |  |
| 5-9 cigarettes | 158 | 20 | 1.235 | 0.759,1.563 |  | 112 | 12 | 1.046 | | 0.544,2.014 |  | 81 | 9 | 0.972 | 0.445,2.123 |  |
| >10 cigarettes | 128 | 24 | 1.966 | 1.239,3.121 |  | 91 | 17 | 1.988 | | 1.079,3.663 |  | 69 | 13 | 1.988 | 0.964,4.099 |  |
| **Inattentive** | 3117 | 391 |  |  | 0.039 | 2058 | 240 |  | |  | 0.288 | 1537 | 170 |  |  | 0.143 |
| No cigarettes (ref) | 2472 | 298 | - | - |  | 1628 | 188 | - | | - |  | 1221 | 133 | - | - |  |
| 1-4 cigarettes | 358 | 49 | 1.157 | 0.836,1.601 |  | 226 | 21 | 0.724 | | 0.444,1.183 |  | 165 | 15 | 0.806 | 0.442,1.470 |  |
| 5-9 cigarettes | 159 | 19 | 0.990 | 0.604,1.623 |  | 113 | 10 | 0.646 | | 0.323,1.293 |  | 82 | 7 | 0.681 | 0.292,1.589 |  |
| >10 cigarettes | 128 | 25 | 1.771 | 1.125,2.787 |  | 91 | 21 | 2.013 | | 1.154,3.513 |  | 69 | 15 | 2.492 | 1.259,4.932 |  |
| **ADHD (CBCL)** | 4168 | 595 |  |  | <0.001 | 2565 | 331 |  | |  | 0.004 | 1835 | 211 |  |  | 0.031 |
| No cigarettes (ref) | 3199 | 396 | - | - |  | 1970 | 217 | - | | - |  | 1432 | 140 | - | - |  |
| 1-4 cigarettes | 521 | 99 | 1.661 | 1.303,2.117 |  | 306 | 54 | 1.474 | | 1.040,2.090 |  | 211 | 31 | 1.223 | 0.770,1.942 |  |
| 5-9 cigarettes | 255 | 60 | 2.178 | 1.601,2.963 |  | 161 | 31 | 1.353 | | 0.858,2.132 |  | 107 | 21 | 1.533 | 0.863,2.725 |  |
| >10 cigarettes | 193 | 40 | 1.851 | 1.286,2.662 |  | 128 | 29 | 1.758 | | 1.086,2.846 |  | 85 | 19 | 1.797 | 0.972,3.323 |  |
| **ADHD (TRF)** | 3023 |  |  |  | <0.001 | 1671 | 218 |  | |  | 0.002 | 1148 | 125 |  |  | 0.059 |
| No cigarettes (ref) | 2247 | 307 | - | - |  | 1253 | 142 | - | | - |  | 878 | 85 | - | - |  |
| 1-4 cigarettes | 420 | 76 | 1.396 | 1.059,1840 |  | 221 | 33 | 1.249 | | 0.802,1.944 |  | 141 | 16 | 1.005 | 0.529,1.908 |  |
| 5-9 cigarettes | 207 | 52 | 2.120 | 1.514,2.968 |  | 117 | 25 | 2.238 | | 1.302,3.849 |  | 75 | 12 | 1.594 | 0.737,3.449 |  |
| >10 cigarettes | 149 | 27 | 1.399 | 0.906,2.158 |  | 80 | 18 | 2.034 | | 1.087,3.806 |  | 54 | 12 | 2.155 | 0.944,4.918 |  |
| **Paternal** | | | | | | | | | | | | | | | | |
| **ADHD (CPRS-R)** | 2381 | 333 |  |  | 0.882 | 2117 | 289 | |  |  | 0.220 | 1930 | 260 |  |  | 0.237 |
| No cigarettes (ref) | 1412 | 197 | - | - |  | 1249 | 170 | | - | - |  | 1148 | 149 | - | - |  |
| 1-4 cigarettes | 387 | 52 | 0.957 | 0.689,1.330 |  | 355 | 46 | | 0.845 | 0.588,1.214 |  | 328 | 43 | 0.891 | 0.606,1.310 |  |
| 5-9 cigarettes  >10 cigarettes | 166  416 | 26  58 | 1.145  0.999 | 0.734,1.787  0.729,1.370 |  | 148  365 | 21  52 | | 0.797  0.822 | 0.476,1.334  0.574,1.176 |  | 132  322 | 20  48 | 0.873  0.785 | 0.502,1.518  0.522,1.180 |  |
| **Hyperactive** | 2381 | 241 |  |  | 0.031 | 2117 | 202 | |  |  | 0.193 | 1930 | 186 |  |  | 0.448 |
| No cigarettes (ref) | 1412 | 130 | - | - |  | 1249 | 104 | | - | - |  | 1148 | 98 | - | - |  |
| 1-4 cigarettes | 386 | 39 | 1.108 | 0.760,1.616 |  | 354 | 35 | | 1.116 | 0.735,1.696 |  | 327 | 31 | 1.088 | 0.695,1.704 |  |
| 5-9 cigarettes  >10 cigarettes | 166  417 | 19  53 | 1.275  1.436 | 0.765,2.124  1.022,2.017 |  | 148  366 | 15  48 | | 0.965  1.334 | 0.527,1.766  0.901,1.974 |  | 132  323 | 15  42 | 1.122  1.184 | 0.598,2.106  0.757,1.852 |  |
| **Inattentive** | 2382 | 290 |  |  | 0.661 | 2119 | 256 | |  |  | 0.024 | 1931 | 233 |  |  | 0.078 |
| No cigarettes (ref) | 1412 | 174 | - | - |  | 1250 | 156 | | - | - |  | 1149 | 139 | - | - |  |
| 1-4 cigarettes | 386 | 47 | 0.986 | 0.699,1.391 |  | 354 | 42 | | 0.864 | 0.593,1.259 |  | 327 | 40 | 0.942 | 0.633,1.401 |  |
| 5-9 cigarettes  >10 cigarettes | 166  418 | 22  47 | 1.087  0.901 | 0.675,1.749  0.640,1.269 |  | 148  367 | 17  41 | | 0.709  0.655 | 0.406,1.240  0.443,0.969 |  | 132  323 | 17  37 | 0.906  0.658 | 0.506,1.621  0.423,1.024 |  |
| **ADHD (CBCL)** | 2913 | 366 |  |  | <0.001 | 2541 | 305 | |  |  | 0.025 | 2323 | 275 |  |  | 0.039 |
| No cigarettes (ref) | 1689 | 177 | - | - |  | 1478 | 148 | | - | - |  | 1361 | 130 | - | - |  |
| 1-4 cigarettes | 458 | 61 | 1.313 | 0.961,1.792 |  | 414 | 54 | | 1.290 | 0.911,1.826 |  | 381 | 49 | 1.324 | 0.914,1.919 |  |
| 5-9 cigarettes  >10 cigarettes | 220  546 | 33  95 | 1.507  1.799 | 1.009,2.252  1.373,2.358 |  | 183  466 | 26  77 | | 1.198  1.448 | 0.747,1.921  1.051,1.994 |  | 164  417 | 25  71 | 1.331  1.455 | 0.807,2.195  1.011,2.093 |  |
| **ADHD (TRF)** | 1876 | 214 |  |  | 0.005 | 1570 | 166 | |  |  | 0.230 | 1445 | 151 |  |  | 0.347 |
| No cigarettes (ref) | 1045 | 100 | - | - |  | 883 | 77 | | - | - |  | 816 | 68 | - | - |  |
| 1-4 cigarettes | 306 | 36 | 1.260 | 0.841,1.888 |  | 264 | 30 | | 1.229 | 0.758,1.992 |  | 242 | 29 | 1.413 | 0.851,2.344 |  |
| 5-9 cigarettes  >10 cigarettes | 157  368 | 27  51 | 1.963  1.520 | 1.235,3.118  1.060,2.180 |  | 121  302 | 18  41 | | 1.275  1.291 | 0.703,2.314  0.827,2.015 |  | 109  278 | 17  37 | 1.523  1.211 | 0.806,2.876  0.732,2.002 |  |

Note: CPRS-R – The revised Conners’ Parent Rating Scale; CBCL – Child Behaviour Checklist; TRF – Teacher’s Report Form; N – sample size; n – number of cases; OR – odds ratio; 95% CI – 95% confidence intervals; *adjusted for child’s gender, parity, parental ethnicity, age, education, anxiety and depression problems, financial difficulties, parental alcohol use and prenatal caffeine use in the maternal model; **additionally adjusted for partner’s smoking

### Supplementary Table S7. Associations of maternal and paternal prenatal smoking on high risk of maternal reported offspring ADHD symptoms in MoBa

|  |  |  | **Unadjusted model** | | |  | |  | **Adjusted model*** | | | |  | |  | **Mutually adjusted model**** | | |
| --- | --- | --- | --- | --- | --- | --- | --- | --- | --- | --- | --- | --- | --- | --- | --- | --- | --- | --- |
|  | **N** | **n** | **OR** | **95% CI** | **p-value** | | **N** | **n** | **OR** | **95% CI** | **p-value** | | **N** | | **n** | **OR** | **95% CI** | **p-value** |
| **Maternal** | | | | | | | | | | | | | | | | | | |
| **ADHD (RS-DBD)** | 41,515 | 5509 |  |  | <0.001 | 34,297 | | 4415 |  |  | <0.001 | | 28,055 | | 3543 |  |  | 0.006 |
| No cigarettes (ref) | 39,162 | 4993 | - | - |  | 32,487 | | 4035 | - | - |  | | 26,706 | | 3280 | - | - |  |
| 1-4 cigarettes | 1260 | 234 | 1.561 | 1.350,1.805 |  | 951 | | 168 | 1.134 | 0.946,1.360 |  | | 707 | | 111 | 0.948 | 0.761,1.182 |  |
| 5-9 cigarettes | 650 | 158 | 2.198 | 1.830,2.640 |  | 501 | | 121 | 1.659 | 1.326,2.075 |  | | 383 | | 87 | 1.485 | 1.136,1.941 |  |
| >10 cigarettes | 443 | 124 | 2.660 | 2.159,3.277 |  | 358 | | 91 | 1.495 | 1.149,1.945 |  | | 259 | | 65 | 1.376 | 1.000,1.893 |  |
| **Hyperactive** | 41,508 | 5436 |  |  | <0.001 | 34,290 | | 4353 |  |  | <0.001 | | 28,051 | | 3507 |  |  | <0.001 |
| No cigarettes (ref) | 39,158 | 4916 | - | - |  | 32,483 | | 3971 | - | - |  | | 26,704 | | 3232 | - | - |  |
| 1-4 cigarettes | 1259 | 239 | 1.632 | 1.414,1.883 |  | 950 | | 166 | 1.158 | 0.968,1.384 |  | | 707 | | 117 | 1.039 | 0.840,1.286 |  |
| 5-9 cigarettes | 649 | 159 | 2.260 | 1.884,2.712 |  | 500 | | 124 | 1.754 | 1.409,2.185 |  | | 382 | | 91 | 1.595 | 1.228,2.073 |  |
| >10 cigarettes | 442 | 122 | 2.656 | 2.152,3.278 |  | 357 | | 92 | 1.569 | 1.208,2.036 |  | | 258 | | 67 | 1.454 | 1.061,1.991 |  |
| **Inattentive** | 41,524 | 4824 |  |  | <0.001 | 34,302 | | 3874 |  |  | 0.001 | | 28,058 | | 3102 |  |  | 0.041 |
| No cigarettes (ref) | 39,170 | 4402 | - | - |  | 32,491 | | 3551 | - | - |  | | 26,708 | | 2877 | - | - |  |
| 1-4 cigarettes | 1261 | 198 | 1.471 | 1.259,1.719 |  | 952 | | 151 | 1.136 | 0.940,1.373 |  | | 708 | | 101 | 0.987 | 0.783,1.243 |  |
| 5-9 cigarettes | 650 | 131 | 1.994 | 1.640,2.423 |  | 501 | | 102 | 1.529 | 1.208,1.935 |  | | 383 | | 70 | 1.324 | 0.996,1.760 |  |
| >10 cigarettes | 443 | 93 | 2.099 | 1.663,2.648 |  | 358 | | 70 | 1.239 | 0.924,1.661 |  | | 259 | | 54 | 1.306 | 0.929,1.837 |  |
| **Paternal** | | | | | | | | | | | | | | | | | | |
| **ADHD (RS-DBD)** | 33,955 | 4397 |  |  | <0.001 | 10,804 | | 1462 |  |  | | 0.155 | 10,738 | 1454 | |  |  | 0.417 |
| No cigarettes (ref) | 26,918 | 3312 | - | - |  | 9018 | | 1198 | - | - | |  | 8965 | 1193 | | - | - |  |
| 1-4 cigarettes | 3252 | 436 | 1.104 | 0.991,1.229 |  | 1001 | | 122 | 0.859 | 0.702,1.050 | |  | 998 | 120 | | 0.833 | 0.680,1.021 |  |
| 5-9 cigarettes | 870 | 140 | 1.367 | 1.134,1.648 |  | 213 | | 41 | 1.398 | 0.980,1.994 | |  | 211 | 40 | | 1.317 | 0.919,1.889 |  |
| >10 cigarettes | 2915 | 509 | 1.508 | 1.360,1.672 |  | 572 | | 101 | 1.166 | 0.917,1.484 | |  | 564 | 101 | | 1.090 | 0.846,1.404 |  |
| **Hyperactive** | 33,951 | 4370 |  |  | <0.001 | 10,802 | | 1432 |  |  | | 0.082 | 10,736 | 1424 | |  |  | 0.242 |
| No cigarettes (ref) | 26,917 | 3284 | - | - |  | 9017 | | 1162 | - | - | |  | 8964 | 1157 | | - | - |  |
| 1-4 cigarettes | 3252 | 434 | 1.108 | 0.995,1.234 |  | 1000 | | 134 | 1.000 | 0.822,1.217 | |  | 997 | 132 | | 0.976 | 0.801,1.190 |  |
| 5-9 cigarettes | 870 | 133 | 1.299 | 1.073,1.572 |  | 213 | | 40 | 1.412 | 0.988,2.020 | |  | 211 | 39 | | 1.351 | 0.939,1.945 |  |
| >10 cigarettes | 2912 | 519 | 1.561 | 1.408,1.730 |  | 572 | | 96 | 1.142 | 0.893,1.459 | |  | 564 | 96 | | 1.091 | 0.843,1.411 |  |
| **Inattentive** | 33,958 | 3851 |  |  | <0.001 | 10,806 | | 1254 |  |  | | 0.133 | 10,740 | 1248 | |  |  | 0.295 |
| No cigarettes (ref) | 26,923 | 2922 | - | - |  | 9019 | | 1023 | - | - | |  | 8966 | 1018 | | - | - |  |
| 1-4 cigarettes | 3251 | 383 | 1.097 | 0.979,1.229 |  | 1001 | | 111 | 0.915 | 0.741,1.131 | |  | 998 | 110 | | 0.901 | 0.727,1.115 |  |
| 5-9 cigarettes | 869 | 122 | 1.341 | 1.102,1.633 |  | 213 | | 31 | 1.178 | 0.794,1.746 | |  | 211 | 31 | | 1.163 | 0.784,1.725 |  |
| >10 cigarettes | 2915 | 424 | 1.398 | 1.253,1.560 |  | 573 | | 89 | 1.209 | 0.937,1.560 | |  | 565 | 89 | | 1.157 | 0.887,1.509 |  |

Note: RS-DBD – the Disruptive Behaviour Disorders scale; N – sample size; n – number of cases; OR – odds ratio; 95% CI – 95% confidence interval;. *adjusted for child’s gender, birth year, parity, parental age, education, marital status, financial difficulties, depression and anxiety symptoms, prenatal alcohol and caffeine consumption; **additionally adjusted for partner’s smoking

### Supplementary Table S8. Associations of maternal prenatal smoking on high risk of maternal reported offspring ADHD symptoms in MoBa (additionally adjusted for maternal ADHD)

|  |  |  | **Unadjusted model** | | |  |  | **Adjusted model*** | | |  |  | **Mutually adjusted model**** | | |
| --- | --- | --- | --- | --- | --- | --- | --- | --- | --- | --- | --- | --- | --- | --- | --- |
|  | **N** | **n** | **OR** | **95% CI** | **p-value** | **N** | **n** | **OR** | **95% CI** | **p-value** | **N** | **n** | **OR** | **95% CI** | **p-value** |
| **ADHD (RS-DBD)** | 41,515 | 5509 |  |  | <0.001 | 28,507 | 3655 |  |  | <0.001 | 23,491 | 2954 |  |  | 0.026 |
| No cigarettes (ref) | 39,162 | 4993 | - | - |  | 27,093 | 3360 | - | - |  | 22,430 | 2748 | - | - |  |
| 1-4 cigarettes | 1260 | 234 | 1.561 | 1.350,1.805 |  | 764 | 138 | 1.185 | 0.969,1.450 |  | 572 | 94 | 1.035 | 0.813,1.318 |  |
| 5-9 cigarettes | 650 | 158 | 2.198 | 1.830,2.640 |  | 371 | 91 | 1.639 | 1.266,2.121 |  | 285 | 64 | 1.461 | 1.071,1.994 |  |
| >10 cigarettes | 443 | 124 | 2.660 | 2.159,3.277 |  | 279 | 66 | 1.315 | 0.961,1.799 |  | 204 | 48 | 1.276 | 0.875,1.861 |  |
| **Hyperactive** | 41,508 | 5436 |  |  | <0.001 | 28,504 | 3600 |  |  | <0.001 | 23,489 | 2933 |  |  | 0.010 |
| No cigarettes (ref) | 39,158 | 4916 | - | - |  | 27,092 | 3308 | - | - |  | 22,429 | 2719 | - | - |  |
| 1-4 cigarettes | 1259 | 239 | 1.632 | 1.414,1.883 |  | 763 | 136 | 1.188 | 0.974,1.448 |  | 572 | 99 | 1.103 | 0.873,1.394 |  |
| 5-9 cigarettes | 649 | 159 | 2.260 | 1.884,2.712 |  | 371 | 90 | 1.626 | 1.263,2.094 |  | 285 | 64 | 1.430 | 1.055,1.939 |  |
| >10 cigarettes | 442 | 122 | 2.656 | 2.152,3.278 |  | 278 | 66 | 1.343 | 0.982,1.836 |  | 203 | 51 | 1.359 | 0.938,1.970 |  |
| **Inattentive** | 41,524 | 4824 |  |  | <0.001 | 28,512 | 3186 |  |  | 0.030 | 23,494 | 2571 |  |  | 0.195 |
| No cigarettes (ref) | 39,170 | 4402 | - | - |  | 27,098 | 2940 | - | - |  | 22,433 | 2398 | - | - |  |
| 1-4 cigarettes | 1261 | 198 | 1.471 | 1.259,1.719 |  | 764 | 119 | 1.144 | 0.924,1.416 |  | 572 | 81 | 1.004 | 0.776,1.299 |  |
| 5-9 cigarettes | 650 | 131 | 1.994 | 1.640,2.423 |  | 371 | 80 | 1.614 | 1.234,2.112 |  | 285 | 55 | 1.406 | 1.015,1.947 |  |
| >10 cigarettes | 443 | 93 | 2.099 | 1.663,2.648 |  | 279 | 47 | 0.998 | 0.698,1.427 |  | 204 | 37 | 1.077 | 0.711,1.631 |  |

Note: RS-DBD – the Disruptive Behaviour Disorders scale; N – sample size; n – number of cases; OR – odds ratio; 95% CI – 95% confidence intervals; *adjusted for child’s gender, birth year, parity, maternal age, education, marital status, financial difficulties, depression, anxiety and ADHD symptoms, prenatal alcohol and caffeine consumption; **additionally adjusted for partner’s smoking

### Supplementary Table S9. Associations of maternal smoking before pregnancy on high risk of maternal reported offspring ADHD symptoms in MoBa

|  |  |  | **Unadjusted model** | | |  |  | **Adjusted model*** | | |  |  | **Mutually adjusted model**** | | |
| --- | --- | --- | --- | --- | --- | --- | --- | --- | --- | --- | --- | --- | --- | --- | --- |
|  | **N** | **n** | **OR** | **95% CI** | **p-value** | **N** | **n** | **OR** | **95% CI** | **p-value** | **N** | **n** | **OR** | **95% CI** | **p-value** |
| **ADHD (RS-DBD)** | 41,263 | 5471 |  |  | <0.001 | 36,636 | 4750 |  |  | <0.001 | 30,013 | 3807 |  |  | 0.051 |
| No cigarettes (ref) | 31,691 | 3880 | - | - |  | 28,375 | 3417 | - | - |  | 23,448 | 2788 | - | - |  |
| 1-4 cigarettes | 4364 | 574 | 1.086 | 0.988,1.193 |  | 3837 | 497 | 0.968 | 0.871,1.075 |  | 3117 | 395 | 0.935 | 0.830,1.054 |  |
| 5-9 cigarettes | 1619 | 258 | 1.359 | 1.184,1.560 |  | 1395 | 217 | 1.085 | 0.927,1.269 |  | 1095 | 161 | 0.986 | 0.819,1.187 |  |
| >10 cigarettes | 3589 | 759 | 1.922 | 1.762,2.098 |  | 3029 | 619 | 1.319 | 1.184,1.470 |  | 2353 | 463 | 1.182 | 1.036,1.350 |  |
| **Hyperactive** | 41,254 | 5390 |  |  | <0.001 | 36,626 | 4693 |  |  | <0.001 | 30,007 | 3792 |  |  | <0.001 |
| No cigarettes (ref) | 31,686 | 3787 | - | - |  | 28,369 | 3343 | - | - |  | 23,445 | 2730 | - | -- |  |
| 1-4 cigarettes | 4364 | 597 | 1.168 | 1.064,1.282 |  | 3837 | 523 | 1.063 | 0.958,1.179 |  | 3117 | 427 | 1.070 | 0.952,1.203 |  |
| 5-9 cigarettes | 1617 | 261 | 1.418 | 1.236,1.627 |  | 1393 | 213 | 1.101 | 0.941,1.289 |  | 1094 | 164 | 1.057 | 0.879,1.271 |  |
| >10 cigarettes | 3587 | 745 | 1.931 | 1.769,2.109 |  | 3027 | 614 | 1.366 | 1.227,1.521 |  | 2351 | 471 | 1.279 | 1.122,1.457 |  |
| **Inattentive** | 41,271 | 4791 |  |  | <0.001 | 36,642 | 4169 |  |  | 0.002 | 30,015 | 3331 |  |  | 0.112 |
| No cigarettes (ref) | 31,694 | 3432 | - | - |  | 28,375 | 3022 | - | - |  | 23,446 | 2464 | - | - |  |
| 1-4 cigarettes | 4364 | 485 | 1.030 | 0.930,1.140 |  | 3838 | 422 | 0.922 | 0.824,1.032 |  | 3118 | 328 | 0.885 | 0.778,1.007 |  |
| 5-9 cigarettes | 1620 | 230 | 1.363 | 1.181,1.573 |  | 1396 | 193 | 1.081 | 0.917,1.274 |  | 1095 | 141 | 1.002 | 0.825,1.218 |  |
| >10 cigarettes | 3593 | 644 | 1.798 | 1.638,1.974 |  | 3033 | 532 | 1.236 | 1.101,1.387 |  | 2356 | 398 | 1.165 | 1.012,1.341 |  |

Note: RS-DBD – the Disruptive Behaviour Disorders scale; N – sample size; n – number of cases; OR – odds ratio; 95% CI – 95% confidence intervals; *adjusted for child’s gender, birth year, parity, maternal age, education, marital status, financial difficulties, depression and anxiety symptoms, alcohol and caffeine consumption before pregnancy; **additionally adjusted for partner’s smoking before partner’s pregnancy

### Supplementary Table S10. Associations of maternal and paternal prenatal smoking on high risk of teacher reported offspring ADHD symptoms in ALSPAC

|  |  | |  | | **Unadjusted model** | | | | |  | |  | | **Adjusted model*** | | | | |  | |  | **Mutually adjusted model**** | | | |
| --- | --- | --- | --- | --- | --- | --- | --- | --- | --- | --- | --- | --- | --- | --- | --- | --- | --- | --- | --- | --- | --- | --- | --- | --- | --- |
|  | **N** | | **n** | | **OR** | **95% CI** | | **p-value** | | **N** | | **n** | | **OR** | | **95% CI** | | **p-value** | **N** | | **n** | **OR** | | **95% CI** | **p-value** |
| **Maternal** | | | | | | | | | | | | | | | | | | | | | | | | | |
| **ADHD (DAWBA)** | 5767 | | 808 | |  |  | | <0.001 | | 4584 | | 576 | |  | |  | | 0.087 | 3452 | | 396 |  | |  | 0.331 |
| No cigarettes (ref) | 4433 | | 530 | | - | - | |  | | 3648 | | 404 | | - | | - | |  | 2792 | | 286 | - | | - |  |
| 1-4 cigarettes | 296 | | 63 | | 1.991 | 1.486,2.668 | |  | | 226 | | 46 | | 1.668 | | 1.157,2.404 | |  | 164 | | 30 | 1.631 | | 1.041,2.554 |  |
| 5-9 cigarettes | 314 | | 47 | | 1.296 | 0.938,1.791 | |  | | 230 | | 31 | | 0.921 | | 0.604,1.405 | |  | 163 | | 20 | 0.819 | | 0.481,1.396 |  |
| >10 cigarettes | 724 | | 168 | | 2.225 | 1.831,2.704 | |  | | 480 | | 95 | | 1.317 | | 0.984,1.762 | |  | 333 | | 60 | 1.240 | | 0.863,1.782 |  |
| **Hyperactive** | 5766 | | 726 | |  |  | | <0.001 | | 4584 | | 520 | |  | |  | | 0.253 | 3452 | | 357 |  | |  | 0.419 |
| No cigarettes (ref) | 4432 | | 489 | | - | - | |  | | 3648 | | 372 | | - | | - | |  | 2792 | | 263 | - | | - |  |
| 1-4 cigarettes | 296 | | 57 | | 1.923 | 1.419,2.606 | |  | | 226 | | 42 | | 1.682 | | 1.155,2.451 | |  | 164 | | 28 | 1.761 | | 1.110,2.795 |  |
| 5-9 cigarettes | 314 | | 44 | | 1.314 | 0.943,1.832 | |  | | 230 | | 32 | | 1.169 | | 0.769,1.777 | |  | 163 | | 22 | 1.256 | | 0.748,2.107 |  |
| >10 cigarettes | 724 | | 136 | | 1.865 | 1.514,2.298 | |  | | 480 | | 74 | | 1.145 | | 0.836,1.567 | |  | 333 | | 44 | 1.099 | | 0.736,1.639 |  |
| **Inattentive** | 5767 | | 713 | |  |  | | <0.001 | | 4583 | | 508 | |  | |  | | 0.503 | 3450 | | 358 |  | |  | 0.822 |
| No cigarettes (ref) | 4433 | | 476 | | - | - | |  | | 3648 | | 365 | | - | | - | |  | 2791 | | 266 | - | | - |  |
| 1-4 cigarettes | 297 | | 53 | | 1.806 | 1.322,2.467 | |  | | 226 | | 41 | | 1.599 | | 1.094,2.337 | |  | 164 | | 26 | 1.398 | | 0.875,2.236 |  |
| 5-9 cigarettes | 313 | | 39 | | 1.183 | 0.835,1.677 | |  | | 229 | | 23 | | 0.727 | | 0.454,1.165 | |  | 162 | | 14 | 0.590 | | 0.322,1.080 |  |
| >10 cigarettes | 724 | | 145 | | 2.082 | 1.695,2.556 | |  | | 480 | | 79 | | 1.169 | | 0.859,1.591 | |  | 333 | | 52 | 1.134 | | 0.777,1.656 |  |
| **ADHD (SDQ)** | 5764 | | 633 | |  |  | | <0.001 | | 4587 | | 444 | |  | |  | | 0.025 | 3455 | |  |  | |  | 0.102 |
| No cigarettes (ref) | 4430 | | 405 | | - | - | |  | | 3650 | | 303 | | - | | - | |  | 2795 | | 217 | - | | - |  |
| 1-4 cigarettes | 296 | | 46 | | 1.829 | 1.314,2.545 | |  | | 226 | | 34 | | 1.557 | | 1.034,2.345 | |  | 164 | | 24 | 1.645 | | 1.009,2.682 |  |
| 5-9 cigarettes | 314 | | 40 | | 1.451 | 1.025,2.053 | |  | | 230 | | 27 | | 1.077 | | 0.687,1.689 | |  | 162 | | 21 | 1.285 | | 0.756,2.185 |  |
| >10 cigarettes | 724 | | 142 | | 2.425 | 1.966,2.991 | |  | | 481 | | 80 | | 1.445 | | 1.055,1.981 | |  | 334 | | 49 | 1.339 | | 0.903,1.985 |  |
| **Paternal** | | | | | | | | | | | | | | | | | | | | | | | | | |
| **ADHD (DAWBA)** | | 4081 | | 509 |  | |  | | <0.001 | | 3075 | | 334 |  |  | | 0.173 | | | 3067 | 332 | |  |  | 0.454 |
| No cigarettes (ref) | | 2648 | | 287 | - | | - | |  | | 2100 | | 200 | - | - | |  | | | 2097 | 200 | | - | - |  |
| 1-4 cigarettes | | 218 | | 26 | 1.114 | | 0.726,1.708 | |  | | 162 | | 18 | 1.074 | 0.633,1.822 | |  | | | 161 | 18 | | 1.037 | 0.608,1.769 |  |
| 5-9 cigarettes | | 186 | | 24 | 1.219 | | 0.780,1.903 | |  | | 132 | | 15 | 0.979 | 0.547,1.753 | |  | | | 132 | 15 | | 0.964 | 0.535,1.734 |  |
| >10 cigarettes | | 1029 | | 172 | 1.651 | | 1.346,2.026 | |  | | 681 | | 101 | 1.237 | 0.926,1.653 | |  | | | 677 | 99 | | 1.135 | 0.834,1.545 |  |
| **Hyperactive** | | 4081 | | 462 |  | |  | | 0.004 | | 3075 | | 313 |  |  | | 0.802 | | | 3067 | 310 | |  |  | 0.413 |
| No cigarettes (ref) | | 2648 | | 276 | - | | - | |  | | 2100 | | 203 | - | - | |  | | | 2097 | 202 | | - | - |  |
| 1-4 cigarettes | | 218 | | 24 | 1.063 | | 0.683,1.654 | |  | | 162 | | 17 | 1.033 | 0.601,1.777 | |  | | | 161 | 17 | | 0.992 | 0.573,1.716 |  |
| 5-9 cigarettes | | 186 | | 18 | 0.921 | | 0.557,1.521 | |  | | 132 | | 11 | 0.697 | 0.361,1.344 | |  | | | 132 | 11 | | 0.669 | 0.346,1.296 |  |
| >10 cigarettes | | 1029 | | 144 | 1.398 | | 1.127,1.735 | |  | | 681 | | 82 | 0.991 | 0.727,1.349 | |  | | | 677 | 80 | | 0.898 | 0.646,1.248 |  |
| **Inattentive** | | 4079 | | 456 |  | |  | | <0.001 | | 3073 | | 307 |  |  | | 0.196 | | | 3065 | 306 | |  |  | 0.215 |
| No cigarettes (ref) | | 2647 | | 250 | - | | - | |  | | 2099 | | 178 | - | - | |  | | | 2096 | 178 | | - | - |  |
| 1-4 cigarettes | | 218 | | 31 | 1.589 | | 1.064,2.375 | |  | | 162 | | 23 | 1.630 | 1.005,2.643 | |  | | | 161 | 23 | | 1.628 | 0.999,2.652 |  |
| 5-9 cigarettes | | 186 | | 20 | 1.155 | | 0.713,1.870 | |  | | 132 | | 13 | 0.936 | 0.507,1.729 | |  | | | 132 | 13 | | 0.964 | 0.519,1.790 |  |
| >10 cigarettes | | 1028 | | 155 | 1.702 | | 1.373,2.110 | |  | | 680 | | 93 | 1.233 | 0.914,1.664 | |  | | | 676 | 92 | | 1.234 | 0.900,1.694 |  |
| **ADHD (SDQ)** | | 4079 | | 401 |  | |  | | <0.001 | | 3079 | |  |  |  | | 0.124 | | | 3071 | 258 | |  |  | 0.782 |
| No cigarettes (ref) | | 2645 | | 219 | - | | - | |  | | 2101 | | 151 | - | - | |  | | | 2098 | 151 | | - | - |  |
| 1-4 cigarettes | | 218 | | 25 | 1.435 | | 0.925,2.226 | |  | | 162 | | 18 | 1.449 | 0.847,2.478 | |  | | | 161 | 18 | | 1.385 | 0.805,2.384 |  |
| 5-9 cigarettes | | 186 | | 16 | 1.043 | | 0.613,1.773 | |  | | 132 | | 9 | 0.759 | 0.369,1.559 | |  | | | 132 | 9 | | 0.694 | 0.334,1.440 |  |
| >10 cigarettes | | 1030 | | 141 | 1.757 | | 1.403,2.201 | |  | | 684 | | 83 | 1.321 | 0.961,1.817 | |  | | | 680 | 80 | | 1.079 | 0.766,1.520 |  |

Note: DAWBA – Development and Well-Being Assessment; SDQ – Strengths and Difficulties Questionnaire; N – total sample size; n – number of ADHD cases; OR – odds ratio; 95% CI – 95% confidence intervals; *adjusted for child’s gender, ethnicity, parity, parental age, marital status, education, financial difficulties, depression and anxiety symptoms, prenatal alcohol and caffeine use; **additionally adjusted for partners’ prenatal smoking

### Supplementary Table S11. Associations of maternal smoking before pregnancy on high risk of maternal and teacher reported offspring ADHD symptoms in GenR

|  |  |  | **Unadjusted model** | | |  |  | **Adjusted model*** | | |  |  | **Mutually adjusted model**** | | |
| --- | --- | --- | --- | --- | --- | --- | --- | --- | --- | --- | --- | --- | --- | --- | --- |
|  | **N** | **n** | **OR** | **95% CI** | **p-value** | **N** | **n** | **OR** | **95% CI** | **p-value** | **N** | **n** | **OR** | **95% CI** | **p-value** |
| **ADHD (CPRS-R)** | 3028 | 444 |  |  | 0.001 | 1979 | 277 |  |  | 0.100 | 1481 | 194 |  |  | 0.080 |
| No cigarettes (ref) | 1883 | 248 | - | - |  | 1226 | 159 | - | - |  | 928 | 110 | - | - |  |
| 1-4 cigarettes | 427 | 66 | 1.205 | 0.898,1.618 |  | 269 | 37 | 1.017 | 0.681,1.518 |  | 203 | 29 | 1.178 | 0.737,1.881 |  |
| 5-9 cigarettes  >10 cigarettes | 250  468 | 40  90 | 1.256  1.570 | 0.873,1.806  1.203,2.048 |  | 161  323 | 22  59 | 0.953  1.409 | 0.575,1.578  0.977,2.031 |  | 112  238 | 14  41 | 0.935  1.595 | 0.489,1.787  1.007,2.527 |  |
| **Hyperactive** | 3031 | 336 |  |  | 0.004 | 1983 | 194 |  |  | 0.059 | 1482 | 138 |  |  | 0.111 |
| No cigarettes (ref) | 1888 | 187 | - | - |  | 1231 | 105 | - | - |  | 930 | 72 | - | - |  |
| 1-4 cigarettes | 426 | 53 | 1.292 | 0.934,1.789 |  | 269 | 33 | 1.533 | 0.993,2.366 |  | 203 | 24 | 1.444 | 0.858,2.431 |  |
| 5-9 cigarettes  >10 cigarettes | 250  467 | 26  70 | 1.056  1.604 | 0.685,1.628  1.193,2.155 |  | 161  322 | 15  41 | 1.041  1.480 | 0.575,1.883  0.964,2.272 |  | 112  237 | 10  32 | 0.931  1.619 | 0.442,1.961  0.958,2.738 |  |
| **Inattentive** | 3030 | 380 |  |  | 0.079 | 1985 | 231 |  |  | 0.715 | 1484 | 165 |  |  | 0.192 |
| No cigarettes (ref) | 1887 | 224 | - | - |  | 1232 | 142 | - | - |  | 931 | 99 | - | - |  |
| 1-4 cigarettes | 426 | 54 | 1.078 | 0.784,1.481 |  | 269 | 27 | 0.815 | 0.521,1.275 |  | 203 | 21 | 0.932 | 0.553,1.569 |  |
| 5-9 cigarettes  >10 cigarettes | 250  467 | 32  70 | 1.090  1.309 | 0.733,1.620  0.980,1.749 |  | 161  323 | 17  45 | 0.840  1.148 | 0.483,1.460  0.771,1.711 |  | 112  238 | 12  33 | 0.974  1.459 | 0.493,1.923  0.892,2.387 |  |
| **ADHD (CBCL)** | 4076 | 596 |  |  | <0.001 | 2484 | 332 |  |  | 0.006 | 1778 | 214 |  |  | 0.083 |
| No cigarettes (ref) | 2477 | 315 | - | - |  | 1497 | 167 | - | - |  | 1090 | 111 | - | - |  |
| 1-4 cigarettes  5-9 cigarettes  >10 cigarettes | 570  357  672 | 75  70  136 | 1.040  1.674  1.741 | 0.794,1.362  1.257,2.230  1.394,2.175 |  | 341  212  434 | 43  39  83 | 1.088  1.583  1.454 | 0.749,1.581  1.056,2.372  1.051,2.013 |  | 240  144  304 | 25  25  53 | 0.926  1.455  1.396 | 0.571,1.499  0.870,2.434  0.912,2.139 |  |
| **ADHD (TRF)** | 2997 | 464 |  |  | <0.001 | 1633 | 213 |  |  | 0.002 | 1119 | 123 |  |  | 0.019 |
| No cigarettes (ref) | 1791 | 252 | - | - |  | 970 | 108 | - | - |  | 675 | 65 | - | - |  |
| 1-4 cigarettes | 402 | 52 | 0.907 | 0.659,1.250 |  | 212 | 22 | 0.995 | 0.594,1.666 |  | 144 | 9 | 0.580 | 0.268,1.257 |  |
| 5-9 cigarettes  >10 cigarettes | 282  522 | 57  103 | 1.547  1.501 | 1.124,2.130  1.165,1.934 |  | 156  295 | 25  58 | 1.534  1.855 | 0.910,2.586  1.232,2.791 |  | 98  202 | 13  36 | 1.463  1.950 | 0.722,2.968  1.111,3.422 |  |

Note: CPRS-R – The revised Conners’ Parent Rating Scale; CBCL – Child Behaviour Checklist; TRF – Teacher’s Report Form; N – sample size; n – number of cases; OR – odds ratio; 95% CI – 95% confidence intervals; *adjusted for child’s gender, parity, maternal ethnicity, age, education, anxiety and depression problems, financial difficulties, alcohol use before pregnancy and prenatal caffeine use; **additional adjusted for partner’s smoking before pregnancy

### Supplementary Table S12. Associations of maternal and paternal prenatal smoking on high risk of maternal reported offspring ADHD symptoms in ALSPAC (complete cases)

|  |  | |  | | **Unadjusted model** | | | | **Adjusted model*** | | | **Mutually adjusted model**** | | |
| --- | --- | --- | --- | --- | --- | --- | --- | --- | --- | --- | --- | --- | --- | --- |
|  | **N** | | **n** | | **OR** | **95% CI** | **p-value** | | **OR** | **95% CI** | **p-value** | **OR** | **95% CI** | **p-value** |
| **Maternal** | | | | | | | | | | | | | | |
| **ADHD (DAWBA)** | 5201 | | 709 | |  |  | <0.001 | |  |  | 0.130 |  |  | 0.654 |
| No cigarettes (ref) | 4319 | | 542 | | - | - |  | | - | - |  | - | - |  |
| 1-4 cigarettes | 244 | | 38 | | 1.285 | 0.899,1.838 |  | | 0.964 | 0.663,1.400 |  | 0.843 | 0.576,1.232 |  |
| 5-9 cigarettes | 228 | | 44 | | 1.666 | 1.185,2.344 |  | | 1.223 | 0.846,1.767 |  | 1.068 | 0.734,1.553 |  |
| >10 cigarettes | 410 | | 85 | | 1.823 | 1.412,2.352 |  | | 1.225 | 0.916,1.637 |  | 1.074 | 0.797,1.449 |  |
| **Hyperactive** | 5211 | | 622 | |  |  | <0.001 | |  |  | 0.669 |  |  | 0.839 |
| No cigarettes (ref) | 4329 | | 486 | | - | - |  | | - | - |  | - | - |  |
| 1-4 cigarettes | 245 | | 32 | | 1.188 | 0.810,1.743 |  | | 0.914 | 0.615,1.360 |  | 0.837 | 0.559,1.254 |  |
| 5-9 cigarettes | 227 | | 35 | | 1.441 | 0.993,2.092 |  | | 1.046 | 0.704,1.555 |  | 0.977 | 0.652,1.463 |  |
| >10 cigarettes | 410 | | 69 | | 1.600 | 1.215,2.107 |  | | 1.076 | 0.790,1.465 |  | 0.982 | 0.715,1.348 |  |
| **Inattentive** | 5208 | | 686 | |  |  | <0.001 | |  |  | 0.246 |  |  | 0.651 |
| No cigarettes (ref) | 4326 | | 541 | | - | - |  | | - | - |  | - | - |  |
| 1-4 cigarettes | 243 | | 31 | | 1.023 | 0.694,1.507 |  | | 0.803 | 0.538,1.199 |  | 0.731 | 0.487,1.099 |  |
| 5-9 cigarettes | 228 | | 39 | | 1.444 | 1.011,2.062 |  | | 1.161 | 0.792,1.702 |  | 1.063 | 0.721,1.568 |  |
| >10 cigarettes | 411 | | 75 | | 1.562 | 1.197,2.038 |  | | 1.201 | 0.889,1.622 |  | 1.092 | 0.802,1.488 |  |
| **ADHD (SDQ)** | 5405 | | 561 | |  |  | <0.001 | |  |  | 0.030 |  |  | 0.159 |
| No cigarettes (ref) | 4459 | | 415 | | - | - |  | | - | - |  | - | - |  |
| 1-4 cigarettes | 249 | | 35 | | 1.594 | 1.099,2.310 |  | | 1.290 | 0.880,1.892 |  | 1.172 | 0.792,1.732 |  |
| 5-9 cigarettes | 238 | | 28 | | 1.299 | 0.865,1.952 |  | | 0.951 | 0.619,1.461 |  | 0.878 | 0.568,1.358 |  |
| >10 cigarettes | 459 | | 83 | | 2.151 | 1.661,2.785 |  | | 1.431 | 1.068,1.918 |  | 1.303 | 0.963,1.764 |  |
| **Paternal** | | | | | | | | | | | | | | |
| **ADHD (DAWBA)** | | 4647 | 618 |  | |  | <0.001 |  | |  | 0.001 |  |  | 0.005 |
| No cigarettes (ref) | | 3284 | 381 | - | | - |  | - | | - |  | - | - |  |
| 1-4 cigarettes | | 235 | 34 | 1.289 | | 0.882,1.882 |  | 1.189 | | 0.804,1.758 |  | 1.184 | 0.799,1.755 |  |
| 5-9 cigarettes | | 184 | 28 | 1.368 | | 0.902,2.074 |  | 1.254 | | 0.816,1.930 |  | 1.239 | 0.803,1.912 |  |
| >10 cigarettes | | 944 | 175 | 1.734 | | 1.426,2.109 |  | 1.435 | | 1.155,1.784 |  | 1.378 | 1.097,1.730 |  |
| **Hyperactive** | | 4656 | 532 |  | |  | <0.001 |  | |  | 0.045 |  |  | 0.105 |
| No cigarettes (ref) | | 3291 | 340 | - | | - |  | - | | - |  | - | - |  |
| 1-4 cigarettes | | 235 | 28 | 1.174 | | 0.779,1.770 |  | 1.123 | | 0.737,1.711 |  | 1.112 | 0.728,1.700 |  |
| 5-9 cigarettes | | 183 | 16 | 0.832 | | 0.492,1.406 |  | 0.738 | | 0.431,1.263 |  | 0.727 | 0.424,1.248 |  |
| >10 cigarettes | | 947 | 148 | 1.608 | | 1.305,1.980 |  | 1.309 | | 1.040,1.648 |  | 1.262 | 0.991,1.606 |  |
| **Inattentive** | | 4656 | 610 |  | |  | <0.001 |  | |  | 0.076 |  |  | 0.089 |
| No cigarettes (ref) | | 3289 | 393 | - | | - |  | - | | - |  | - | - |  |
| 1-4 cigarettes | | 237 | 36 | 1.320 | | 0.912,1.911 |  | 1.214 | | 0.828,1.780 |  | 1.236 | 0.841,1.817 |  |
| 5-9 cigarettes | | 184 | 26 | 1.213 | | 0.790,1.860 |  | 1.112 | | 0.715,1.729 |  | 1.128 | 0.723,1.760 |  |
| >10 cigarettes | | 946 | 155 | 1.444 | | 1.180,1.767 |  | 1.220 | | 0.976,1.525 |  | 1.219 | 0.966,1.538 |  |
| **ADHD (SDQ)** | | 4793 |  |  | |  | <0.001 |  | |  | 0.001 |  |  | 0.023 |
| No cigarettes (ref) | | 3353 | 289 | - | | - |  | - | | - |  | - | - |  |
| 1-4 cigarettes | | 239 | 29 | 1.464 | | 0.975,2.199 |  | 1.379 | | 0.909,2.091 |  | 1.322 | 0.869,2.013 |  |
| 5-9 cigarettes | | 195 | 23 | 1.418 | | 0.903,2.227 |  | 1.273 | | 0.802,2.022 |  | 1.215 | 0.762,1.937 |  |
| >10 cigarettes | | 1006 | 144 | 1.771 | | 1.430,2.193 |  | 1.470 | | 1.163,1.857 |  | 1.326 | 1.036,1.699 |  |

Note: DAWBA – Development and Well-Being Assessment; SDQ – Strengths and Difficulties Questionnaire; N – total sample size; n – number of ADHD cases; OR – odds ratio; 95% CI – 95% confidence intervals; *adjusted for child’s gender, ethnicity, parity, parental age, marital status, education, financial difficulties, depression and anxiety symptoms, prenatal alcohol and caffeine use; **additionally adjusted for partners’ prenatal smoking

### Supplementary Table S13. Associations of maternal and paternal prenatal smoking on high risk of maternal reported offspring ADHD symptoms in GenR (complete cases)

|  | |  | |  | | **Unadjusted model** | | | **Adjusted model*** | | | | **Mutually adjusted model**** | | | |
| --- | --- | --- | --- | --- | --- | --- | --- | --- | --- | --- | --- | --- | --- | --- | --- | --- |
|  | | **N** | | **n** | | **OR** | **95% CI** | **p-value** | **OR** | **95% CI** | **p-value** | | **OR** | **95% CI** | | **p-value** |
| **Maternal** | | | | | | | | | | | | | | | | |
| **ADHD (CPRS-R)** | | 1535 | | 196 | |  |  | 0.004 |  |  | 0.013 | |  |  | | 0.018 |
| No cigarettes (ref) | | 1219 | | 145 | | - | - |  | - | - |  | | - | - | |  |
| 1-4 cigarettes | | 165 | | 21 | | 1.080 | 0.662,1.762 |  | 0.999 | 0.600,1.665 |  | | 1.009 | 0.592,1.720 | |  |
| 5-9 cigarettes  >10 cigarettes | | 82  69 | | 13  17 | | 1.396  2.421 | 0.753,2.588  1.363,4.301 |  | 1.287  2.425 | 0.669,2.476  1.301,4.520 |  | | 1.260  2.480 | 0.639,2.484  1.293,4.755 | |  |
| **Hyperactive** | | 1536 | | 143 | |  |  | 0.017 |  |  | 0.034 | |  |  | | 0.217 |
| No cigarettes (ref) | | 1221 | | 107 | | - | - |  | - | - |  | | - | - | |  |
| 1-4 cigarettes | | 165 | | 14 | | 0.965 | 0.539,1.728 |  | 0.985 | 0.540,1.798 |  | | 0.771 | 0.413,1.440 | |  |
| 5-9 cigarettes  >10 cigarettes | | 81  69 | | 9  13 | | 1.301  2.417 | 0.633,2.676  1.281,4.562 |  | 1.205  2.458 | 0.564,2.572  1.230,4.913 |  | | 0.972  1.988 | 0.445,2.123  0.964,4.099 | |  |
| **Inattentive** | | 1537 | | 170 | |  |  | 0.125 |  |  | 0.244 | |  |  | | 0.143 |
| No cigarettes (ref) | | 1221 | | 133 | | - | - |  | - | - |  | | - | - | |  |
| 1-4 cigarettes | | 165 | | 15 | | 0.818 | 0.467,1.433 |  | 0.757 | 0.425,1.349 |  | | 0.806 | 0.442,1.470 | |  |
| 5-9 cigarettes  >10 cigarettes | | 82  69 | | 7  15 | | 0.764  2.272 | 0.345,1.691  1.247,4.139 |  | 0.681  2.200 | 0.299,1.552  1.150,4.206 |  | | 0.681  2.492 | 0.292,1.589  1.259,4.932 | |  |
| **ADHD (CBCL)** | | 1835 | | 211 | |  |  | <0.001 |  |  | <0.001 | |  |  | | 0.031 |
| No cigarettes (ref) | | 1432 | | 140 | | - | - |  | - | - |  | | - | - | |  |
| 1-4 cigarettes | | 211 | | 31 | | 1.589 | 1.045,2.417 |  | 1.528 | 0.983,2.375 |  | | 1.223 | 0.770,1.942 | |  |
| 5-9 cigarettes  >10 cigarettes | | 107  85 | | 21  19 | | 2.253  2.657 | 1.356,3.745  1.549,4.556 |  | 1.943  2.258 | 1.119,3.374  1.255,4.061 |  | | 1.533  1.797 | 0.863,2.725  0.972,3.323 | |  |
| **ADHD (TRF)** | | 1148 | | 125 | |  |  | 0.002 |  |  | 0.017 | |  |  | | 0.059 |
| No cigarettes (ref) | | 878 | | 85 | | - | - |  | - | - |  | | - | - | |  |
| 1-4 cigarettes | | 141 | | 16 | | 1.194 | 0.678,2.104 |  | 1.250 | 0.681,2.293 |  | | 1.005 | 0.529,1.908 | |  |
| 5-9 cigarettes  >10 cigarettes | | 75  54 | | 12  12 | | 1.777  2.666 | 0.922,3.427  1.351,5.258 |  | 1.906  2.358 | 0.907,4.006  1.064,5.224 |  | | 1.594  2.155 | 0.737,3.449  0.944,4.918 | |  |
| **Paternal** | | | | | | | | | | | | | | | | |
| **ADHD (CPRS-R)** | 1930 | | 260 | |  | |  | 0.316 |  |  | 0.605 |  | | |  | 0.237 |
| No cigarettes (ref) | 1148 | | 149 | | - | | - |  | - | - |  | - | | | - |  |
| 1-4 cigarettes | 328 | | 43 | | 1.012 | | 0.703,1.456 |  | 0.904 | 0.619,1.320 |  | 0.891 | | | 0.606,1.310 |  |
| 5-9 cigarettes  >10 cigarettes | 132  322 | | 20  48 | | 1.197  1.175 | | 0.722,1.986  0.826,1.670 |  | 0.930  0.912 | 0.546,1.583  0.625,1.332 |  | 0.873  0.785 | | | 0.502,1.518  0.522,1.180 |  |
| **Hyperactive** | 1930 | | 186 | |  | |  | 0.013 |  |  | 0.330 |  | | |  | 0.448 |
| No cigarettes (ref) | 1148 | | 98 | | - | | - |  | - | - |  | - | | | - |  |
| 1-4 cigarettes | 327 | | 31 | | 1.122 | | 0.734,1.715 |  | 1.040 | 0.669,1.617 |  | 1.088 | | | 0.695,1.704 |  |
| 5-9 cigarettes  >10 cigarettes | 132  323 | | 15  42 | | 1.374  1.601 | | 0.772,2.444  1.090,2.353 |  | 1.049  1.244 | 0.570,1.929  0.820,1.886 |  | 1.122  1.184 | | | 0.598,2.106  0.757,1.852 |  |
| **Inattentive** | 1931 | | 233 | |  | |  | 0.843 |  |  | 0.082 |  | | |  | 0.078 |
| No cigarettes (ref) | 1149 | | 139 | | - | | - |  | - | - |  | - | | | - |  |
| 1-4 cigarettes | 327 | | 40 | | 1.013 | | 0.696,1.474 |  | 0.917 | 0.621,1.355 |  | 0.942 | | | 0.633,1.401 |  |
| 5-9 cigarettes  >10 cigarettes | 132  323 | | 17  37 | | 1.074  0.940 | | 0.626,1.824  0.639,1.382 |  | 0.842  0.693 | 0.478,1.483  0.458,1.049 |  | 0.906  0.658 | | | 0.506,1.621  0.423,1.024 |  |
| **ADHD (CBCL)** | 2323 | | 275 | |  | |  | <0.001 |  |  | 0.003 |  | | |  | 0.039 |
| No cigarettes (ref) | 1361 | | 130 | | - | | - |  | - | - |  | - | | | - |  |
| 1-4 cigarettes | 381 | | 49 | | 1.398 | | 0.985,1.984 |  | 1.349 | 0.936,1.943 |  | 1.324 | | | 0.914,1.919 |  |
| 5-9 cigarettes  >10 cigarettes | 164  417 | | 25  71 | | 1.703  1.943 | | 1.072,2.705  1.421,2.657 |  | 1.436  1.636 | 0.884,2.333  1.167,2.293 |  | 1.331  1.455 | | | 0.807,2.195  1.011,2.093 |  |
| **ADHD (TRF)** | 1445 | | 151 | |  | |  | 0.005 |  |  | 0.211 |  | | |  | 0.347 |
| No cigarettes (ref) | 816 | | 68 | | - | | - |  | - | - |  | - | | | - |  |
| 1-4 cigarettes | 242 | | 29 | | 1.498 | | 0.945,2.374 |  | 1.347 | 0.817,2.220 |  | 1.413 | | | 0.851,2.344 |  |
| 5-9 cigarettes  >10 cigarettes | 109  278 | | 17  37 | | 2.033  1.689 | | 1.145,3.608  1.103,2.585 |  | 1.451  1.302 | 0.781,2.695  0.814,2.081 |  | 1.523  1.211 | | | 0.806,2.876  0.732,2.002 |  |

Note: CPRS-R – The revised Conners’ Parent Rating Scale; CBCL – Child Behaviour Checklist; TRF – Teacher’s Report Form; N – sample size; n – number of cases; OR – odds ratio; 95% CI – 95% confidence intervals; *adjusted for child’s gender, parity, parental ethnicity, age, education, anxiety and depression problems, financial difficulties and parental smoking and prenatal caffeine use in the maternal model; **additionally adjusted for partner’s smoking

### Supplementary Table S14. Associations of maternal and paternal prenatal smoking on high risk of maternal reported offspring ADHD symptoms in MoBa (complete cases)

|  |  | |  | | | **Unadjusted model** | | | | | | **Adjusted model*** | | | | | | **Mutually adjusted model**** | | | | |
| --- | --- | --- | --- | --- | --- | --- | --- | --- | --- | --- | --- | --- | --- | --- | --- | --- | --- | --- | --- | --- | --- | --- |
|  | **N** | | **n** | | | **OR** | | **95% CI** | | **p-value** | | **OR** | **95% CI** | | | **p-value** | | **OR** | | **95% CI** | **p-value** | |
| **Maternal** | | | | | | | | | | | | | | | | | | | | | | |
| **ADHD (RS-DBD)** | 28,055 | | 3543 | | |  | |  | | <0.001 | |  |  | | | 0.001 | |  | |  | 0.006 | |
| No cigarettes (ref) | 26,706 | | 3280 | | | - | | - | |  | | - | - | | |  | | - | | - |  | |
| 1-4 cigarettes | 707 | | 111 | | | 1.330 | | 1.082,1.635 | |  | | 0.980 | 0.788,1.219 | | |  | | 0.948 | | 0.761,1.182 |  | |
| 5-9 cigarettes | 383 | | 87 | | | 2.099 | | 1.645,2.679 | |  | | 1.564 | 1.201,2.038 | | |  | | 1.485 | | 1.136,1.941 |  | |
| >10 cigarettes | 259 | | 65 | | | 2.393 | | 1.810,3.163 | |  | | 1.471 | 1.078,2.008 | | |  | | 1.376 | | 1.000,1.893 |  | |
| **Hyperactive** | 28,051 | | 3507 | | |  | |  | | <0.001 | |  |  | | | <0.001 | |  | |  | <0.001 | |
| No cigarettes (ref) | 26,704 | | 3232 | | | - | | - | |  | | - | - | | |  | | - | | - |  | |
| 1-4 cigarettes | 707 | | 117 | | | 1.440 | | 1.177,1.763 | |  | | 1.076 | 0.871,1.329 | | |  | | 1.039 | | 0.840,1.286 |  | |
| 5-9 cigarettes | 382 | | 91 | | | 2.271 | | 1.789,2.884 | |  | | 1.680 | 1.300,2.170 | | |  | | 1.595 | | 1.228,2.073 |  | |
| >10 cigarettes | 258 | | 67 | | | 2.548 | | 1.924,3.373 | |  | | 1.565 | 1.152,2.127 | | |  | | 1.454 | | 1.061,1.991 |  | |
| **Inattentive** | 28,058 | | 3102 | | |  | |  | | <0.001 | |  |  | | | 0.014 | |  | |  | 0.041 | |
| No cigarettes (ref) | 26,708 | | 2877 | | | - | | - | |  | | - | - | | |  | | - | | - |  | |
| 1-4 cigarettes | 708 | | 101 | | | 1.378 | | 1.110,1.711 | |  | | 1.006 | 0.800,1.265 | | |  | | 0.987 | | 0.783,1.243 |  | |
| 5-9 cigarettes | 383 | | 70 | | | 1.852 | | 1.423,2.412 | |  | | 1.369 | 1.032,1.815 | | |  | | 1.324 | | 0.996,1.760 |  | |
| >10 cigarettes | 259 | | 54 | | | 2.182 | | 1.615,2.947 | |  | | 1.363 | 0.977,1.902 | | |  | | 1.306 | | 0.929,1.837 |  | |
| **Paternal** | | | | | | | | | | | | | | | | | | | | | | |
| **ADHD (RS-DBD)** | | 10,738 | | 1454 |  | |  | | 0.002 | |  | | |  | 0.142 | |  | |  | | | 0.417 |
| No cigarettes (ref) | | 8965 | | 1193 | - | | - | |  | | - | | | - |  | | - | | - | | |  |
| 1-4 cigarettes | | 998 | | 120 | 0.890 | | 0.729,1.088 | |  | | 0.845 | | | 0.690,1.035 |  | | 0.833 | | 0.680,1.021 | | |  |
| 5-9 cigarettes | | 211 | | 40 | 1.524 | | 1.074,2.162 | |  | | 1.375 | | | 0.960,1.969 |  | | 1.317 | | 0.919,1.889 | | |  |
| >10 cigarettes | | 564 | | 101 | 1.421 | | 1.136,1.777 | |  | | 1.187 | | | 0.933,1.511 |  | | 1.090 | | 0.846,1.404 | | |  |
| **Hyperactive** | | 10,736 | | 1424 |  | |  | | 0.001 | |  | | |  | 0.075 | |  | |  | | | 0.242 |
| No cigarettes (ref) | | 8964 | | 1157 | - | | - | |  | | - | | | - |  | | - | | - | | |  |
| 1-4 cigarettes | | 997 | | 132 | 1.030 | | 0.849,1.249 | |  | | 0.986 | | | 0.809,1.201 |  | | 0.976 | | 0.801,1.190 | | |  |
| 5-9 cigarettes | | 211 | | 39 | 1.530 | | 1.075,2.178 | |  | | 1.389 | | | 0.967,1.995 |  | | 1.351 | | 0.939,1.945 | | |  |
| >10 cigarettes | | 564 | | 96 | 1.384 | | 1.100,1.742 | |  | | 1.161 | | | 0.909,1.485 |  | | 1.091 | | 0.843,1.411 | | |  |
| **Inattentive** | | 10,740 | | 1248 |  | |  | | 0.002 | |  | | |  | 0.117 | |  | |  | | | 0.295 |
| No cigarettes (ref) | | 8966 | | 1018 | - | | - | |  | | - | | | - |  | | - | | - | | |  |
| 1-4 cigarettes | | 998 | | 110 | 0.967 | | 0.785,1.191 | |  | | 0.908 | | | 0.734,1.123 |  | | 0.901 | | 0.727,1.115 | | |  |
| 5-9 cigarettes | | 211 | | 31 | 1.345 | | 0.913,1.980 | |  | | 1.191 | | | 0.803,1.767 |  | | 1.163 | | 0.784,1.725 | | |  |
| >10 cigarettes | | 565 | | 89 | 1.460 | | 1.154,1.847 | |  | | 1.223 | | | 0.948,1.579 |  | | 1.157 | | 0.887,1.509 | | |  |

Note: RS-DBD – the Disruptive Behaviour Disorders scale; N – sample size; n – number of cases; OR – odds ratio; 95% CI – 95% confidence intervals;**adjusted for child’s gender, birth year, parity, parental age, education, marital status, financial difficulties, depression and anxiety symptoms, prenatal alcohol and caffeine consumption; ** additionally adjusted for partner’s smoking

### Supplementary Table S15. Associations of maternal weighted PRS on maternal exposure

### phenotypes in ALSPAC

| **Exposure** | **Beta** | **95% CI** | **P-value** | **Sample size** | **R^2^** |
| --- | --- | --- | --- | --- | --- |
| Smoking heaviness | 0.523 | 0.258, 0.788 | <0.001 | 1537 | 0.015 |
| Lifetime smoking* | 0.666 | 0.495, 0.837 | <0.001 | 7107 | 0.010 |
| Lifetime smoking** | 9.090 | 4.521, 18.275 | <0.001 | 3413 | 0.03 |
| Alcohol consumption | 0.287 | 0.074, 0.501 | 0.008 | 3962 | 0.019 |
| Coffee consumption | 53.451 | 30.651, 76.252 | <0.001 | 7074 | 0.004 |

Note: *smoking heaviness phenotype **smoking cessation phenotype (in OR’s);

95% CI – 95% confidence interval; adjusted for principal components

### Supplementary Table S16. Associations of maternal weighted PRS on maternal exposure

### phenotypes in MoBa

| **Exposure** | **Beta** | **95% CI** | **P-value** | **Sample size** | **R^2^** |
| --- | --- | --- | --- | --- | --- |
| Smoking heaviness | 0.393 | 0.112, 0.674 | 0.006 | 1029 | 0.020 |
| Lifetime smoking* | 0.276 | 0.120, 0.351 | <0.001 | 14,488 | 0.012 |
| Lifetime smoking** | 3.21 | 1.544, 6.660 | 0.002 | 3118 | 0.027 |
| Alcohol consumption | 0.649 | -0.757, 2.055 | 0.365 | 1362 | NA |
| Alcohol consumption*** | 1.058 | 0.258, 1.859 | 0.010 | 12,953 | 0.007 |
| Coffee consumption | 18.804 | 9.206, 28.402 | <0.001 | 14,583 | 0.003 |

Note: *smoking heaviness phenotype **smoking cessation phenotype (in OR’s);

95% CI – 95% confidence intervals. ***alcohol consumption before pregnancy. NA – alcohol PRS was not associated with alcohol use during pregnancy; adjusted for principal components, birth year and genotyping batch

### Supplementary Table S17. Associations of maternal unweighted PRS on maternal exposure

### phenotypes in ALSPAC

| **Exposure** | **Beta** | **95% CI** | **P-value** | **Sample size** |
| --- | --- | --- | --- | --- |
| Smoking heaviness | 0.016 | 0.008, 0.023 | <0.001 | 1537 |
| Lifetime smoking* | 0.010 | 0.007, 0.124 | <0.001 | 7107 |
| Lifetime smoking** | 1.033 | 1.022, 1.044 | <0.001 | 3413 |
| Alcohol consumption | 0.004 | 0.002, 0.007 | <0.001 | 3962 |
| Coffee consumption | 3.494 | 1.477, 5.511 | <0.001 | 7074 |

Note: *smoking heaviness phenotype; **smoking cessation phenotype (in OR’s);

95% CI – 95% confidence intervals; adjusted for principal components

### Supplementary Table S18. Associations of maternal unweighted PRS on maternal exposure

### phenotypes in MoBa

| **Exposure** | **Beta** | **95% CI** | **P-value** | **Sample size** |
| --- | --- | --- | --- | --- |
| Smoking heaviness | 0.010 | 0.002, 0.180 | 0.021 | 1029 |
| Lifetime smoking* | 0.004 | 0.003, 0.005 | <0.001 | 14,488 |
| Lifetime smoking** | 1.016 | 1.005, 1.027 | 0.004 | 3118 |
| Alcohol consumption | 0.001 | -0.016, 0.018 | 0.911 | 1362 |
| Alcohol consumption*** | 1.058 | 0.258, 1.859 | 0.010 | 12,953 |
| Coffee consumption | 1.144 | 0.279, 2.008 | 0.010 | 14,583 |

Note: *smoking heaviness phenotype; **smoking cessation phenotype (in OR’s);

95% CI – 95% confidence intervals; ***alcohol consumption before pregnancy; adjusted for principal components, birth year and genotyping batch

### Supplementary Table S19. Associations of maternal smoking heaviness PRS on confounders in ALSPAC

| **Confounder** | **Effect estimate** | **Effect size** | **95% CI** | **P-value** | **Sample size** |
| --- | --- | --- | --- | --- | --- |
| Maternal age | beta | 0.043 | -1.437, 1.524 | 0.954 | 1853 |
| Maternal education | beta | -0.380 | -0.782, 0.023 | 0.065 | 1527 |
| Financial difficulties | beta | 0.506 | -0.816, 1.826 | 0.453 | 1475 |
| Marital status | OR | 1.242 | 0.662, 2.330 | 0.500 | 1653 |
| Depression symptoms | OR | 0.895 | 0.395, 2.027 | 0.791 | 1457 |
| Anxiety symptoms | OR | 0.988 | 0.452, 2.162 | 0.976 | 1451 |
| Parity | beta | 0.038 | -0.265, 0.341 | 0.806 | 1540 |

Note: adjusted for principal components; OR- odds ratio; 95% CI – 95% confidence intervals.

### Supplementary Table S20. Associations of maternal smoking heaviness PRS on confounders in MoBa

| **Confounder** | **Effect estimate** | **Effect size** | **95% CI** | **P-value** | **Sample size** |
| --- | --- | --- | --- | --- | --- |
| Maternal age | beta | -0.211 | -13.382, 12.96 | 0.975 | 1125 |
| Maternal education | beta | -0.123 | -0.330, 0.084 | 0.242 | 1069 |
| Financial difficulties | OR | 1.101 | 0.502, 2.416 | 0.810 | 966 |
| Marital status | beta | -0.035 | -0.171, 0101 | 0.611 | 1120 |
| Depr/Anxiety symptoms | OR | 1.392 | 0.489, 3.958 | 0.535 | 1112 |
| Maternal ADHD | OR | 0.591 | 0.061, 5.694 | 0.649 | 557 |
| Parity | beta | -0.412 | -0.732, -0.092 | 0.012 | 1125 |

Note: adjusted for principal components, birth year and genotyping batch; OR – odds ratio; 95% CI – 95% confidence intervals.

### Supplementary Table S21. Associations of maternal lifetime smoking PRS on confounders in ALSPAC

| **Confounder** | **Effect estimate** | **Effect size** | **95% CI** | **P-value** | **Sample size** |
| --- | --- | --- | --- | --- | --- |
| Maternal age | beta | -2.637 | -3.688, -1.586 | <0.001 | 7421 |
| Maternal education | beta | -0.999 | -1.286, -0.711 | <0.001 | 6860 |
| Financial difficulties | beta | 1.115 | 0.317, 1.912 | 0.006 | 6691 |
| Marital status | OR | 0.240 | 0.138, 0.415 | <0.001 | 7124 |
| Depression symptoms | OR | 1.849 | 0.910, 3.757 | 0.089 | 6706 |
| Anxiety symptoms | OR | 1.984 | 1.035, 3.801 | 0.039 | 6669 |
| Parity | beta | 0.086 | -0.111, 0.282 | 0.392 | 7040 |

Note: adjusted for principal components; OR – odds ratio; 95% CI – 95% confidence intervals.

### Supplementary Table S22. Associations of maternal lifetime smoking PRS on confounders in MoBa

| **Confounder** | **Effect estimate** | **Effect size** | **95% CI** | **P-value** | **Sample size** |
| --- | --- | --- | --- | --- | --- |
| Maternal age | beta | -0.686 | -3.095, 1.723 | 0.577 | 14,584 |
| Maternal education | beta | -0.274 | -0.356, -0.191 | <0.001 | 13,836 |
| Financial difficulties | OR | 1.570 | 0.995, 2.477 | 0.053 | 13,484 |
| Marital status | beta | 0.006 | -0.027, 0.038 | 0.718 | 14,519 |
| Depr/Anxiety symptoms | OR | 1.975 | 1.052, 3.705 | 0.034 | 14,464 |
| Maternal ADHD | OR | 1.897 | 0.562, 6.403 | 0.302 | 8841 |
| Parity | beta | 0.067 | -0.065, 0.200 | 0.319 | 14,584 |

Note: adjusted for principal components, birth year, genotyping batch; OR- odds ratio; 95% CI – 95% confidence intervals.

### Supplementary Table S23. Associations of maternal smoking heaviness PRS on high risk of maternal reported offspring ADHD symptoms in ALSPAC

|  | **Maternal PRS** | | | **Maternal PRS adj. for PC** | | |  |
| --- | --- | --- | --- | --- | --- | --- | --- |
| **Outcome** | **OR** | **95% CI** | **P-value** | **OR** | **95% CI** | **P-value** | **Sample size** |
| ADHD (DAWBA) | 1.043 | 0.377, 2.885 | 0.936 | 1.087 | 0.389, 3.037 | 0.873 | 958 |
| Hyperactive | 0.461 | 0.156, 1.367 | 0.163 | 0.453 | 0.151, 1.362 | 0.159 | 959 |
| Inattentive | 0.861 | 0.289, 2.561 | 0.787 | 0.888 | 0.294, 2.680 | 0.833 | 958 |
| ADHD (SDQ) | 0.124 | 0.038, 0.403 | 0.001 | 0.123 | 0.037, 0.407 | 0.001 | 979 |

Note: DAWBA – Development and Well-Being Assessment; SDQ – Strengths and Difficulties Questionnaire; OR – odds ratio; 95% CI – 95% confidence intervals; PC – principal components

### Supplementary Table S24. Associations of maternal smoking heaviness PRS on high risk of teacher reported offspring ADHD symptoms in ALSPAC

|  | **Maternal PRS** | | | **Maternal PRS adj. for PC** | | |  |
| --- | --- | --- | --- | --- | --- | --- | --- |
| **Outcome** | **OR** | **95% CI** | **P-value** | **OR** | **95% CI** | **P-value** | **Sample size** |
| ADHD (DAWBA) | 2.512 | 0.816, 7.735 | 0.108 | 2.395 | 0.764, 7.510 | 0.134 | 833 |
| Hyperactive | 2.299 | 0.736, 7.180 | 0.152 | 2.186 | 0.688, 6.947 | 0.185 | 833 |
| Inattentive | 2.998 | 0.888, 10.122 | 0.077 | 2.680 | 0.781, 9.198 | 0.117 | 833 |
| ADHD (SDQ) | 1.383 | 0.419, 4.560 | 0.595 | 1.305 | 0.390, 4.364 | 0.666 | 834 |

Note: DAWBA – Development and Well-Being Assessment; SDQ – Strengths and Difficulties Questionnaire; OR – odds ratio; 95% CI – 95% confidence intervals; PC – principal components

### Supplementary Table S25. Associations of maternal smoking heaviness PRS on high risk of maternal reported offspring ADHD symptoms in MoBa

|  | **Maternal PRS** | | | **Maternal PRS adj. for PC** | | |  |
| --- | --- | --- | --- | --- | --- | --- | --- |
| **Outcome** | **OR** | **95% CI** | **P-value** | **OR** | **95% CI** | **P-value** | **Sample size** |
| ADHD (RS-DBD) | 1.768 | 0.423, 7.385 | 0.435 | 1.860 | 0.430, 8.057 | 0.406 | 396 |
| Hyperactive | 0.801 | 0.195, 3.282 | 0.757 | 0.883 | 0.211, 3.693 | 0.865 | 394 |
| Inattentive | 2.821 | 0.608, 13.097 | 0.186 | 3.050 | 0.639, 14.564 | 0.162 | 396 |

Note: RS-DBD – The Disruptive Behavior Disorders Scale; OR – odds ratio; 95% CI – 95% confidence intervals; adjusted for principal components (PC), birth year and genotyping batch

### Supplementary Table S26. Associations of maternal lifetime smoking PRS on high risk of maternal reported offspring ADHD symptoms in ALSPAC

|  | **Maternal PRS** | | | **Maternal PRS adj. for PC** | | |  |
| --- | --- | --- | --- | --- | --- | --- | --- |
| **Outcome** | **OR** | **95% CI** | **P-value** | **OR** | **95% CI** | **P-value** | **Sample size** |
| ADHD (DAWBA) | 0.913 | 0.421, 1.979 | 0.818 | 0.899 | 0.414, 1.950 | 0.787 | 5005 |
| Hyperactive | 0.855 | 0.380, 1.927 | 0.706 | 0.833 | 0.369, 1.881 | 0.661 | 5016 |
| Inattentive | 0.988 | 0.451, 2.168 | 0.977 | 0.976 | 0.445, 2.142 | 0.952 | 5013 |
| ADHD (SDQ) | 1.529 | 0.641, 3.650 | 0.338 | 1.528 | 0.638, 3.658 | 0.341 | 5103 |

Note: DAWBA – Development and Well-Being Assessment; SDQ – Strengths and Difficulties Questionnaire; OR – odds ratio; 95% CI – 95% confidence intervals; PC – principal components

### Supplementary Table S27. Associations of maternal lifetime smoking PRS on high risk of teacher reported offspring ADHD symptoms in ALSPAC

|  | **Maternal PRS** | | | **Maternal PRS adj. for PC** | | |  |
| --- | --- | --- | --- | --- | --- | --- | --- |
| **Outcome** | **OR** | **95% CI** | **P-value** | **OR** | **95% CI** | **P-value** | **Sample size** |
| ADHD (DAWBA) | 2.683 | 1.025, 7.022 | 0.044 | 2.695 | 1.026, 7.079 | 0.044 | 3486 |
| Hyperactive | 2.392 | 0.883, 6.478 | 0.086 | 2.428 | 0.893, 6.603 | 0.082 | 3485 |
| Inattentive | 1.909 | 0.685, 5.321 | 0.216 | 1.907 | 0.682, 5.332 | 0.219 | 3487 |
| ADHD (SDQ) | 2.966 | 1.029, 8.548 | 0.044 | 2.997 | 1.034, 8.688 | 0.043 | 3486 |

Note: DAWBA – Development and Well-Being Assessment; SDQ – Strengths and Difficulties Questionnaire; OR – odds ratio; 95% CI – 95% confidence intervals; PC – principal components

### Supplementary Table S28. Associations of maternal lifetime smoking PRS on high risk of maternal reported offspring ADHD symptoms in MoBa

|  | **Maternal PRS** | | | **Maternal PRS adj. for PC** | | |  |
| --- | --- | --- | --- | --- | --- | --- | --- |
| **Outcome** | **OR** | **95% CI** | **P-value** | **OR** | **95% CI** | **P-value** | **Sample size** |
| ADHD (RS-DBD) | 1.019 | 0.510, 2.036 | 0.958 | 1.019 | 0.510, 2.040 | 0.957 | 7017 |
| Hyperactive | 1.017 | 0.512, 2.021 | 0.961 | 1.019 | 0.513, 2.025 | 0.957 | 7012 |
| Inattentive | 1.055 | 0.510, 2.181 | 0.885 | 1.071 | 0.518, 2.216 | 0.853 | 7017 |

Note: RS-DBD – The Disruptive Behavior Disorders Scale; OR – odds ratio; 95% CI – 95% confidence intervals; adjusted for principal components, birth year and genotyping batch

### Supplementary Table S29. Associations of maternal and paternal prenatal alcohol consumption on high risk of maternal reported offspring ADHD symptoms in ALSPAC

|  | |  | |  | | **Unadjusted model** | | | | |  | |  | **Adjusted model*** | | | |  | |  | **Mutually adjusted model**** | | | | | |
| --- | --- | --- | --- | --- | --- | --- | --- | --- | --- | --- | --- | --- | --- | --- | --- | --- | --- | --- | --- | --- | --- | --- | --- | --- | --- | --- |
|  | | **N** | | **n** | | **OR** | **95% CI** | | **p-value** | | **N** | | **n** | **OR** | | **95% CI** | **p-value** | **N** | | **n** | **OR** | | **95% CI** | | **p-value** | |
| **Maternal** | | | | | | | | | | | | | | | | | | | | | | | | | | |
| **ADHD (DAWBA)** | | 7711 | | 1114 | |  |  | | <0.001 | | 6675 | | 934 |  | |  | 0.010 | 5384 | | 733 |  | |  | | 0.002 | |
| None (ref) | | 3411 | | 450 | | - | - | |  | | 2938 | | 374 | - | | - |  | 2369 | | 288 | - | | - | |  | |
| <1 unit a week | | 3119 | | 455 | | 1.124 | 0.977,1.293 | |  | | 2725 | | 389 | 1.163 | | 0.993,1.362 |  | 2220 | | 311 | 1.229 | | 1.026,1.471 | |  | |
| >1 unit a week | | 1181 | | 209 | | 1.415 | 1.183,1.693 | |  | | 1012 | | 171 | 1.287 | | 1.045,1.586 |  | 795 | | 134 | 1.437 | | 1.128,1.832 | |  | |
| **Hyperactive** | | 7732 | | 982 | |  |  | | 0.009 | | 6693 | | 828 |  | |  | 0.048 | 5394 | | 642 |  | |  | | 0.009 | |
| None (ref) | | 3426 | | 407 | | - | - | |  | | 2951 | | 338 | - | | - |  | 2378 | | 260 | - | | - | |  | |
| <1 unit a week | | 3124 | | 398 | | 1.083 | 0.934,1.255 | |  | | 2729 | | 346 | 1.146 | | 0.972,1.350 |  | 2221 | | 272 | 1.196 | | 0.991,1.443 | |  | |
| >1 unit a week | | 1182 | | 177 | | 1.306 | 1.080,1.580 | |  | | 1013 | | 144 | 1.210 | | 0.971,1.507 |  | 795 | | 110 | 1.377 | | 1.064,1.781 | |  | |
| **Inattentive** | | 7723 | | 1057 | |  |  | | 0.011 | | 6688 | | 890 |  | |  | 0.061 | 5392 | | 708 |  | |  | | 0.004 | |
| None (ref) | | 3416 | | 437 | | - | - | |  | | 2943 | | 363 | - | | - |  | 2375 | | 279 | - | | - | |  | |
| <1 unit a week | | 3125 | | 434 | | 1.099 | 0.953,1.268 | |  | | 2723 | | 373 | 1.150 | | 0.980,1.348 |  | 2222 | | 301 | 1.236 | | 1.031,1.483 | |  | |
| >1 unit a week | | 1182 | | 186 | | 1.273 | 1.057,1.533 | |  | | 1013 | | 154 | 1.194 | | 0.964,1.480 |  | 795 | | 128 | 1.414 | | 1.106,1.808 | |  | |
| **ADHD (SDQ)** | | 7983 | | 890 | |  |  | | 0.003 | | 6946 | | 737 |  | |  | 0.019 | 5613 | | 579 |  | |  | | 0.008 | |
| None (ref) | | 3521 | | 356 | | - | - | |  | | 3055 | | 291 | - | | - |  | 2471 | | 222 | - | | - | |  | |
| <1 unit a week | | 3225 | | 373 | | 1.163 | 0.997,1.356 | |  | | 2837 | | 317 | 1.232 | | 1.037,1.464 |  | 2310 | | 257 | 1.317 | | 1.082,1.603 | |  | |
| >1 unit a week | | 1237 | | 161 | | 1.330 | 1.091,1.622 | |  | | 1054 | | 129 | 1.266 | | 1.005,1.593 |  | 832 | | 100 | 1.362 | | 1.041,1.781 | |  | |
| **Paternal** | | | | | | | | | | | | | | | | | | | | | | | | | | |
| **ADHD (DAWBA)** | 6049 | | 843 | |  | | |  | | 0.019 | 4657 | 622 | | |  |  | 0.201 | | 4648 | 622 | |  | |  | | 0.048 |
| None (ref) | 226 | | 49 | | - | | | - | |  | 148 | 36 | | | - | - |  | | 148 | 36 | | - | | - | |  |
| <1 unit a week | 1404 | | 202 | | 0.607 | | | 0.428,0.861 | |  | 1054 | 136 | | | 0.485 | 0.314,0.748 |  | | 1053 | 136 | | 0.452 | | 0.292,0.701 | |  |
| 1-6 units a week | 3164 | | 424 | | 0.559 | | | 0.401,0.779 | |  | 2471 | 320 | | | 0.527 | 0.349,0.795 |  | | 2465 | 320 | | 0.471 | | 0.310,0.715 | |  |
| >1 unit a day | 1255 | | 168 | | 0.558 | | | 0.391,0.797 | |  | 984 | 130 | | | 0.507 | 0.327,0.787 |  | | 982 | 130 | | 0.436 | | 0.279,0.683 | |  |
| **Hyperactive** | 6059 | | 735 | |  | | |  | | <0.001 | 4666 | 535 | | |  |  | 0.018 | | 4657 | 535 | |  | |  | | 0.004 |
| None (ref) | 226 | | 43 | | - | | | - | |  | 148 | 30 | | | - | - |  | | 148 | 30 | | - | | - | |  |
| <1 unit a week | 1410 | | 194 | | 0.679 | | | 0.471,0.978 | |  | 1060 | 131 | | | 0.604 | 0.383,0.953 |  | | 1059 | 131 | | 0.563 | | 0.350,0.905 | |  |
| 1-6 units a week | 3170 | | 365 | | 0.554 | | | 0.390,0.786 | |  | 2476 | 275 | | | 0.579 | 0.374,0.895 |  | | 2470 | 275 | | 0.541 | | 0.343,0.855 | |  |
| >1 unit a day | 1253 | | 133 | | 0.505 | | | 0.346,0.737 | |  | 982 | 99 | | | 0.498 | 0.312,0.796 |  | | 980 | 99 | | 0.471 | | 0.288,0.771 | |  |
| **Inattentive** | 6058 | | 804 | |  | | |  | | 0.217 | 4666 | 614 | | |  |  | 0.359 | | 4657 | 614 | |  | |  | | 0.102 |
| None (ref) | 227 | | 45 | | - | | | - | |  | 148 | 32 | | | - | - |  | | 148 | 32 | | - | | - | |  |
| <1 unit a week | 1407 | | 192 | | 0.639 | | | 0.446,0.916 | |  | 1057 | 141 | | | 0.591 | 0.379,0.922 |  | | 1056 | 141 | | 0.505 | | 0.320,0.796 | |  |
| 1-6 units a week | 3167 | | 390 | | 0.568 | | | 0.403,0.800 | |  | 2473 | 304 | | | 0.561 | 0.367,0.859 |  | | 2467 | 304 | | 0.451 | | 0.291,0.699 | |  |
| >1 unit a day | 1257 | | 177 | | 0.663 | | | 0.461,0.953 | |  | 988 | 137 | | | 0.621 | 0.396,0.974 |  | | 986 | 137 | | 0.491 | | 0.308,0.781 | |  |
| **ADHD (SDQ)** | 6264 | | 669 | |  | | |  | | 0.040 | 4804 | 487 | | |  |  | 0.436 | | 4796 | 487 | |  | |  | | 0.156 |
| None (ref) | 233 | | 35 | | - | | | - | |  | 160 | 24 | | | - | - |  | | 160 | 24 | | - | | - | |  |
| <1 unit a week | 1467 | | 172 | | 0.751 | | | 0.507,1.113 | |  | 1096 | 114 | | | 0.681 | 0.418,1.110 |  | | 1095 | 114 | | 0.620 | | 0.374,1.027 | |  |
| 1-6 units a week | 3276 | | 327 | | 0.627 | | | 0.430,0.915 | |  | 2541 | 245 | | | 0.664 | 0.416,1.061 |  | | 2537 | 245 | | 0.579 | | 0.356,0.941 | |  |
| >1 unit a day | 1288 | | 135 | | 0.662 | | | 0.443,0.989 | |  | 1007 | 104 | | | 0.686 | 0.418,1.127 |  | | 1004 | 104 | | 0.552 | | 0.328,0.928 | |  |

Note: DAWBA – Development and Well-Being Assessment; SDQ – Strengths and Difficulties Questionnaire; N – total sample size; n – number of ADHD cases; OR – odds ratio; 95% CI – 95% confidence intervals; *adjusted for child’s gender, ethnicity, parity, parental age, marital status, education, financial difficulties, depression and anxiety symptoms, prenatal smoking and caffeine use; **additionally adjusted for partner’s prenatal alcohol use

### Supplementary Table S30. Associations of maternal and paternal prenatal alcohol consumption on high risk of maternal and teacher reported offspring ADHD symptoms in GenR

|  |  |  | **Unadjusted model** | | |  |  | **Adjusted model*** | | |  |  | **Mutually adjusted model**** | | |
| --- | --- | --- | --- | --- | --- | --- | --- | --- | --- | --- | --- | --- | --- | --- | --- |
|  | **N** | **n** | **OR** | **95% CI** | **p-value** | **N** | **n** | **OR** | **95% CI** | **p-value** | **N** | **n** | **OR** | **95% CI** | **p-value** |
| **Maternal** | | | | | | | | | | | | | | | |
| **ADHD (CPRS-R)** | 3030 | 440 |  |  | 0.414 | 1983 | 274 |  |  | 0.484 | 1532 | 196 |  |  | 0.581 |
| None (ref) | 1521 | 210 | - | - |  | 920 | 120 | - | - |  | 630 | 72 | - | - |  |
| <1 unit a week | 933 | 145 | 1.149 | 0.913,1.445 |  | 644 | 93 | 1.110 | 0.809,1.522 |  | 507 | 71 | 1.251 | 0.854,1.834 |  |
| >1 unit a week | 576 | 85 | 1.081 | 0.823,1.419 |  | 419 | 61 | 1.129 | 0.781,1.633 |  | 395 | 53 | 1.118 | 0.720,1.735 |  |
| **Hyperactive** | 3033 | 337 |  |  | 0.053 | 1987 | 194 |  |  | 0.198 | 1533 | 144 |  |  | 0.164 |
| None (ref) | 1523 | 186 | - | - |  | 923 | 98 | - | - |  | 631 | 66 | - | - |  |
| <1 unit a week | 933 | 96 | 0.824 | 0.635,1.070 |  | 644 | 60 | 0.842 | 0.586,1.210 |  | 507 | 44 | 0.784 | 0.509,1.207 |  |
| >1 unit a week | 577 | 55 | 0.757 | 0.551,1.040 |  | 420 | 36 | 0.762 | 0.492,1.180 |  | 395 | 34 | 0.712 | 0.433,1.169 |  |
| **Inattentive** | 3032 | 379 |  |  | 0.656 | 1988 | 230 |  |  | 0.698 | 1534 | 170 |  |  | 0.498 |
| None (ref) | 1523 | 182 | - | - |  | 924 | 100 | - | - |  | 632 | 60 | - | - |  |
| <1 unit a week | 932 | 127 | 1.162 | 0.912,1.482 |  | 644 | 82 | 1.188 | 0.851,1.659 |  | 507 | 62 | 1.276 | 0.851,1.913 |  |
| >1 unit a week | 577 | 70 | 1.017 | 0.758,1.365 |  | 420 | 48 | 1.047 | 0.702,1.563 |  | 395 | 48 | 1.161 | 0.732,1.842 |  |
| **ADHD (CBCL)** | 4075 | 585 |  |  | 0.002 | 2485 | 321 |  |  | 0.170 | 1828 | 212 |  |  | 0.105 |
| None (ref) | 2210 | 347 | - | - |  | 1221 | 167 | - | - |  | 809 | 101 | - | - |  |
| <1 unit a week | 1159 | 159 | 0.854 | 0.697,1.046 |  | 769 | 103 | 1.044 | 0.780,1.397 |  | 574 | 67 | 0.940 | 0.651,1.356 |  |
| >1 unit a week | 706 | 79 | 0.676 | 0.521,0.878 |  | 495 | 51 | 0.734 | 0.507,1.064 |  | 445 | 44 | 0.684 | 0.441,1.062 |  |
| **ADHD (TRF)** | 2970 | 458 |  |  | <0.001 | 1622 | 208 |  |  | 0.129 | 1148 | 126 |  |  | 0.178 |
| None (ref) | 1784 | 333 | - | - |  | 849 | 129 | - | - |  | 539 | 72 | - | - |  |
| <1 unit a week | 723 | 76 | 0.512 | 0.392,0.668 |  | 469 | 48 | 0.712 | 0.480,1.056 |  | 342 | 28 | 0.601 | 0.357,1.013 |  |
| >1 unit a week | 463 | 49 | 0.516 | 0.375,0.710 |  | 304 | 31 | 0.748 | 0.466,1.201 |  | 267 | 26 | 0.715 | 0.404,1.264 |  |
| **Paternal** | | | | | | | | | | | | | | | |
| **ADHD (CPRS-R)** | 2373 | 333 |  |  | 0.266 | 2117 | 289 |  |  | 0.070 | 1937 | 261 |  |  | 0.163 |
| None (ref) | 279 | 28 | - | - |  | 221 | 19 | - | - |  | 201 | 19 | - | - |  |
| <1 unit a week | 311 | 54 | 1.884 | 1.156,3.070 |  | 266 | 42 | 2.026 | 1.107,3.708 |  | 238 | 35 | 1.575 | 0.841,2.948 |  |
| 1-6 units a week  >1 unit a day | 1166  617 | 159  92 | 1.415  1.571 | 0.926,2.165  1.003,2.461 |  | 1069  561 | 146  82 | 1.850  2.056 | 1.069,3.202  1.146,3.688 |  | 982  516 | 133  74 | 1.555  1.700 | 0.880,2.746  0.921,3.139 |  |
| **Hyperactive** | 2373 | 242 |  |  | 0.901 | 2117 | 202 |  |  | 0.636 | 1937 | 186 |  |  | 0.401 |
| None (ref) | 279 | 26 | - | - |  | 221 | 17 | - | - |  | 201 | 17 | - | - |  |
| <1 unit a week | 311 | 42 | 1.519 | 0.905,2.551 |  | 266 | 35 | 1.840 | 0.962,3.519 |  | 238 | 31 | 1.683 | 0.866,3.270 |  |
| 1-6 units a week  >1 unit a day | 1166  617 | 104  70 | 0.953  1.245 | 0.607,1.496  0.775,2.001 |  | 1069  561 | 90  60 | 1.174  1.497 | 0.646,2.137  0.792,2.828 |  | 982  516 | 83  55 | 1.180  1.568 | 0.637,2.185  0.808,3.045 |  |
| **Inattentive** | 2374 | 290 |  |  | 0.208 | 2119 | 256 |  |  | 0.111 | 1939 | 234 |  |  | 0.144 |
| None (ref) | 279 | 29 | - | - |  | 221 | 19 | - | - |  | 201 | 18 | - | - |  |
| <1 unit a week | 311 | 41 | 1.309 | 0.789,2.171 |  | 266 | 36 | 1.554 | 0.838,2.884 |  | 238 | 31 | 1.375 | 0.719,2.629 |  |
| 1-6 units a week  >1 unit a day | 1165  619 | 133  87 | 1.111  1.410 | 0.727,1.699  0.902,2.203 |  | 1069  563 | 123  78 | 1.374  1.735 | 0.788,2.395  0.963,3.126 |  | 982  518 | 113  72 | 1.283  1.627 | 0.714,2.306  0.869,3.045 |  |
| **ADHD (CBCL)** | 2898 | 363 |  |  | 0.320 | 2541 | 305 |  |  | 0.913 | 2332 | 278 |  |  | 0.865 |
| None (ref) | 413 | 52 | - | - |  | 319 | 40 | - | - |  | 299 | 39 | - | - |  |
| <1 unit a week | 413 | 66 | 1.320 | 0.892,1.954 |  | 350 | 54 | 1.414 | 0.885,2.258 |  | 317 | 48 | 1.382 | 0.848,2.251 |  |
| 1-6 units a week  >1 unit a day | 1376  696 | 160  85 | 0.913  0.966 | 0.654,1.276  0.668,1.396 |  | 1244  628 | 137  74 | 1.019  1.141 | 0.668,1.555  0.710,1.833 |  | 1139  577 | 126  65 | 1.093  1.173 | 0.699,1.710  0.701,1.953 |  |
| **ADHD (TRF)** | 1876 | 216 |  |  | 0.029 | 1570 | 166 |  |  | 0.829 | 1452 | 149 |  |  | 0.680 |
| None (ref) | 325 | 49 | - | - |  | 229 | 31 | - | - |  | 213 | 28 | - | - |  |
| <1 unit a week | 261 | 31 | 0.759 | 0.469,1.230 |  | 210 | 19 | 0.784 | 0.400,1.536 |  | 196 | 18 | 0.851 | 0.411,1.713 |  |
| 1-6 units a week  >1 unit a day | 865  425 | 93  43 | 0.679  0.634 | 0.468,0.985  0.409,0.982 |  | 756  375 | 79  37 | 1.047  0.963 | 0.616,1.779  0.519,1.789 |  | 695  348 | 70  33 | 1.145  1.050 | 0.643,2.039  0.534,2.063 |  |

Note: CPRS-R – The revised Conners’ Parent Rating Scale; CBCL – Child Behaviour Checklist; TRF – Teacher’s Report Form; N – sample size; n – number of cases; OR – odds ratio; 95% CI – 95% confidence intervals; *adjusted for child’s gender, parity, parental ethnicity, age, education, anxiety and depression problems, financial difficulties, smoking and parental caffeine use in the maternal model; **additionally adjusted for partner’s alcohol use

### Supplementary Table S31. Associations of maternal and paternal prenatal alcohol consumption on high risk of maternal reported offspring ADHD symptoms in MoBa

|  |  |  | **Unadjusted model** | | |  |  | **Adjusted model*** | | |  |  | **Mutually adjusted model**** | | |
| --- | --- | --- | --- | --- | --- | --- | --- | --- | --- | --- | --- | --- | --- | --- | --- |
|  | **N** | **n** | **OR** | **95% CI** | **p-value** | **N** | **n** | **OR** | **95% CI** | **p-value** | **N** | **n** | **OR** | **95% CI** | **p-value** |
| **Maternal** | | | | | | | | | | | | | | | |
| **ADHD (RS-DBD)** | 38,134 | 5030 |  |  | <0.001 | 34,297 | 4415 |  |  | <0.001 | 10,641 | 1395 |  |  | <0.001 |
| None (ref) | 33,605 | 4342 | - | - |  | 30,216 | 3791 | - | - |  | 9800 | 1247 | - | - |  |
| <1 unit a week | 4366 | 663 | 1.207 | 1.104,1.319 |  | 3931 | 601 | 1.328 | 1.202,1.466 |  | 811 | 146 | 1.533 | 1.260,1.865 |  |
| >1 unit a week | 163 | 25 | 1.221 | 0.795,1.875 |  | 150 | 23 | 1.124 | 0.709,1.781 |  | 30 | 2 | 0.374 | 0.085,1.647 |  |
| **Hyperactive** | 38,127 | 4957 |  |  | <0.001 | 34,290 | 4353 |  |  | <0.001 | 10,638 | 1351 |  |  | 0.005 |
| None (ref) | 33,601 | 4281 | - | - |  | 30,211 | 3741 | - | - |  | 9797 | 1214 | - | - |  |
| <1 unit a week | 4363 | 653 | 1.205 | 1.102,1.319 |  | 3929 | 591 | 1.287 | 1.166,1.421 |  | 811 | 133 | 1.395 | 1.139,1.710 |  |
| >1 unit a week | 163 | 23 | 1.125 | 0.722,1.754 |  | 150 | 21 | 0.999 | 0.625,1.597 |  | 30 | 4 | 0.890 | 0.283,2.796 |  |
| **Inattentive** | 38,140 | 4393 |  |  | <0.001 | 34,302 | 3874 |  |  | <0.001 | 10,639 | 1210 |  |  | 0.002 |
| None (ref) | 33,610 | 3786 | - | - |  | 30,220 | 3326 | - | - |  | 9799 | 1087 | - | - |  |
| <1 unit a week | 4367 | 584 | 1.216 | 1.106,1.337 |  | 3932 | 527 | 1.310 | 1.180,1.455 |  | 810 | 120 | 1.452 | 1.176,1.793 |  |
| >1 unit a week | 163 | 23 | 1.294 | 0.833,2.011 |  | 150 | 21 | 1.196 | 0.744,1.924 |  | 30 | 3 | 0.740 | 0.197,2.777 |  |
| **Paternal** | | | | | | | | | | | | | | | |
| **ADHD (RS-DBD)** | 12,820 | 1723 |  |  | 0.594 | 10,804 | 1462 |  |  | 0.807 | 9861 | 1322 |  |  | 0.366 |
| None (ref) | 1904 | 255 | - | - |  | 613 | 86 | - | - |  | 586 | 82 | - | - |  |
| <1 unit a week | 3405 | 463 | 1.018 | 0.863,1.200 |  | 3192 | 438 | 0.999 | 0.770,1.297 |  | 2945 | 403 | 0.971 | 0.742,1.270 |  |
| 1-6 units a week | 4684 | 601 | 0.952 | 0.812,1.277 |  | 4360 | 556 | 0.889 | 0.686,1.360 |  | 3971 | 501 | 0.850 | 0.651,1.284 |  |
| >1 unit per day | 2827 | 404 | 1.078 | 0.910,1.805 |  | 2639 | 382 | 1.037 | 0.791,1.805 |  | 2359 | 336 | 0.969 | 0.731,1.805 |  |
| **Hyperactive** | 12,818 | 1684 |  |  | 0.833 | 10,802 | 1432 |  |  | 0.837 | 9858 | 1294 |  |  | 0.616 |
| None (ref) | 1904 | 256 | - | - |  | 613 | 89 | - | - |  | 586 | 86 | - | - |  |
| <1 unit a week | 3404 | 452 | 0.986 | 0.834,1.164 |  | 3191 | 421 | 0.917 | 0.711,1.182 |  | 2944 | 387 | 0.877 | 0.676,1.137 |  |
| 1-6 units a week | 4683 | 580 | 0.910 | 0.776,1.244 |  | 4359 | 545 | 0.845 | 0.656,1.299 |  | 3969 | 490 | 0.793 | 0.611,1.206 |  |
| >1 unit per day | 2827 | 396 | 1.049 | 0.884,1.805 |  | 2639 | 377 | 0.997 | 0.765,1.805 |  | 2359 | 331 | 0.917 | 0.698,1.805 |  |
| **Inattentive** | 12,821 | 1486 |  |  | 0.802 | 10,806 | 1254 |  |  | 0.660 | 9862 | 1129 |  |  | 0.293 |
| None (ref) | 1903 | 233 | - | - |  | 613 | 76 | - | - |  | 586 | 72 | - | - |  |
| <1 unit a week | 3404 | 396 | 0.944 | 0.793,1.123 |  | 3191 | 375 | 0.953 | 0.726,1.252 |  | 2944 | 345 | 0.939 | 0.708,1.244 |  |
| 1-6 units a week | 4686 | 512 | 0.879 | 0.744,1.191 |  | 4362 | 473 | 0.838 | 0.639,1.299 |  | 3972 | 423 | 0.805 | 0.607,1.238 |  |
| >1 unit per day | 2828 | 345 | 0.996 | 0.833,1.805 |  | 2640 | 330 | 0.978 | 0.736,1.805 |  | 2360 | 289 | 0.920 | 0.684,1.805 |  |

Note: RS-DBD – the Disruptive Behaviour Disorders scale; N – sample size; n – number of cases; OR – odds ratio; 95% CI – 95% confidence intervals; *adjusted for child’s gender, birth year, parity, parental age, education, marital status, financial difficulties, depression and anxiety symptoms, prenatal smoking and caffeine consumption; **additionally adjusted for partner’s alcohol consumption

### Supplementary Table S32. Associations of maternal prenatal alcohol consumption on high risk of maternal reported offspring ADHD symptoms in MoBa (additionally adjusted for maternal ADHD)

|  |  |  | **Unadjusted model** | | |  |  | **Adjusted model** | | |  |  | **Mutually adjusted model** | | |
| --- | --- | --- | --- | --- | --- | --- | --- | --- | --- | --- | --- | --- | --- | --- | --- |
|  | **N** | **n** | **OR** | **95% CI** | **p-value** | **N** | **n** | **OR** | **95% CI** | **p-value** | **N** | **n** | **OR** | **95% CI** | **p-value** |
| **ADHD RS-DBD)** | 38,134 | 5030 |  |  | <0.001 | 28,507 | 3655 |  |  | <0.001 | 9031 | 1187 |  |  | 0.001 |
| None (ref) | 33,605 | 4342 | - | - |  | 25,110 | 3140 | - | - |  | 8317 | 1059 | - | - |  |
| <1 unit a week | 4366 | 663 | 1.207 | 1.104,1.319 |  | 3271 | 500 | 1.334 | 1.197,1.488 |  | 691 | 126 | 1.530 | 1.238,1.891 |  |
| >1 unit a week | 163 | 25 | 1.221 | 0.795,1.875 |  | 126 | 15 | 0.849 | 0.476,1.516 |  | NA | NA | NA | NA |  |
| **Hyperactive** | 38,127 | 4957 |  |  | <0.001 | 28,504 | 3600 |  |  | <0.001 | 9030 | 1149 |  |  | 0.007 |
| None (ref) | 33,601 | 4281 | - | - |  | 25,109 | 3097 | - | - |  | 8316 | 1031 | - | - |  |
| <1 unit a week | 4363 | 653 | 1.205 | 1.102,1.319 |  | 3269 | 489 | 1.287 | 1.154,1.435 |  | 691 | 114 | 1.377 | 1.105,1.716 |  |
| >1 unit a week | 163 | 23 | 1.125 | 0.722,1.754 |  | 126 | 14 | 0.780 | 0.436,1.395 |  | NA | NA | NA | NA |  |
| **Inattentive** | 38,140 | 4393 |  |  | <0.001 | 28,512 | 3186 |  |  | <0.001 | 9031 | 1026 |  |  | 0.009 |
| None (ref) | 33,610 | 3786 | - | - |  | 25,113 | 2747 | - | - |  | 8317 | 922 | - | - |  |
| <1 unit a week | 4367 | 584 | 1.216 | 1.106,1.337 |  | 3273 | 426 | 1.282 | 1.142,1.440 |  | 691 | 103 | 1.459 | 1.162,1.833 |  |
| >1 unit a week | 163 | 23 | 1.294 | 0.833,2.011 |  | 126 | 13 | 0.866 | 0.474,1.583 |  | NA | NA | NA | NA |  |

Note: RS-DBD – the Disruptive Behaviour Disorders scale; N – sample size; n – number of cases; OR – odds ratio; 95% CI – 95% confidence intervals; *adjusted model adjusted for child’s gender, birth year, parity, maternal age, education, marital status, financial difficulties, depression and anxiety symptoms, prenatal smoking and caffeine consumption; **additionally adjusted for partner’s alcohol consumption; NA – identifiability issue due to the low number of cases (<5 cases)

### Supplementary Table S33. Associations of maternal alcohol consumption before pregnancy on high risk of maternal reported offspring ADHD symptoms in MoBa

|  |  |  | **Unadjusted model** | | |  |  | **Adjusted model*** | | |  |  | **Mutually adjusted model**** | | |
| --- | --- | --- | --- | --- | --- | --- | --- | --- | --- | --- | --- | --- | --- | --- | --- |
|  | **N** | **n** | **OR** | **95% CI** | **p-value** | **N** | **n** | **OR** | **95% CI** | **p-value** | **N** | **n** | **OR** | **95% CI** | **p-value** |
| **ADHD (RS-DBD)** | 41020 | 5455 |  |  | <0.001 | 36636 | 4750 |  |  | 0.002 | 11302 | 1500 |  |  | 0.217 |
| None (ref) | 4363 | 537 | - | - |  | 3882 | 462 | - | - |  | 1608 | 194 | - | - |  |
| <1 unit a week | 15062 | 1825 | 0.982 | 0.886,1.089 |  | 13551 | 1595 | 1.011 | 0.901,1.133 |  | 3808 | 487 | 1.088 | 0.877,1.348 |  |
| 1-6 units a week | 18790 | 2602 | 1.145 | 1.036,1.728 |  | 16733 | 2273 | 1.081 | 0.964,1.500 |  | 4922 | 654 | 1.059 | 0.844,1.755 |  |
| >1 unit per day | 2805 | 491 | 1.512 | 1.322,1.805 |  | 2470 | 420 | 1.284 | 1.099,1.805 |  | 964 | 165 | 1.312 | 0.981,1.805 |  |
| **Hyperactive** | 41011 | 5381 |  |  | <0.001 | 36626 | 4693 |  |  | 0.004 | 11301 | 1458 |  |  | 0.069 |
| None (ref) | 4363 | 552 | - | - |  | 3882 | 477 | - | - |  | 1609 | 194 | - | - |  |
| <1 unit a week | 15059 | 1768 | 0.918 | 0.828,1.018 |  | 13548 | 1552 | 0.940 | 0.840,1.053 |  | 3808 | 464 | 1.030 | 0.829,1.279 |  |
| 1-6 units a week | 18785 | 2589 | 1.104 | 0.999,1.598 |  | 16728 | 2255 | 1.038 | 0.927,1.419 |  | 4921 | 635 | 1.051 | 0.836,1.867 |  |
| >1 unit per day | 2804 | 472 | 1.397 | 1.222,1.805 |  | 2468 | 409 | 1.215 | 1.040,1.805 |  | 963 | 165 | 1.394 | 1.041,1.805 |  |
| **Inattentive** | 41029 | 4775 |  |  | <0.001 | 36642 | 4169 |  |  | 0.115 | 11302 | 1307 |  |  | 0.629 |
| None (ref) | 4363 | 476 | - | - |  | 3881 | 415 | - | - |  | 1607 | 171 | - | - |  |
| <1 unit a week | 15065 | 1604 | 0.973 | 0.873,1.085 |  | 13554 | 1407 | 0.986 | 0.875,1.111 |  | 3808 | 421 | 1.092 | 0.866,1.376 |  |
| 1-6 units a week | 18790 | 2288 | 1.132 | 1.019,1.594 |  | 16731 | 2001 | 1.047 | 0.929,1.180 |  | 4922 | 591 | 1.108 | 0.869,1.472 |  |
| >1 unit per day | 2811 | 407 | 1.383 | 1.199,1.805 |  | 2476 | 346 | 1.215 | 1.040,1.805 |  | 965 | 124 | 1.073 | 0.783,1.805 |  |

Note: RS-DBD – the Disruptive Behaviour Disorders scale; N – sample size; n – number of cases; OR – odds ratio; 95% CI – 95% confidence intervals;*adjusted for child’s gender, birth year, parity, maternal age, education, marital status, financial difficulties, depression and anxiety symptoms, smoking and caffeine consumption before pregnancy; **additionally adjusted for partner’s alcohol consumption before partner’s pregnancy

### Supplementary Table S34. Associations of maternal alcohol consumption before pregnancy on high risk of maternal reported offspring ADHD symptoms in ALSPAC

|  | |  | |  | **Unadjusted model** | | |  |  | **Adjusted model*** | | |  |  | **Mutually adjusted model**** | | |
| --- | --- | --- | --- | --- | --- | --- | --- | --- | --- | --- | --- | --- | --- | --- | --- | --- | --- |
|  | **N** | | **n** | | **OR** | **95% CI** | **p-value** | **N** | **n** | **OR** | **95% CI** | **p-value** | **N** | **n** | **OR** | **95% CI** | **p-value** |
| **ADHD (DAWBA)** | 7722 | | 1117 | |  |  | 0.177 | 6639 | 931 |  |  | 0.087 | 5410 | 738 |  |  | 0.002 |
| None (ref) | 483 | | 91 | | - | - |  | 397 | 67 | - | - |  | 297 | 46 | - | - |  |
| <1 unit a week | 2897 | | 385 | | 0.660 | 0.513,0.849 |  | 2484 | 317 | 0.759 | 0.562,1.024 |  | 2027 | 249 | 0.871 | 0.603,1.260 |  |
| 1-6 units a week | 3446 | | 471 | | 0.682 | 0.532,0.874 |  | 3012 | 413 | 0.848 | 0.629,1.143 |  | 2486 | 340 | 1.119 | 0.769,1.628 |  |
| >1 unit a week | 896 | | 170 | | 1.009 | 0.760,1.338 |  | 746 | 134 | 1.080 | 0.764,1.525 |  | 600 | 103 | 1.450 | 0.937,2.243 |  |
| **Hyperactive** | 7743 | | 984 | |  |  | 0.664 | 6656 | 824 |  |  | 0.632 | 5420 | 645 |  |  | 0.023 |
| None (ref) | 485 | | 86 | | - | - |  | 398 | 65 | - | - |  | 298 | 41 | - | - |  |
| <1 unit a week | 2905 | | 355 | | 0.646 | 0.499,0.836 |  | 2491 | 299 | 0.751 | 0.555,1.016 |  | 2031 | 237 | 0.950 | 0.649,1.391 |  |
| 1-6 units a week | 3454 | | 404 | | 0.615 | 0.476,0.793 |  | 3018 | 347 | 0.756 | 0.558,1.023 |  | 2490 | 280 | 1.074 | 0.727,1.587 |  |
| >1 unit a week | 899 | | 139 | | 0.849 | 0.632,1.140 |  | 749 | 113 | 0.996 | 0.699,1.420 |  | 601 | 87 | 1.521 | 0.964,2.401 |  |
| **Inattentive** | 7734 | | 1060 | |  |  | 0.213 | 6653 | 885 |  |  | 0.168 | 5418 | 714 |  |  | 0.004 |
| None (ref) | 484 | | 85 | | - | - |  | 397 | 63 | - | - |  | 297 | 45 | - | - |  |
| <1 unit a week | 2900 | | 367 | | 0.680 | 0.525,0.881 |  | 2491 | 302 | 0.758 | 0.559,1.029 |  | 2032 | 236 | 0.845 | 0.583,1.225 |  |
| 1-6 units a week | 3459 | | 450 | | 0.702 | 0.544,0.905 |  | 3023 | 393 | 0.817 | 0.603,1.106 |  | 2492 | 331 | 1.080 | 0.740,1.576 |  |
| >1 unit a week | 891 | | 158 | | 1.012 | 0.757,1.353 |  | 742 | 127 | 1.046 | 0.736,1.486 |  | 597 | 102 | 1.408 | 0.908,2.184 |  |
| **ADHD (SDQ)** | 7990 | | 893 | |  |  | 0.815 | 6907 | 733 |  |  | 0.846 | 5627 | 580 |  |  | 0.530 |
| None (ref) | 515 | | 65 | | - | - |  | 419 | 49 | - | - |  | 319 | 31 | - | - |  |
| <1 unit a week | 2987 | | 343 | | 0.898 | 0.677,1.192 |  | 2588 | 286 | 1.028 | 0.737,1.433 |  | 2115 | 234 | 1.336 | 0.878,2.031 |  |
| 1-6 units a week | 3566 | | 363 | | 0.785 | 0.592,1.040 |  | 3130 | 306 | 0.930 | 0.666,1.299 |  | 2572 | 241 | 1.193 | 0.777,1.831 |  |
| >1 unit a week | 922 | | 122 | | 1.056 | 0.765,1.457 |  | 770 | 92 | 1.090 | 0.738,1.611 |  | 621 | 74 | 1.487 | 0.906,2.439 |  |

Note: DAWBA – Development and Well-Being Assessment; SDQ – Strengths and Difficulties Questionnaire; N – total sample size; n – number of ADHD cases; OR – odds ratio; 95% CI – 95% confidence intervals. *adjusted for child’s gender, ethnicity, parity, maternal age, marital status, education, financial difficulties, depression and anxiety symptoms, prenatal smoking and caffeine use; **additionally adjusted for partner’s alcohol use before pregnancy

### Supplementary Table S35. Associations of maternal and paternal prenatal alcohol consumption on high risk of teacher reported offspring ADHD symptoms in ALSPAC

|  |  |  | **Unadjusted model** | | |  | |  | **Adjusted model*** | | | | | |  | |  | **Mutually adjusted model**** | | | |
| --- | --- | --- | --- | --- | --- | --- | --- | --- | --- | --- | --- | --- | --- | --- | --- | --- | --- | --- | --- | --- | --- |
|  | **N** | **n** | **OR** | **95% CI** | **p-value** | **N** | | **n** | **OR** | | | **95% CI** | | **p-value** | **N** | | **n** | **OR** | | **95% CI** | **p-value** |
| **Maternal** | | | | | | | | | | | | | | | | | | | | | |
| **ADHD (DAWBA)** | 5736 | 806 |  |  | 0.353 | 4584 | | 576 |  | | |  | | 0.753 | 3565 | | 420 |  | |  | 0.941 |
| None (ref) | 2572 | 350 | - | - |  | 2044 | | 256 | - | | | - | |  | 1615 | | 184 | - | | - |  |
| <1 unit a week | 2262 | 323 | 1.058 | 0.898,1.245 |  | 1844 | | 234 | 1.069 | | | 0.875,1.307 | |  | 1433 | | 178 | 1.148 | | 0.910,1.449 |  |
| >1 unit a week | 902 | 133 | 1.098 | 0.885,1.362 |  | 696 | | 86 | 0.893 | | | 0.675,1.182 | |  | 517 | | 58 | 0.896 | | 0.638,1.259 |  |
| **Hyperactive** | 5735 | 725 |  |  | 0.053 | 4584 | | 520 |  | | |  | | 0.261 | 3565 | | 376 |  | |  | 0.228 |
| None (ref) | 2572 | 313 | - | - |  | 2044 | | 225 | - | | | - | |  | 1615 | | 160 | - | | - |  |
| <1 unit a week | 2261 | 275 | 0.999 | 0.841,1.188 |  | 1844 | | 203 | 1.032 | | | 0.835,1.274 | |  | 1433 | | 155 | 1.132 | | 0.885,1.448 |  |
| >1 unit a week | 902 | 137 | 1.293 | 1.041,1.606 |  | 696 | | 92 | 1.176 | | | 0.890,1.553 | |  | 517 | | 61 | 1.159 | | 0.824,1.630 |  |
| **Inattentive** | 5736 | 710 |  |  | 0.974 | 4583 | | 508 |  | | |  | | 0.312 | 3563 | | 370 |  | |  | 0.543 |
| None (ref) | 2571 | 321 | - | - |  | 2043 | | 232 | - | | | - | |  | 1614 | | 167 | - | | - |  |
| <1 unit a week | 2263 | 274 | 0.966 | 0.813,1.147 |  | 1844 | | 205 | 1.022 | | | 0.829,1.259 | |  | 1432 | | 154 | 1.074 | | 0.842,1.370 |  |
| >1 unit a week | 902 | 115 | 1.024 | 0.815,1.286 |  | 696 | | 71 | 0.800 | | | 0.593,1.078 | |  | 517 | | 49 | 0.807 | | 0.563,1.156 |  |
| **ADHD (SDQ)** | 5733 | 634 |  |  | 0.241 | 4587 | | 444 |  | | |  | | 0.915 | 3567 | | 321 |  | |  | 0.512 |
| None (ref) | 2576 | 272 | - | - |  | 2048 | | 192 | - | | | - | |  | 1618 | | 130 | - | | - |  |
| <1 unit a week | 2255 | 255 | 1.080 | 0.901,1.294 |  | 1843 | | 190 | 1.206 | | | 0.965,1.507 | |  | 1432 | | 147 | 1.368 | | 1.053,1.776 |  |
| >1 unit a week | 902 | 107 | 1.140 | 0.899,1.446 |  | 696 | | 62 | 0.864 | | | 0.628,1.190 | |  | 517 | | 44 | 0.931 | | 0.634,1.368 |  |
| **Paternal** | | | | | | | | | | | | | | | | | | | | | |
| **ADHD (DAWBA)** | 4212 | 538 |  |  | 0.103 | 3075 | 334 | | |  |  | | 0.854 | | 3067 | 334 | | |  |  | 0.787 |
| None (ref) | 155 | 22 | - | - |  | 84 | 11 | | | - | - | |  | | 84 | 11 | | | - | - |  |
| <1 unit a week | 1022 | 143 | 0.984 | 0.606,1.597 |  | 724 | 81 | | | 0.960 | 0.472,1.954 | |  | | 722 | 81 | | | 0.905 | 0.442,1.852 |  |
| 1-6 units a week | 2231 | 280 | 0.868 | 0.543,1.386 |  | 1661 | 181 | | | 1.071 | 0.537,2.136 | |  | | 1658 | 181 | | | 1.003 | 0.499,2.019 |  |
| >1 unit a day | 804 | 93 | 0.791 | 0.480,1.304 |  | 606 | 61 | | | 0.906 | 0.438,1.875 | |  | | 603 | 61 | | | 0.853 | 0.408,1.784 |  |
| **Hyperactive** | 4211 | 483 |  |  | 0.353 | 3075 | 334 | | |  |  | | 0.506 | | 3067 | 313 | | |  |  | 0.341 |
| None (ref) | 155 | 16 | - | - |  | 84 | 11 | | | - | - | |  | | 84 | 9 | | | - | - |  |
| <1 unit a week | 1022 | 129 | 1.255 | 0.725,2.174 |  | 724 | 81 | | | 1.196 | 0.556,2.569 | |  | | 722 | 81 | | | 1.134 | 0.526,2.447 |  |
| 1-6 units a week | 2230 | 252 | 1.107 | 0.649,1.887 |  | 1661 | 181 | | | 1.152 | 0.546,2.432 | |  | | 1658 | 166 | | | 1.068 | 0.502,2.272 |  |
| >1 unit a day | 804 | 86 | 1.041 | 0.592,1.828 |  | 606 | 61 | | | 0.997 | 0.456,2.183 | |  | | 603 | 57 | | | 0.905 | 0.409,2.000 |  |
| **Inattentive** | 4210 | 471 |  |  | 0.442 | 3073 | 307 | | |  |  | | 0.529 | | 3065 | 307 | | |  |  | 0.562 |
| None (ref) | 155 | 22 | - | - |  | 84 | 10 | | | - | - | |  | | 84 | 10 | | | - | - |  |
| <1 unit a week | 1022 | 124 | 0.835 | 0.512,1.361 |  | 724 | 76 | | | 1.006 | 0.485,2.085 | |  | | 722 | 76 | | | 0.953 | 0.457,1.986 |  |
| 1-6 units a week | 2230 | 228 | 0.688 | 0.430,1.103 |  | 1660 | 151 | | | 0.946 | 0.464,1.925 | |  | | 1657 | 151 | | | 0.893 | 0.435,1.833 |  |
| >1 unit a day | 803 | 97 | 0.831 | 0.505,1.367 |  | 605 | 70 | | | 1.156 | 0.552,2.421 | |  | | 602 | 70 | | | 1.101 | 0.520,2.329 |  |
| **ADHD (SDQ)** | 4209 | 416 |  |  | 0.190 | 3079 | 261 | | |  |  | | 0.991 | | 3071 | 261 | | |  |  | 0.793 |
| None (ref) | 155 | 18 |  |  |  | 84 | 9 | | |  |  | |  | | 84 | 9 | | |  |  |  |
| <1 unit a week | 1021 | 109 | 0.910 | 0.535,1.545 |  | 725 | 63 | | | 0.934 | 0.432,2.021 | |  | | 723 | 63 | | | 0.824 | 0.378,1.797 |  |
| 1-6 units a week | 2227 | 215 | 0.813 | 0.488,1.356 |  | 1662 | 137 | | | 0.992 | 0.469,2.097 | |  | | 1659 | 137 | | | 0.858 | 0.401,1.836 |  |
| >1 unit a day | 806 | 74 | 0.769 | 0.445,1.329 |  | 608 | 52 | | | 0.936 | 0.426,2.056 | |  | | 605 | 52 | | | 0.809 | 0.363,1.799 |  |

Note: DAWBA – Development and Well-Being Assessment; SDQ – Strengths and Difficulties Questionnaire; N – total sample size; n – number of ADHD cases; OR – odds ratio; 95% CI – 95% confidence intervals; *adjusted for child’s gender, ethnicity, parity, parental age, marital status, education, financial difficulties, depression and anxiety symptoms, prenatal smoking and caffeine use; **additionally adjusted for partner’s prenatal alcohol use

### Supplementary Table S36. Associations of maternal prenatal weekly alcohol consumption in grams on high risk of maternal reported offspring ADHD symptoms in ALSPAC

|  |  |  | **Unadjusted model** | | |  |  | **Adjusted model*** | | |  |  | **Mutually adjusted model**** | | |
| --- | --- | --- | --- | --- | --- | --- | --- | --- | --- | --- | --- | --- | --- | --- | --- |
|  | N | **n** | **OR** | **95% CI** | **p-value** | **N** | **n** | **OR** | **95% CI** | **p-value** | **N** | **n** | **OR** | **95% CI** | **p-value** |
| **ADHD (DAWBA)** | 7180 | 1029 | 1.003 | 1.000,1.004 | 0.003 | 6671 | 933 | 1.001 | 0.999,1.003 | 0.263 | 5381 | 732 | 1.001 | 0.999,1.003 | 0.352 |
| **Hyperactive** | 7200 | 911 | 1.002 | 1.000,1.004 | 0.007 | 6689 | 828 | 1.001 | 0.999,1.003 | 0.170 | 5391 | 642 | 1.001 | 0.999,1.004 | 0.240 |
| **Inattentive** | 7192 | 987 | 1.003 | 1.001,1.004 | 0.004 | 6684 | 889 | 1.001 | 0.999,1.003 | 0.168 | 5389 | 707 | 1.002 | 0.999,1.004 | 0.126 |
| **ADHD (SDQ)** | 7442 | 809 | 1.001 | 0.999,1.003 | 0.325 | 6942 | 737 | 0.999 | 0.997,1.002 | 0.669 | 5610 | 579 | 0.999 | 0.997,1.002 | 0.804 |

Note: DAWBA – Development and Well-Being Assessment; SDQ – Strengths and Difficulties Questionnaire; N – total sample size; n – number of ADHD cases; OR – odds ratio; 95% CI – 95% confidence intervals; *adjusted for child’s gender, ethnicity, parity, parental age, marital status, education, financial difficulties, depression and anxiety symptoms, prenatal smoking and caffeine use; **additionally adjusted for partner’s prenatal alcohol use

### Supplementary Table S37. Associations of maternal prenatal weekly alcohol consumption in grams on high risk of maternal reported offspring ADHD symptoms in MoBa

|  |  |  | **Unadjusted model** | | |  |  | **Adjusted model*** | | |  |  | **Mutually adjusted model**** | | |
| --- | --- | --- | --- | --- | --- | --- | --- | --- | --- | --- | --- | --- | --- | --- | --- |
|  | **N** | **n** | **OR** | **95% CI** | **p-value** | **N** | **n** | **OR** | **95% CI** | **p-value** | **N** | **n** | **OR** | **95% CI** | **p-value** |
| **ADHD (RS-DBD)** | 38,134 | 5,030 | 1.011 | 1.000,1.021 | 0.041 | 34,297 | 4,415 | 1.009 | 0.999,1.020 | 0.085 | 10,641 | 1,395 | 1.002 | 0.989,1.015 | 0.789 |
| **Hyperactive** | 38,127 | 4,957 | 1.009 | 0.999,1.019 | 0.074 | 34,290 | 4,353 | 1.006 | 0.996,1.015 | 0.231 | 10,638 | 1,351 | 0.999 | 0.985,1.014 | 0.969 |
| **Inattentive** | 38,140 | 4,393 | 1.011 | 1.001,1.021 | 0.038 | 34,302 | 3,874 | 1.009 | 0.999,1.019 | 0.091 | 10,639 | 1,210 | 1.001 | 0.987,1.015 | 0.920 |

Note: RS-DBD – the Disruptive Behaviour Disorders scale; N – sample size; n – number of cases; OR – odds ratio; 95% CI – 95% confidence intervals;*adjusted for child’s gender, birth year, parity, maternal age, education, marital status, financial difficulties, depression and anxiety symptoms, smoking and caffeine consumption before pregnancy; **additionally adjusted for partner’s alcohol consumption before partner’s pregnancy

### Supplementary Table S38. Associations of maternal and paternal prenatal alcohol consumption on high risk of maternal reported offspring ADHD symptoms in ALSPAC (complete cases)

|  | |  |  | **Unadjusted model** | | | | | **Adjusted model*** | | | **Mutually adjusted model**** | | | | |
| --- | --- | --- | --- | --- | --- | --- | --- | --- | --- | --- | --- | --- | --- | --- | --- | --- |
|  | | **N** | **n** | **OR** | | **95% CI** | | **p-value** | **OR** | **95% CI** | **p-value** | **OR** | | **95% CI** | **p-value** | |
| **Maternal** | | | | | | | | | | | | | | | | |
| **ADHD (DAWBA)** | | 5384 | 733 |  | |  | | 0.001 |  |  | 0.006 |  | |  | 0.002 | |
| None (ref) | | 2369 | 288 | - | | - | |  | - | - |  | - | | - |  | |
| <1 unit a week | | 2220 | 311 | 1.177 | | 0.991,1.398 | |  | 1.198 | 1.002,1.432 |  | 1.229 | | 1.026,1.471 |  | |
| >1 unit a week | | 795 | 134 | 1.465 | | 1.172,1.830 | |  | 1.368 | 1.079,1.733 |  | 1.437 | | 1.128,1.832 |  | |
| **Hyperactive** | | 5394 | 642 |  | |  | | 0.023 |  |  | 0.045 |  | |  | 0.009 | |
| None (ref) | | 2378 | 260 | - | | - | |  | - | - |  | - | | - |  | |
| <1 unit a week | | 2221 | 272 | 1.137 | | 0.949,1.362 | |  | 1.157 | 0.961,1.394 |  | 1.196 | | 0.991,1.443 |  | |
| >1 unit a week | | 795 | 110 | 1.308 | | 1.030,1.661 | |  | 1.261 | 0.981,1.620 |  | 1.377 | | 1.064,1.781 |  | |
| **Inattentive** | | 5392 | 708 |  | |  | | 0.001 |  |  | 0.007 |  | |  | 0.004 | |
| None (ref) | | 2375 | 279 | - | | - | |  | - | - |  | - | | - |  | |
| <1 unit a week | | 2222 | 301 | 1.177 | | 0.989,1.401 | |  | 1.206 | 1.007,1.444 |  | 1.196 | | 0.991,1.443 |  | |
| >1 unit a week | | 795 | 128 | 1.442 | | 1.149,1.808 | |  | 1.366 | 1.075,1.736 |  | 1.377 | | 1.064,1.781 |  | |
| **ADHD (SDQ)** | | 5613 | 579 |  | |  | | 0.004 |  |  | 0.020 |  | |  | 0.008 | |
| None (ref) | | 2471 | 222 | - | | - | |  | - | - |  | - | | - |  | |
| <1 unit a week | | 2310 | 257 | 1.268 | | 1.049,1.532 | |  | 1.290 | 1.062,1.568 |  | 1.196 | | 0.991,1.443 |  | |
| >1 unit a week | | 832 | 100 | 1.384 | | 1.077,1.778 | |  | 1.296 | 0.997,1.684 |  | 1.377 | | 1.064,1.781 |  | |
| **Paternal** | | | | | | | | | | | | | | | | |
| **ADHD (DAWBA)** | 4648 | | 622 | |  |  | 0.096 | |  |  | 0.204 | |  |  | | 0.048 |
| None (ref) | 148 | | 36 | | - | - |  | | - | - |  | | - | - | |  |
| <1 unit a week | 1053 | | 136 | | 0.461 | 0.304,0.700 |  | | 0.485 | 0.314,0.749 |  | | 0.452 | 0.292,0.701 | |  |
| 1-6 units a week | 2465 | | 320 | | 0.464 | 0.313,0.688 |  | | 0.528 | 0.350,0.798 |  | | 0.471 | 0.310,0.715 | |  |
| >1 unit a day | 982 | | 130 | | 0.475 | 0.312,0.721 |  | | 0.508 | 0.327,0.788 |  | | 0.436 | 0.279,0.683 | |  |
| **Hyperactive** | 4657 | | 535 | |  |  | 0.003 | |  |  | 0.018 | |  |  | | 0.004 |
| None (ref) | 148 | | 30 | | - | - |  | | - | - |  | | - | - | |  |
| <1 unit a week | 1059 | | 131 | | 0.555 | 0.357,0.863 |  | | 0.605 | 0.384,0.954 |  | | 0.563 | 0.350,0.905 | |  |
| 1-6 units a week | 2470 | | 275 | | 0.493 | 0.324,0.750 |  | | 0.581 | 0.376,0.898 |  | | 0.541 | 0.343,0.855 | |  |
| >1 unit a day | 980 | | 99 | | 0.442 | 0.281,0.694 |  | | 0.499 | 0.312,0.798 |  | | 0.471 | 0.288,0.771 | |  |
| **Inattentive** | 4657 | | 614 | |  |  | 0.253 | |  |  | 0.363 | |  |  | | 0.102 |
| None (ref) | 148 | | 32 | | - | - |  | | - | - |  | | - | - | |  |
| <1 unit a week | 1056 | | 141 | | 0.559 | 0.364,0.858 |  | | 0.592 | 0.379,0.923 |  | | 0.505 | 0.320,0.796 | |  |
| 1-6 units a week | 2467 | | 304 | | 0.509 | 0.338,0.767 |  | | 0.563 | 0.368,0.862 |  | | 0.451 | 0.291,0.699 | |  |
| >1 unit a day | 986 | | 137 | | 0.585 | 0.380,0.900 |  | | 0.622 | 0.397,0.975 |  | | 0.491 | 0.308,0.781 | |  |
| **ADHD (SDQ)** | 4796 | | 487 | |  |  | 0.300 | |  |  | 0.447 | |  |  | | 0.156 |
| None (ref) | 160 | | 24 | | - | - |  | | - | - |  | | - | - | |  |
| <1 glass a week | 1095 | | 114 | | 0.659 | 0.409,1.059 |  | | 0.682 | 0.418,1.111 |  | | 0.620 | 0.374,1.027 | |  |
| 1-6 units a week | 2537 | | 245 | | 0.606 | 0.385,0.953 |  | | 0.666 | 0.417,1.064 |  | | 0.579 | 0.356,0.941 | |  |
| >1 unit a day | 1004 | | 104 | | 0.655 | 0.406,1.057 |  | | 0.688 | 0.419,1.131 |  | | 0.552 | 0.328,0.928 | |  |

Note: DAWBA – Development and Well-Being Assessment; SDQ – Strengths and Difficulties Questionnaire; N – total sample size; n – number of ADHD cases; OR – odds ratio; 95% CI – 95% confidence intervals; *adjusted for child’s gender, ethnicity, parity, parental age, marital status, education, financial difficulties, depression and anxiety symptoms, prenatal smoking and caffeine use; **additionally adjusted for partner’s prenatal alcohol use

### Supplementary Table S39. Associations of maternal and paternal prenatal alcohol consumption on high risk of maternal and teacher reported offspring ADHD symptoms in GenR (complete cases)

|  |  | |  | | **Unadjusted model** | | | | | | **Adjusted model*** | | | | | | **Mutually adjusted model**** | | | | | |
| --- | --- | --- | --- | --- | --- | --- | --- | --- | --- | --- | --- | --- | --- | --- | --- | --- | --- | --- | --- | --- | --- | --- |
|  | **N** | | **n** | | **OR** | | **95% CI** | | **p-value** | | **OR** | | **95% CI** | | **p-value** | | **OR** | | **95% CI** | | **p-value** | |
| **Maternal** | | | | | | | | | | | | | | | | | | | | | | |
| **ADHD (CPRS-R)** | 1532 | | 196 | |  | |  | | 0.292 | |  | |  | | 0.353 | |  | |  | | 0.581 | |
| None (ref) | 630 | | 72 | | - | | - | |  | | - | | - | |  | | - | | - | |  | |
| <1 unit a week | 507 | | 71 | | 1.262 | | 0.889,1.793 | |  | | 1.307 | | 0.898,1.902 | |  | | 1.251 | | 0.854,1.834 | |  | |
| >1 unit a week | 395 | | 53 | | 1.201 | | 0.822,1.755 | |  | | 1.201 | | 0.788,1.831 | |  | | 1.118 | | 0.720,1.735 | |  | |
| **Hyperactive** | 1533 | | 144 | |  | |  | | 0.285 | | - | |  | | 0.284 | |  | |  | | 0.164 | |
| None (ref) | 631 | | 66 | | - | | - | |  | |  | | - | |  | | - | | - | |  | |
| <1 unit a week | 507 | | 44 | | 0.814 | | 0.545,1.215 | |  | | 0.828 | | 0.541,1.265 | |  | | 0.784 | | 0.509,1.207 | |  | |
| >1 unit a week | 395 | | 34 | | 0.806 | | 0.522,1.245 | |  | | 0.779 | | 0.483,1.257 | |  | | 0.712 | | 0.433,1.169 | |  | |
| **Inattentive** | 1534 | | 170 | |  | |  | | 0.149 | | - | | - | | 0.242 | | - | | - | | 0.498 | |
| None (ref) | 632 | | 60 | | - | | - | |  | |  | |  | |  | |  | |  | |  | |
| <1 unit a week | 507 | | 62 | | 1.328 | | 0.912,1.934 | |  | | 1.354 | | 0.909,2.016 | |  | | 1.276 | | 0.851,1.913 | |  | |
| >1 unit a week | 395 | | 48 | | 1.319 | | 0.882,1.972 | |  | | 1.285 | | 0.824,2.003 | |  | | 1.161 | | 0.732,1.842 | |  | |
| **ADHD (CBCL)** | 1828 | | 212 | |  | |  | | 0.180 | | - | | - | | 0.166 | | - | | - | | 0.105 | |
| None (ref) | 809 | | 101 | | - | | - | |  | |  | |  | |  | |  | |  | |  | |
| <1 unit a week | 574 | | 67 | | 0.926 | | 0.667,1.288 | |  | | 0.979 | | 0.685,1.400 | |  | | 0.940 | | 0.651,1.356 | |  | |
| >1 unit a week | 445 | | 44 | | 0.769 | | 0.529,1.119 | |  | | 0.727 | | 0.476,1.108 | |  | | 0.684 | | 0.441,1.062 | |  | |
| **ADHD (TRF)** | 1148 | | 126 | |  | |  | | 0.059 | | - | | - | | 0.267 | | - | | - | | 0.178 | |
| None (ref) | 539 | | 72 | | - | | - | |  | |  | |  | |  | |  | |  | |  | |
| <1 unit a week | 342 | | 28 | | 0.578 | | 0.365,0.916 | |  | | 0.643 | | 0.387,1.068 | |  | | 0.601 | | 0.357,1.013 | |  | |
| >1 unit a week | 267 | | 26 | | 0.700 | | 0.435,1.125 | |  | | 0.784 | | 0.454,1.352 | |  | | 0.715 | | 0.404,1.264 | |  | |
| **Paternal** | | | | | | | | | | | | | | | | | | | | | | |
| **ADHD (CPRS-R)** | | 1937 | | 261 | |  | |  | | 0.188 | |  | |  | | 0.114 | |  | |  | | 0.163 |
| None (ref) | | 201 | | 19 | | - | | - | |  | | - | | - | |  | | - | | - | |  |
| <1 unit a week | | 238 | | 35 | | 1.652 | | 0.913,2.989 | |  | | 1.642 | | 0.881,3.061 | |  | | 1.575 | | 0.841,2.948 | |  |
| 1-6 units a week  >1 unit a day | | 982  516 | | 133  74 | | 1.501  1.604 | | 0.904,2.491  0.941,2.732 | |  | | 1.638  1.778 | | 0.940,2.855  0.982,3.220 | |  | | 1.555  1.700 | | 0.880,2.746  0.921,3.139 | |  |
| **Hyperactive** | | 1937 | | 186 | |  | |  | | 0.869 | |  | |  | | 0.748 | |  | |  | | 0.401 |
| None (ref) | | 201 | | 17 | | - | | - | |  | | - | | - | |  | | - | | - | |  |
| <1 unit a week | | 238 | | 31 | | 1.621 | | 0.869,3.025 | |  | | 1.617 | | 0.835,3.130 | |  | | 1.683 | | 0.866,3.270 | |  |
| 1-6 units a week  >1 unit a day | | 982  516 | | 83  55 | | 0.999  1.291 | | 0.579,1.724  0.730,2.284 | |  | | 1.061  1.349 | | 0.580,1.940  0.709,2.567 | |  | | 1.180  1.568 | | 0.637,2.185  0.808,3.045 | |  |
| **Inattentive** | | 1939 | | 234 | |  | |  | | 0.125 | |  | |  | | 0.120 | |  | |  | | 0.144 |
| None (ref) | | 201 | | 18 | | - | | - | |  | | - | | - | |  | | - | | - | |  |
| <1 unit a week | | 238 | | 31 | | 1.523 | | 0.824,2.813 | |  | | 1.415 | | 0.743,2.693 | |  | | 1.375 | | 0.719,2.629 | |  |
| 1-6 units a week  >1 unit a day | | 982  518 | | 113  72 | | 1.322  1.641 | | 0.784,2.229  0.952,2.829 | |  | | 1.323  1.662 | | 0.746,2.345  0.907,3.049 | |  | | 1.283  1.627 | | 0.714,2.306  0.869,3.045 | |  |
| **ADHD (CBCL)** | | 2332 | | 278 | |  | |  | | 0.158 | |  | |  | | 0.637 | |  | |  | | 0.865 |
| None (ref) | | 299 | | 39 | | - | | - | |  | | - | | - | |  | | - | | - | |  |
| <1 unit a week | | 317 | | 48 | | 1.190 | | 0.754,1.876 | |  | | 1.313 | | 0.810,2.129 | |  | | 1.382 | | 0.848,2.251 | |  |
| 1-6 units a week  >1 unit a day | | 1139  577 | | 126  65 | | 0.829  0.846 | | 0.565,1.218  0.554,1.293 | |  | | 0.976  1.013 | | 0.633,1.504  0.621,1.653 | |  | | 1.093  1.173 | | 0.699,1.710  0.704,1.953 | |  |
| **ADHD (TRF)** | | 1452 | | 149 | |  | |  | | 0.247 | |  | |  | | 0.977 | |  | |  | | 0.680 |
| None (ref) | | 213 | | 28 | | - | | - | |  | | - | | - | |  | | - | | - | |  |
| <1 unit a week | | 196 | | 18 | | 0.668 | | 0.357,1.251 | |  | | 0.786 | | 0.392,1.576 | |  | | 0.851 | | 0.422,1.713 | |  |
| 1-6 units a week  >1 unit a day | | 695  348 | | 70  33 | | 0.740  0.692 | | 0.463,1.182  0.405,1.182 | |  | | 0.984  0.908 | | 0.565,1.713  0.476,1.731 | |  | | 1.145  1.050 | | 0.643,2.039  0.534,2.064 | |  |

Note: CPRS-R – The revised Conners’ Parent Rating Scale; CBCL – Child Behaviour Checklist; TRF – Teacher’s Report Form; N – sample size; n – number of cases; OR – odds ratio; 95% CI – 95% confidence intervals; *adjusted for child’s gender, parity, parental ethnicity, age, education, anxiety and depression problems, financial difficulties, parental smoking and prenatal caffeine use in the maternal model; **additionally adjusted for partner’s alcohol use

### Supplementary Table S40. Associations of maternal and paternal prenatal alcohol consumption on high risk of maternal reported offspring ADHD symptoms in MoBa (complete cases)

|  |  | |  | **Unadjusted model** | | | | | **Adjusted model*** | | | | | **Mutually adjusted model**** | | |
| --- | --- | --- | --- | --- | --- | --- | --- | --- | --- | --- | --- | --- | --- | --- | --- | --- |
|  | **N** | | **n** | **OR** | | **95% CI** | | **p-value** | **OR** | | **95% CI** | **p-value** | | **OR** | **95% CI** | **p-value** |
| **Maternal** | | | | | | | | | | | | | | | | |
| **ADHD (RS-DBD)** | 10,641 | | 1395 |  | |  | | <0.001 |  | |  | <0.001 | |  |  | <0.001 |
| None (ref) | 9800 | | 1247 | - | | - | |  | - | | - |  | | - | - |  |
| <1 unit a week | 811 | | 146 | 1.506 | | 1.246,1.820 | |  | 1.529 | | 1.258,1.857 |  | | 1.533 | 1.260,1.865 |  |
| >1 unit a week | 30 | | 2 | 0.490 | | 0.117,2.059 | |  | 0.377 | | 0.087,1.633 |  | | 0.374 | 0.085,1.647 |  |
| **Hyperactive** | 10,638 | | 1351 |  | |  | | 0.002 |  | |  | 0.006 | |  |  | 0.005 |
| None (ref) | 9797 | | 1214 | - | | - | |  | - | | - |  | | - | - |  |
| <1 unit a week | 811 | | 133 | 1.387 | | 1.138,1.690 | |  | 1.381 | | 1.129,1.690 |  | | 1.395 | 1.139,1.710 |  |
| >1 unit a week | 30 | | 4 | 1.088 | | 0.379,3.122 | |  | 0.887 | | 0.285,2.759 |  | | 0.890 | 0.283,2.796 |  |
| **Inattentive** | 10,639 | | 1210 |  | |  | | 0.003 |  | |  | 0.003 | |  |  | 0.002 |
| None (ref) | 9799 | | 1087 | - | | - | |  | - | | - |  | | - | - |  |
| <1 unit a week | 810 | | 120 | 1.394 | | 1.138,1.708 | |  | 1.437 | | 1.166,1.771 |  | | 1.452 | 1.176,1.793 |  |
| >1 unit a week | 30 | | 3 | 0.891 | | 0.270,2.941 | |  | 0.742 | | 0.202,2.730 |  | | 0.740 | 0.197,2.777 |  |
| **Paternal** | | | | | | | | | | | | | | | | |
| **ADHD (RS-DBD)** | | 9861 | 1322 | |  | |  | 0.875 | |  |  | | 0.666 |  |  | 0.366 |
| None (ref) | | 586 | 82 | | - | | - |  | | - | - | |  | - | - |  |
| <1 unit a week | | 2945 | 403 | | 0.974 | | 0.753,1.261 |  | | 0.995 | 0.761,1.301 | |  | 0.971 | 0.742,1.270 |  |
| 1-6 units a week | | 3971 | 501 | | 0.887 | | 0.689,1.327 |  | | 0.880 | 0.674,1.354 | |  | 0.850 | 0.651,1.284 |  |
| >1 units per day | | 2359 | 336 | | 1.021 | | 0.786,1.509 |  | | 1.023 | 0.773,1.509 | |  | 0.969 | 0.731,1.509 |  |
| **Hyperactive** | | 9858 | 1294 | |  | |  | 0.929 | |  |  | | 0.950 |  |  | 0.616 |
| None (ref) | | 586 | 86 | | - | | - |  | | - | - | |  | - | - |  |
| <1 unit a week | | 2944 | 387 | | 0.880 | | 0.684,1.132 |  | | 0.897 | 0.691,1.163 | |  | 0.877 | 0.676,1.137 |  |
| 1-6 units a week | | 3969 | 490 | | 0.819 | | 0.640,1.226 |  | | 0.819 | 0.632,1.268 | |  | 0.793 | 0.611,1.206 |  |
| >1 units per day | | 2359 | 331 | | 0.949 | | 0.735,1.509 |  | | 0.966 | 0.736,1.509 | |  | 0.917 | 0.698,1.509 |  |
| **Inattentive** | | 9862 | 1129 | |  | |  | 0.971 | |  |  | | 0.524 |  |  | 0.293 |
| None (ref) | | 586 | 72 | | - | | - |  | | - | - | |  | - | - |  |
| <1 unit a week | | 2944 | 345 | | 0.948 | | 0.721,1.245 |  | | 0.960 | 0.724,1.272 | |  | 0.939 | 0.708,1.244 |  |
| 1-6 units a week | | 3972 | 423 | | 0.851 | | 0.651,1.314 |  | | 0.832 | 0.628,1.303 | |  | 0.805 | 0.607,1.238 |  |
| >1 units per day | | 2360 | 289 | | 0.996 | | 0.755,1.509 |  | | 0.970 | 0.722,1.509 | |  | 0.920 | 0.684,1.509 |  |

Note: RS-DBD – the Disruptive Behaviour Disorders scale; N – sample size; n – number of cases; OR – odds ratio; 95% CI – 95% confidence intervals; *adjusted for child’s gender, birth year, parity, parental age, education, marital status, financial difficulties, depression and anxiety symptoms, prenatal smoking and caffeine use; **additionally adjusted for partner’s alcohol consumption

### Supplementary Table S41. Associations of maternal alcohol PRS on confounders in ALSPAC

| **Confounder** | **Effect estimate** | **Effect size** | **95% CI** | **P-value** | **Sample size** |
| --- | --- | --- | --- | --- | --- |
| Maternal age | beta | 0.462 | -1.241, 2.165 | 0.595 | 6944 |
| Maternal education | beta | 0.520 | 0.058, 0.983 | 0.027 | 6438 |
| Financial difficulties | beta | 1.091 | -0.186, 2.368 | 0.094 | 6277 |
| Marital status | OR | 0.441 | 0.182, 1.068 | 0.070 | 6660 |
| Depression symptoms | OR | 3.418 | 1.058, 11.047 | 0.040 | 6262 |
| Anxiety symptoms | OR | 1.850 | 0.639, 5.362 | 0.257 | 6229 |
| Parity | beta | 0.086 | -0.228, 0.401 | 0.590 | 6574 |

Note: adjusted for principal components; OR – odds ratio; 95% CI – 95% confidence intervals and excluding mothers who did not report drinking before pregnancy

### Supplementary Table S42. Associations of maternal alcohol PRS on confounders in MoBa

| **Confounder** | **Effect estimate** | **Effect size** | **95% CI** | **P-value** | **Sample size** |
| --- | --- | --- | --- | --- | --- |
| Maternal age | beta | -1.766 | -4.678, 1.146 | 0.235 | 13,216 |
| Maternal education | beta | -0.069 | -0.199 0.060 | 0.292 | 12,524 |
| Financial difficulties | OR | 0.830 | 0.403, 1.049 | 0.690 | 12,228 |
| Marital status | beta | -0.044 | -0.097, 0.009 | 0.102 | 13,152 |
| Depr/Anxiety symptoms | OR | 0.616 | 0.230, 1.652 | 0.336 | 13,109 |
| Maternal ADHD | OR | 2.320 | 0.327, 16.477 | 0.400 | 7985 |
| Parity | beta | 0.017 | -0.188, 0.222 | 0.870 | 13,216 |

Note: adjusted for principal components, birth year, genotyping batch; OR – odds ratio; 95% CI – 95% confidence intervals and excluding mothers who did not report drinking before pregnancy

### Supplementary Table S43. Associations of maternal alcohol consumption PRS on high risk of maternal reported offspring ADHD symptoms in ALSPAC

|  | **Maternal PRS** | | | **Maternal PRS adj. for PC** | | |  |
| --- | --- | --- | --- | --- | --- | --- | --- |
| **Outcome** | **OR** | **95% CI** | **P-value** | **OR** | **95% CI** | **P-value** | **Sample size** |
| ADHD (DAWBA) | 2.006 | 0.422, 9.542 | 0.382 | 2.143 | 0.449, 10.218 | 0.339 | 2890 |
| Hyperactive | 0.461 | 0.088, 2.429 | 0.361 | 0.494 | 0.094, 2.609 | 0.406 | 2890 |
| Inattentive | 1.213 | 0.249, 5.908 | 0.811 | 1.268 | 0.260, 6.182 | 0.769 | 2893 |
| ADHD (SDQ) | 1.525 | 0.272, 8.566 | 0.632 | 1.453 | 0.259, 8.164 | 0.671 | 2934 |

Note: DAWBA – Development and Well-Being Assessment; SDQ – Strengths and Difficulties Questionnaire; OR – odds ratio; 95% CI – 95% confidence intervals; PC – principal components

### Supplementary Table S44. Associations of maternal alcohol consumption PRS on high risk of teacher reported offspring ADHD symptoms in ALSPAC

|  | **Maternal PRS** | | | **Maternal PRS adj. for PC** | | |  |
| --- | --- | --- | --- | --- | --- | --- | --- |
| **Outcome** | **OR** | **95% CI** | **P-value** | **OR** | **95% CI** | **P-value** | **Sample size** |
| ADHD (DAWBA) | 1.410 | 0.197, 10.082 | 0.732 | 1.614 | 0.222, 11.715 | 0.636 | 2022 |
| Hyperactive | 2.142 | 0.284, 16.174 | 0.460 | 2.443 | 0.316, 18.910 | 0.392 | 2021 |
| Inattentive | 1.367 | 0.165, 11.347 | 0.772 | 1.666 | 0.197, 14.125 | 0.640 | 2023 |
| ADHD (SDQ) | 2.220 | 0.259, 19.033 | 0.467 | 2.627 | 0.300, 22.989 | 0.383 | 2018 |

Note: DAWBA – Development and Well-Being Assessment; SDQ – Strengths and Difficulties Questionnaire; OR – odds ratio; 95% CI – 95% confidence intervals; PC – principal components

### Supplementary Table S45. Associations of maternal alcohol consumption PRS on high risk of maternal reported offspring ADHD symptoms in MoBa

|  | **Maternal PRS** | | | **Maternal PRS adj. for PC** | | |  |
| --- | --- | --- | --- | --- | --- | --- | --- |
| **Outcome** | **OR** | **95% CI** | **P-value** | **OR** | **95% CI** | **P-value** | **Sample size** |
| ADHD (RS-DBD) | 0.659 | 0.064, 6.837 | 0.727 | 0.640 | 0.060, 6.769 | 0.710 | 1356 |
| Hyperactive | 0.759 | 0.077, 7.455 | 0.813 | 0.832 | 0.083, 8.374 | 0.876 | 1355 |
| Inattentive | 0.951 | 0.086, 10.479 | 0.967 | 0.989 | 0.089, 11.048 | 0.993 | 1358 |

Note: RS-DBD – The Disruptive Behavior Disorders Scale; OR – odds ratio; 95% CI – 95% confidence intervals; adjusted for principal components (PC), birth year and genotyping batch

### Supplementary Table S46. Associations of maternal and paternal daily prenatal caffeine consumption on high risk of maternal reported offspring ADHD symptoms in MoBa

|  |  |  | **Unadjusted model** | | | |  | |  | **Adjusted model*** | | | |  |  | **Mutually adjusted model**** | | |
| --- | --- | --- | --- | --- | --- | --- | --- | --- | --- | --- | --- | --- | --- | --- | --- | --- | --- | --- |
|  | **N** | **n** | **OR** | **95% CI** | **p-value** | | **N** | | **n** | **OR** | | **95% CI** | **p-value** | **N** | **n** | **OR** | **95% CI** | **p-value** |
| **Maternal** | | | | | | | | | | | | | | | | | | |
| **ADHD (RS-DBD)** | 42206 | 5607 |  |  | 0.002 | | 34297 | | 4415 |  | |  | 0.019 | 12621 | 1686 |  |  | 0.045 |
| 0-49mg (ref) | 26564 | 3506 | - | - |  | | 21616 | | 2757 | - | | - |  | 7844 | 1021 | - | - |  |
| 50-199mg | 12867 | 1651 | 0.968 | 0.909,1.031 |  | | 10462 | | 1303 | 0.993 | | 0.922,1.068 |  | 4133 | 564 | 1.073 | 0.957,1.203 |  |
| 200-299mg | 1869 | 282 | 1.169 | 1.024,1.334 |  | | 1497 | | 215 | 1.135 | | 0.969,1.329 |  | 473 | 66 | 1.085 | 0.820,1.435 |  |
| >300 mg | 906 | 168 | 1.497 | 1.258,1.782 |  | | 722 | | 140 | 1.403 | | 1.142,1.723 |  | 171 | 35 | 1.607 | 1.073,2.409 |  |
| **Hyperactive** | 42198 | 5538 |  |  | <0.001 | | 34290 | | 4353 |  | |  | <0.001 | 12620 | 1630 |  |  | <0.001 |
| 0-49mg (ref) | 26564 | 3386 | - | - |  | | 21615 | | 2644 | - | | - |  | 7845 | 956 | - | - |  |
| 50-199mg | 12861 | 1701 | 1.043 | 0.980,1.111 |  | | 10456 | | 1348 | 1.080 | | 1.004,1.162 |  | 4130 | 573 | 1.187 | 1.058,1.332 |  |
| 200-299mg | 1868 | 282 | 1.217 | 1.067,1.389 |  | | 1497 | | 221 | 1.206 | | 1.031,1.411 |  | 474 | 65 | 1.166 | 0.878,1.550 |  |
| >300 mg | 905 | 169 | 1.572 | 1.322,1.869 |  | | 722 | | 140 | 1.436 | | 1.171,1.760 |  | 171 | 36 | 1.857 | 1.250,2.758 |  |
| **Inattentive** | 42215 | 4913 |  |  | 0.070 | | 34302 | | 3874 |  | |  | 0.115 | 12620 | 1448 |  |  | 0.249 |
| 0-49mg (ref) | 26569 | 3088 | - | - |  | | 21619 | | 2435 | - | | - |  | 7845 | 888 | - | - |  |
| 50-199mg | 12872 | 1443 | 0.960 | 0.898,1.026 |  | | 10465 | | 1135 | 0.984 | | 0.911,1.064 |  | 4132 | 474 | 1.025 | 0.906,1.159 |  |
| 200-299mg | 1869 | 256 | 1.207 | 1.051,1.386 |  | | 1497 | | 200 | 1.221 | | 1.038,1.437 |  | 473 | 58 | 1.085 | 0.807,1.459 |  |
| >300 mg | 905 | 126 | 1.230 | 1.014,1.492 |  | | 721 | | 104 | 1.151 | | 0.917,1.443 |  | 170 | 28 | 1.435 | 0.936,2.198 |  |
| **Paternal** | | | | | | | | | | | | | | | | | | |
| **ADHD (RS-DBD)** | 15348 | 2085 |  |  | 0.365 | 10804 | | 1462 | | |  |  | 0.535 | 10804 | 1462 |  |  | 0.710 |
| 0-49mg (ref) | 3433 | 511 | - | - |  | 2353 | | 344 | | | - | - |  | 2353 | 344 | - | - |  |
| 50-199mg | 5527 | 705 | 0.836 | 0.739,0.945 |  | 3861 | | 478 | | | 0.854 | 0.731,0.999 |  | 3861 | 478 | 0.842 | 0.720,0.985 |  |
| 200-299mg | 4518 | 609 | 0.891 | 0.784,1.012 |  | 3231 | | 448 | | | 0.985 | 0.838,1.157 |  | 3231 | 448 | 0.965 | 0.820,1.136 |  |
| >300 mg | 1870 | 260 | 0.923 | 0.786,1.085 |  | 1359 | | 192 | | | 1.006 | 0.824,1.227 |  | 1359 | 192 | 0.982 | 0.804,1.199 |  |
| **Hyperactive** | 15349 | 2045 |  |  | 0.189 | 10802 | | 1432 | | |  |  | 0.929 | 10802 | 1432 |  |  | 0.729 |
| 0-49mg (ref) | 3433 | 491 | - | - |  | 2352 | | 332 | | | - | - |  | 2352 | 332 | - | - |  |
| 50-199mg | 5530 | 723 | 0.901 | 0.796,1.020 |  | 3862 | | 489 | | | 0.910 | 0.778,1.065 |  | 3862 | 489 | 0.892 | 0.762,1.045 |  |
| 200-299mg | 4517 | 582 | 0.886 | 0.778,1.009 |  | 3230 | | 428 | | | 0.960 | 0.815,1.132 |  | 3230 | 428 | 0.936 | 0.794,1.104 |  |
| >300 mg | 1869 | 249 | 0.921 | 0.781,1.086 |  | 1358 | | 183 | | | 0.977 | 0.798,1.197 |  | 1358 | 183 | 0.949 | 0.775,1.163 |  |
| **Inattentive** | 15350 | 1785 |  |  | 0.640 | 10806 | | 1254 | | |  |  | 0.492 | 10806 | 1254 |  |  | 0.569 |
| 0-49mg (ref) | 3431 | 437 | - | - |  | 2351 | | 288 | | | - | - |  | 2351 | 288 | - | - |  |
| 50-199mg | 5532 | 599 | 0.832 | 0.729,0.949 |  | 3866 | | 427 | | | 0.927 | 0.785,1.096 |  | 3866 | 427 | 0.919 | 0.777,1.086 |  |
| 200-299mg | 4517 | 518 | 0.887 | 0.774,1.017 |  | 3230 | | 368 | | | 0.972 | 0.816,1.158 |  | 3230 | 368 | 0.959 | 0.804,1.143 |  |
| >300 mg | 1870 | 231 | 0.966 | 0.814,1.145 |  | 1359 | | 171 | | | 1.085 | 0.877,1.342 |  | 1359 | 171 | 1.067 | 0.862,1.322 |  |

Note: RS-DBD – the Disruptive Behaviour Disorders scale; N – sample size; n – number of cases; OR – odds ratio; 95% CI – 95% confidence intervals; *adjusted for child’s gender, birth year, parity, parental age, education, marital status, financial difficulties, depression and anxiety symptoms, prenatal smoking and alcohol consumption; **additionally adjusted for partner’s caffeine consumption

### Supplementary Table S47. Associations of maternal and paternal daily prenatal caffeine consumption on high risk of maternal reported offspring ADHD symptoms in ALSPAC

|  |  | |  | | **Unadjusted model** | | | |  | |  | **Adjusted model*** | | |  |  | **Mutually adjusted model**** | | |
| --- | --- | --- | --- | --- | --- | --- | --- | --- | --- | --- | --- | --- | --- | --- | --- | --- | --- | --- | --- |
|  | **N** | | **n** | | **OR** | | **95% CI** | **p-value** | **N** | | **n** | **OR** | **95% CI** | **p-value** | **N** | **n** | **OR** | **95% CI** | **p-value** |
| **Maternal** | | | | | | | | | | | | | | | | | | | |
| **ADHD (DAWBA)** | 7680 | | 1105 | |  | |  | <0.001 | 6675 | | 934 |  |  | 0.045 | 5447 | 745 |  |  | 0.191 |
| 0-49mg (ref) | 1026 | | 119 | | - | | - |  | 895 | | 104 | - | - |  | 735 | 84 | - | - |  |
| 50-199mg | 3149 | | 435 | | 1.222 | | 0.984,1.517 |  | 2765 | | 369 | 1.128 | 0.888,1.432 |  | 2284 | 304 | 1.096 | 0.840,1.429 |  |
| 200-299mg | 1871 | | 270 | | 1.285 | | 1.021,1.619 |  | 1638 | | 227 | 1.151 | 0.891,1.486 |  | 1354 | 175 | 1.023 | 0.767,1.365 |  |
| >300mg/day | 1634 | | 281 | | 1.583 | | 1.257,1.993 |  | 1377 | | 234 | 1.310 | 1.008,1.701 |  | 1074 | 182 | 1.260 | 0.937,1.695 |  |
| **Hyperactive** | 7701 | | 978 | |  | |  | 0.006 | 6693 | | 828 |  |  | 0.269 | 5458 | 653 |  |  | 0.381 |
| 0-49mg (ref) | 1027 | | 105 | | - | | - |  | 895 | | 93 | - | - |  | 735 | 74 | - | - |  |
| 50-199mg | 3154 | | 400 | | 1.275 | | 1.016,1.601 |  | 2769 | | 339 | 1.166 | 0.909,1.495 |  | 2284 | 273 | 1.127 | 0.853,1.489 |  |
| 200-299mg | 1878 | | 238 | | 1.274 | | 0.999,1.625 |  | 1644 | | 199 | 1.132 | 0.866,1.480 |  | 1359 | 152 | 1.022 | 0.755,1.384 |  |
| >300mg | 1642 | | 235 | | 1.467 | | 1.148,1.873 |  | 1385 | | 197 | 1.224 | 0.930,1.611 |  | 1080 | 154 | 1.224 | 0.896,1.673 |  |
| **Inattentive** | 7692 | | 1047 | |  | |  | 0.268 | 6688 | | 890 |  |  | 0.948 | 5455 | 722 |  |  | 0.953 |
| 0-49mg(ref) | 1025 | | 132 | | - | | - |  | 895 | | 113 | - | - |  | 735 | 90 | - | - |  |
| 50-199mg | 3161 | | 424 | | 1.048 | | 0.850,1.292 |  | 2775 | | 363 | 1.004 | 0.796,1.266 |  | 2292 | 302 | 0.997 | 0.769,1.293 |  |
| 200-299mg | 1875 | | 258 | | 1.079 | | 0.862,1.352 |  | 1643 | | 219 | 1.009 | 0.786,1.297 |  | 1356 | 171 | 0.931 | 0.701,1.236 |  |
| >300mg | 1631 | | 233 | | 1.128 | | 0.896,1.418 |  | 1375 | | 195 | 0.991 | 0.763,1.288 |  | 1072 | 159 | 1.034 | 0.769,1.391 |  |
| **ADHD (SDQ)** | 7943 | | 879 | |  | |  | 0.009 | 6946 | | 737 |  |  | 0.710 | 5662 | 586 |  |  | 0.558 |
| 0-49mg(ref) | 1047 | | 113 | | - | | - |  | 926 | | 96 | - | - |  | 766 | 77 | - | - |  |
| 50-199mg | 3228 | | 333 | | 0.951 | | 0.759,1.192 |  | 2847 | | 288 | 0.945 | 0.737,1.212 |  | 2349 | 229 | 0.913 | 0.691,1.206 |  |
| 200-299mg | 1965 | | 204 | | 0.958 | | 0.751,1.221 |  | 1722 | | 166 | 0.863 | 0.658,1.132 |  | 1411 | 127 | 0.810 | 0.596,1.100 |  |
| >300mg | 1703 | | 229 | | 1.284 | | 1.010,1.632 |  | 1451 | | 187 | 1.057 | 0.804,1.390 |  | 1136 | 153 | 1.091 | 0.801,1.486 |  |
| **Paternal** | | | | | | | | | | | | | | | | | | | |
| **ADHD (DAWBA)** | | 6124 | | 859 | |  |  | 0.325 | 4657 | 622 | |  |  | 0.811 | 4625 | 617 |  |  | 0.786 |
| 0-49mg (ref) | | 220 | | 27 | |  |  |  | 152 | 18 | |  |  |  | 151 | 18 |  |  |  |
| 50-199mg | | 773 | | 102 | | 1.087 | 0.691,1.710 |  | 596 | 77 | | 1.147 | 0.655,2.008 |  | 593 | 77 | 1.110 | 0.634,1.945 |  |
| 200-299mg | | 950 | | 135 | | 1.184 | 0.761,1.842 |  | 733 | 95 | | 1.094 | 0.631,1.895 |  | 729 | 94 | 1.021 | 0.588,1.773 |  |
| >300mg | | 4181 | | 595 | | 1.186 | 0.786,1.791 |  | 3176 | 432 | | 1.129 | 0.674,1.890 |  | 3152 | 428 | 1.033 | 0.615,1.734 |  |
| **Hyperactive** | | 6135 | | 749 | |  |  | 0.411 | 4666 | 535 | |  |  | 0.719 | 4634 | 533 |  |  | 0.976 |
| 0-49mg (ref) | | 219 | | 24 | |  |  |  | 151 | 18 | |  |  |  | 150 | 18 |  |  |  |
| 50-199mg | | 771 | | 88 | | 1.047 | 0.649,1.689 |  | 596 | 66 | | 0.969 | 0.550,1.708 |  | 593 | 66 | 0.943 | 0.535,1.663 |  |
| 200-299mg | | 953 | | 119 | | 1.159 | 0.728,1.846 |  | 736 | 76 | | 0.857 | 0.490,1.498 |  | 732 | 76 | 0.821 | 0.469,1.438 |  |
| >300mg | | 4192 | | 518 | | 1.146 | 0.742,1.768 |  | 3183 | 375 | | 0.991 | 0.592,1.662 |  | 3159 | 373 | 0.932 | 0.554,1.567 |  |
| **Inattentive** | | 6133 | | 822 | |  |  | 0.784 | 4666 | 614 | |  |  | 0.518 | 4633 | 609 |  |  | 0.726 |
| 0-49mg | | 220 | | 25 | |  |  |  | 152 | 14 | |  |  |  | 151 | 14 |  |  |  |
| 50-199mg | | 777 | | 108 | | 1.259 | 0.792,2.001 |  | 600 | 78 | | 1.554 | 0.844,2.860 |  | 597 | 78 | 1.523 | 0.827,2.805 |  |
| 200-299mg | | 951 | | 127 | | 1.202 | 0.762,1.897 |  | 734 | 99 | | 1.549 | 0.850,2.823 |  | 730 | 97 | 1.461 | 0.800,2.667 |  |
| >300mg | | 4185 | | 562 | | 1.210 | 0.790,1.852 |  | 3180 | 423 | | 1.520 | 0.859,2.689 |  | 3155 | 420 | 1.437 | 0.810,2.550 |  |
| **ADHD (SDQ)** | | 6323 | | 678 | |  |  | 0.346 | 4804 | 487 | |  |  | 0.346 | 4772 | 481 |  |  | 0.633 |
| 0-49mg | | 216 | | 22 | |  |  |  | 148 | 12 | |  |  |  | 147 | 12 |  |  |  |
| 50-199mg | | 790 | | 76 | | 0.939 | 0.569,1.548 |  | 615 | 58 | | 1.288 | 0.667,2.490 |  | 612 | 58 | 1.258 | 0.650,2.432 |  |
| 200-299mg | | 994 | | 108 | | 1.075 | 0.662,1.744 |  | 757 | 70 | | 1.184 | 0.619,2.266 |  | 753 | 68 | 1.089 | 0.568,2.090 |  |
| >300mg | | 4323 | | 472 | | 1.081 | 0.688,1.697 |  | 3284 | 347 | | 1.342 | 0.729,2.471 |  | 3260 | 343 | 1.229 | 0.665,2.271 |  |

Note: DAWBA – Development and Well-Being Assessment; SDQ – Strengths and Difficulties Questionnaire; N – total sample size; n – number of ADHD cases; OR – odds ratio; 95% CI – 95% confidence intervals; *adjusted for child’s gender, ethnicity, parity, parental age, marital status, education, financial difficulties, depression and anxiety symptoms, prenatal smoking and alcohol use; **additionally adjusted for partner’s prenatal caffeine use

### Supplementary Table S48. Associations of maternal daily prenatal caffeine consumption on high risk of maternal and teacher reported offspring ADHD symptoms in GenR

|  |  |  | **Unadjusted model** | | |  |  | **Adjusted model*** | | |
| --- | --- | --- | --- | --- | --- | --- | --- | --- | --- | --- |
|  | **N** | **n** | **OR** | **95% CI** | **p-value** | **N** | **n** | **OR** | **95% CI** | **p-value** |
| **ADHD (CPRS-R)** | 2613 | 371 |  |  | 0.507 | 2053 | 282 |  |  | 0.438 |
| 0-49mg (ref) | 499 | 83 | - | - |  | 383 | 62 | - | - |  |
| 50-199mg | 1251 | 169 | 0.783 | 0.588,1.042 |  | 972 | 125 | 0.735 | 0.520,1.039 |  |
| 200-299mg | 443 | 55 | 0.710 | 0.492,1.026 |  | 357 | 43 | 0.711 | 0.456,1.106 |  |
| >300mg | 420 | 64 | 0.901 | 0.632,1.286 |  | 341 | 52 | 0.832 | 0.534,1.297 |  |
| **Hyperactive** | 2617 | 273 |  |  | 0.236 | 2057 | 201 |  |  | 0.387 |
| 0-49mg (ref) | 499 | 65 | - | - |  | 383 | 43 | - | - |  |
| 50-199mg | 1255 | 124 | 0.732 | 0.532,1.008 |  | 976 | 94 | 0.851 | 0.572,1.266 |  |
| 200-299mg | 444 | 39 | 0.643 | 0.423,0.978 |  | 358 | 29 | 0.735 | 0.438,1.235 |  |
| >300mg | 419 | 45 | 0.803 | 0.536,1.204 |  | 340 | 35 | 0.841 | 0.500,1.414 |  |
| **Inattentive** | 2616 | 312 |  |  | 0.770 | 2058 | 240 |  |  | 0.701 |
| 0-49mg (ref) | 499 | 70 | - | - |  | 383 | 53 | - | - |  |
| 50-199mg | 1253 | 139 | 0.765 | 0.562,1.041 |  | 976 | 102 | 0.685 | 0.473,0.992 |  |
| 200-299mg | 444 | 47 | 0.726 | 0.489,1.076 |  | 358 | 40 | 0.766 | 0.483,1.214 |  |
| >300mg | 420 | 56 | 0.943 | 0.646,1.376 |  | 341 | 45 | 0.840 | 0.526,1.342 |  |
| **ADHD (CBCL)** | 3462 | 489 |  |  | 0.230 | 2565 | 331 |  |  | 0.285 |
| 0-49mg (ref) | 688 | 109 | - | - |  | 490 | 64 | - | - |  |
| 50-199mg | 1648 | 232 | 0.870 | 0.680,1.114 |  | 1216 | 155 | 1.093 | 0.788,1.516 |  |
| 200-299mg | 591 | 73 | 0.749 | 0.544,1.030 |  | 445 | 54 | 1.120 | 0.742,1.690 |  |
| >300mg | 535 | 75 | 0.866 | 0.630,1.191 |  | 414 | 58 | 1.259 | 0.824,1.923 |  |
| **ADHD (TRF)** | 2543 | 402 |  |  | 0.007 | 1671 | 218 |  |  | 0.128 |
| 0-49mg (ref) | 565 | 95 | - | - |  | 340 | 48 | - | - |  |
| 50-199mg | 1197 | 208 | 1.040 | 0.797,1.358 |  | 777 | 112 | 1.171 | 0.791,1.734 |  |
| 200-299mg | 426 | 60 | 0.811 | 0.571,1.152 |  | 296 | 31 | 0.869 | 0.513,1.472 |  |
| >300mg | 355 | 39 | 0.611 | 0.410,0.910 |  | 258 | 27 | 0.678 | 0.383,1.200 |  |

Note: CPRS-R – The revised Conners’ Parent Rating Scale; CBCL – Child Behaviour Checklist; TRF – Teacher’s Report Form; N – sample size; n –

number of cases; OR – odds ratio; 95% CI – 95% confidence intervals; *adjusted for child’s gender, parity, maternal ethnicity, age, education, anxiety and depression problems, financial difficulties and prenatal smoking and alcohol use

### Supplementary Table S49. Associations of maternal daily prenatal caffeine consumption on high risk of maternal reported offspring ADHD symptoms in MoBa (additionally adjusted for maternal ADHD)

|  |  |  | **Unadjusted model** | | |  |  | **Adjusted model*** | | |  |  | **Mutually adjusted model**** | | |
| --- | --- | --- | --- | --- | --- | --- | --- | --- | --- | --- | --- | --- | --- | --- | --- |
|  | **N** | **n** | **OR** | **95% CI** | **p-value** | **N** | **n** | **OR** | **95% CI** | **p-value** | **N** | **n** | **OR** | **95% CI** | **p-value** |
| **ADHD (RS-DBD)** | 42206 | 5607 |  |  | 0.002 | 28,507 | 3655 |  |  | 0.025 | 10,649 | 1441 |  |  | 0.099 |
| 0-49mg (ref) | 26564 | 3506 | - | - |  | 18,128 | 2294 | - | - |  | 6695 | 880 | - | - |  |
| 50-199mg | 12867 | 1651 | 0.968 | 0.909,1.031 |  | 8590 | 1078 | 1.003 | 0.925,1.087 |  | 3423 | 477 | 1.069 | 0.943,1.211 |  |
| 200-299mg | 1869 | 282 | 1.169 | 1.024,1.334 |  | 1206 | 175 | 1.149 | 0.964,1.368 |  | 387 | 55 | 1.045 | 0.767,1.424 |  |
| >300 mg | 906 | 168 | 1.497 | 1.258,1.782 |  | 583 | 108 | 1.378 | 1.092,1.738 |  | 144 | 29 | 1.558 | 0.997,2.434 |  |
| **Hyperactive** | 42198 | 5538 |  |  | <0.001 | 28,504 | 3600 |  |  | <0.001 | 10,649 | 1390 |  |  | 0.001 |
| 0-49mg (ref) | 26564 | 3386 | - | - |  | 18,129 | 2208 | - | - |  | 6697 | 825 | - | - |  |
| 50-199mg | 12861 | 1701 | 1.043 | 0.980,1.111 |  | 8586 | 1102 | 1.076 | 0.993,1.166 |  | 3421 | 480 | 1.177 | 1.038,1.334 |  |
| 200-299mg | 1868 | 282 | 1.217 | 1.067,1.389 |  | 1206 | 192 | 1.245 | 1.048,1.480 |  | 387 | 57 | 1.225 | 0.903,1.664 |  |
| >300 mg | 905 | 169 | 1.572 | 1.322,1.869 |  | 583 | 108 | 1.420 | 1.127,1.788 |  | 144 | 28 | 1.705 | 1.093,2.660 |  |
| **Inattentive** | 42215 | 4913 |  |  | 0.070 | 28,512 | 3186 |  |  | 0.271 | 10,649 | 1230 |  |  | 0.473 |
| 0-49mg (ref) | 26569 | 3088 | - | - |  | 18,130 | 2031 | - | - |  | 6696 | 766 | - | - |  |
| 50-199mg | 12872 | 1443 | 0.960 | 0.898,1.026 |  | 8594 | 916 | 0.966 | 0.886,1.052 |  | 3423 | 394 | 1.009 | 0.882,1.154 |  |
| 200-299mg | 1869 | 256 | 1.207 | 1.051,1.386 |  | 1206 | 158 | 1.193 | 0.995,1.431 |  | 387 | 47 | 1.030 | 0.740,1.432 |  |
| >300 mg | 905 | 126 | 1.230 | 1.014,1.492 |  | 582 | 81 | 1.154 | 0.895,1.489 |  | 143 | 23 | 1.386 | 0.868,2.215 |  |

Note: RS-DBD – the Disruptive Behaviour Disorders scale; N – sample size; n – number of cases; OR – odds ratio; 95% CI – 95% confidence intervals; *adjusted for child’s gender, birth year, parity, parental age, education, marital status, financial difficulties, depression and anxiety symptoms, prenatal smoking and alcohol consumption; ** additionally adjusted for partner’s caffeine consumption

### Supplementary Table S50. Associations of maternal daily caffeine consumption before pregnancy on high risk of maternal reported offspring ADHD symptoms in MoBa

|  |  |  | **Unadjusted model** | | |  |  | **Adjusted model*** | | |  |  | **Mutually adjusted model**** | | |
| --- | --- | --- | --- | --- | --- | --- | --- | --- | --- | --- | --- | --- | --- | --- | --- |
|  | **N** | **n** | **OR** | **95% CI** | **p-value** | **N** | **n** | **OR** | **95% CI** | **p-value** | **N** | **n** | **OR** | **95% CI** | **p-value** |
| **ADHD (RS-DBD)** | 42,206 | 5607 |  |  | 0.057 | 36,636 | 4750 |  |  | 0.373 | 13,573 | 1819 |  |  | 0.276 |
| 0-49mg (ref) | 17,079 | 2297 | - | - |  | 14,480 | 1905 | - | - |  | 5105 | 672 | - | - |  |
| 50-199mg | 12,139 | 1500 | 0.907 | 0.846,0.974 |  | 10,711 | 1306 | 0.971 | 0.898,1.051 |  | 4251 | 562 | 1.057 | 0.933,1.197 |  |
| 200-299mg | 6351 | 836 | 0.976 | 0.896,1.062 |  | 5606 | 703 | 0.990 | 0.898,1.091 |  | 2198 | 300 | 1.106 | 0.948,1.290 |  |
| >300 mg | 6637 | 974 | 1.107 | 1.020,1.201 |  | 5839 | 836 | 1.058 | 0.962,1.162 |  | 2019 | 285 | 1.071 | 0.913,1.257 |  |
| **Hyperactive** | 42,198 | 5357 |  |  | 0.004 | 36,626 | 4693 |  |  | 0.050 | 13,572 | 1775 |  |  | 0.035 |
| 0-49mg (ref) | 17,079 | 2257 | - | - |  | 14,479 | 1857 | - | - |  | 5105 | 636 | - | - |  |
| 50-199mg | 12,136 | 1452 | 0.892 | 0.831,0.958 |  | 10,707 | 1264 | 0.963 | 0.890,1.042 |  | 4250 | 557 | 1.114 | 0.982,1.264 |  |
| 200-299mg | 6348 | 842 | 1.004 | 0.922,1.094 |  | 5603 | 718 | 1.035 | 0.940,1.140 |  | 2197 | 287 | 1.107 | 0.946,1.295 |  |
| >300 mg | 6635 | 986 | 1.146 | 1.056,1.244 |  | 5837 | 854 | 1.102 | 1.004,1.211 |  | 2020 | 295 | 1.182 | 1.008,1.385 |  |
| **Inattentive** | 42,215 | 4913 |  |  | 0.115 | 36,642 | 4169 |  |  | 0.179 | 13,573 | 1564 |  |  | 0.206 |
| 0-49mg (ref) | 17,078 | 2007 | - | - |  | 14,479 | 1661 | - | - |  | 5104 | 577 | - | - |  |
| 50-199mg | 12,148 | 1320 | 0.915 | 0.850,0.986 |  | 10,717 | 1159 | 1.004 | 0.924,1.091 |  | 4254 | 477 | 1.050 | 0.918,1.200 |  |
| 200-299mg | 6352 | 753 | 1.010 | 0.923,1.105 |  | 5607 | 626 | 1.039 | 0.938,1.151 |  | 2198 | 265 | 1.158 | 0.982,1.364 |  |
| >300 mg | 6637 | 833 | 1.078 | 0.988,1.176 |  | 5839 | 723 | 1.073 | 0.971,1.186 |  | 2017 | 245 | 1.084 | 0.915,1.286 |  |

Note: RS-DBD – the Disruptive Behaviour Disorders scale; N – sample size; n – number of cases; OR – odds ratio; 95% CI – 95% confidence intervals; *adjusted for child’s gender, birth year, parity, maternal age, education, marital status, financial difficulties, depression and anxiety symptoms, smoking and alcohol consumption before pregnancy; **additionally adjusted for partner’s caffeine consumption

### Supplementary Table S51. Associations of maternal and paternal daily prenatal caffeine consumption on high risk of teacher reported offspring ADHD symptoms in ALSPAC

|  |  |  | | **Unadjusted model** | | | | |  |  | | | **Adjusted model*** | | | | |  |  | **Mutually adjusted model**** | | | |
| --- | --- | --- | --- | --- | --- | --- | --- | --- | --- | --- | --- | --- | --- | --- | --- | --- | --- | --- | --- | --- | --- | --- | --- |
|  | **N** | **n** | | **OR** | **95% CI** | | **p-value** | | **N** | **n** | | | **OR** | | **95% CI** | | **p-value** | **N** | **n** | **OR** | | **95% CI** | **p-value** |
| **Maternal** | | | | | | | | | | | | | | | | | | | | | | | |
| **ADHD (DAWBA)** | 5726 | 806 | |  |  | | 0.008 | | 4584 | 576 | | |  | |  | | 0.596 | 3611 | 428 |  | |  | 0.247 |
| 0-49mg (ref) | 728 | 95 | | - | - | |  | | 576 | 66 | | | - | | - | |  | 463 | 47 | - | | - |  |
| 50-199mg | 2259 | 293 | | 0.993 | 0.775,1.273 | |  | | 1855 | 215 | | | 1.077 | | 0.793,1.462 | |  | 1483 | 155 | 1.075 | | 0.752,1.537 |  |
| 200-299mg | 1402 | 201 | | 1.115 | 0.858,1.450 | |  | | 1143 | 154 | | | 1.217 | | 0.881,1.680 | |  | 904 | 124 | 1.423 | | 0.979,2.068 |  |
| >300mg | 1337 | 217 | | 1.291 | 0.995,1.674 | |  | | 1010 | 141 | | | 1.069 | | 0.765,1.494 | |  | 761 | 102 | 1.136 | | 0.767,1.683 |  |
| **Hyperactive** | 5725 | 721 | |  |  | | 0.049 | | 4584 | 520 | | |  | |  | | 0.757 | 3611 | 384 |  | |  | 0.339 |
| 0-49mg (ref) | 728 | 87 | | - | - | |  | | 576 | 62 | | | - | | - | |  | 463 | 44 | - | | - |  |
| 50-199mg | 2259 | 265 | | 0.979 | 0.756,1.267 | |  | | 1855 | 194 | | | 1.001 | | 0.731,1.370 | |  | 1483 | 143 | 1.053 | | 0.728,1.523 |  |
| 200-299mg | 1401 | 181 | | 1.093 | 0.832,1.436 | |  | | 1143 | 140 | | | 1.159 | | 0.832,1.614 | |  | 904 | 107 | 1.301 | | 0.882,1.919 |  |
| >300mg | 1337 | 188 | | 1.206 | 0.919,1.582 | |  | | 1010 | 124 | | | 0.999 | | 0.707,1.410 | |  | 761 | 90 | 1.121 | | 0.746,1.686 |  |
| **Inattentive** | 5726 | 710 | |  |  | | 0.014 | | 4583 | 508 | | |  | |  | | 0.874 | 3609 | 374 |  | |  | 0.610 |
| 0-49mg (ref) | 727 | 79 | | - | - | |  | | 575 | 56 | | | - | | - | |  | 462 | 38 | - | | - |  |
| 50-199mg | 2259 | 263 | | 1.081 | 0.828,1.411 | |  | | 1854 | 200 | | | 1.206 | | 0.873,1.666 | |  | 1482 | 147 | 1.296 | | 0.883,1.904 |  |
| 200-299mg | 1402 | 181 | | 1.216 | 0.918,1.610 | |  | | 1143 | 133 | | | 1.239 | | 0.879,1.746 | |  | 904 | 104 | 1.438 | | 0.959,2.156 |  |
| >300mg | 1338 | 187 | | 1.333 | 1.007,1.763 | |  | | 1011 | 119 | | | 1.092 | | 0.765,1.560 | |  | 761 | 85 | 1.163 | | 0.761,1.779 |  |
| **ADHD (SDQ)** | 5723 | 633 | |  |  | | 0.014 | | 4587 | 444 | | |  | |  | | 0.830 | 3613 | 328 |  | |  | 0.309 |
| 0-49mg (ref) | 727 | 74 | | - | - | |  | | 577 | 52 | | | - | | - | |  | 464 | 35 | - | | - |  |
| 50-199mg | 2261 | 231 | | 1.004 | 0.762,1.324 | |  | | 1858 | 165 | | | 1.039 | | 0.740,1.459 | |  | 1485 | 117 | 1.058 | | 0.705,1.589 |  |
| 200-299mg | 1400 | 155 | | 1.099 | 0.820,1.472 | |  | | 1142 | 116 | | | 1.133 | | 0.790,1.623 | |  | 904 | 94 | 1.365 | | 0.894,2.084 |  |
| >300mg | 1335 | 173 | | 1.314 | 0.984,1.753 | |  | | 1010 | 111 | | | 1.023 | | 0.706,1.483 | |  | 760 | 82 | 1.136 | | 0.730,1.768 |  |
| **Paternal** | | | | | | | | | | | | | | | | | | | | | | | |
| **ADHD (DAWBA)** | 4268 | 551 |  | | |  | | 0.760 | 3075 | | 334 |  | |  | | 0.309 | | 3053 | 330 | |  |  | 0.240 |
| 0-49mg (ref) | 134 | 20 | - | | | - | |  | 98 | | 13 | - | | - | |  | | 97 | 13 | | - | - |  |
| 50-199mg | 511 | 64 | 0.816 | | | 0.474,1.404 | |  | 367 | | 39 | 0.867 | | 0.430,1.746 | |  | | 366 | 38 | | 0.812 | 0.403,1.638 |  |
| 200-299mg | 648 | 85 | 0.861 | | | 0.508,1.458 | |  | 465 | | 51 | 0.798 | | 0.403,1.578 | |  | | 461 | 51 | | 0.769 | 0.389,1.522 |  |
| >300mg | 2975 | 382 | 0.840 | | | 0.516,1.367 | |  | 2145 | | 231 | 0.760 | | 0.406,1.425 | |  | | 2129 | 228 | | 0.710 | 0.378,1.336 |  |
| **Hyperactive** | 4267 | 494 |  | | |  | | 0.349 | 3075 | | 313 |  | |  | | 0.071 | | 3053 | 306 | |  |  | 0.086 |
| 0-49mg (ref) | 134 | 21 | - | | | - | |  | 98 | | 15 | - | | - | |  | | 97 | 14 | | - | - |  |
| 50-199mg | 511 | 57 | 0.676 | | | 0.393,1.161 | |  | 367 | | 39 | 0.756 | | 0.386,1.482 | |  | | 366 | 38 | | 0.777 | 0.390,1.546 |  |
| 200-299mg | 648 | 79 | 0.747 | | | 0.443,1.259 | |  | 465 | | 47 | 0.632 | | 0.327,1.222 | |  | | 461 | 47 | | 0.673 | 0.343,1.320 |  |
| >300mg | 2974 | 337 | 0.688 | | | 0.426,1.111 | |  | 2145 | | 212 | 0.602 | | 0.331,1.096 | |  | | 2129 | 207 | | 0.621 | 0.335,1.153 |  |
| **Inattentive** | 4266 | 480 |  | | |  | | 0.289 | 3073 | | 307 |  | |  | | 0.937 | | 3051 | 303 | |  |  | 0.942 |
| 0-49mg (ref) | 134 | 17 | - | | | - | |  | 98 | | 12 | - | | - | |  | | 97 | 12 | | - | - |  |
| 50-199mg | 511 | 53 | 0.796 | | | 0.445,1.426 | |  | 367 | | 33 | 0.747 | | 0.363,1.541 | |  | | 366 | 33 | | 0.732 | 0.355,1.510 |  |
| 200-299mg | 647 | 58 | 0.678 | | | 0.381,1.205 | |  | 464 | | 37 | 0.580 | | 0.284,1.184 | |  | | 460 | 37 | | 0.571 | 0.280,1.168 |  |
| >300mg | 2974 | 352 | 0.924 | | | 0.549,1.555 | |  | 2144 | | 225 | 0.773 | | 0.407,1.469 | |  | | 2128 | 221 | | 0.746 | 0.391,1.422 |  |
| **ADHD (SDQ)** | 4265 | 426 |  | | |  | | 0.425 | 3079 | | 261 |  | |  | | 0.679 | | 3057 | 258 | |  |  | 0.483 |
| 0-49mg (ref) | 134 | 15 | - | | | - | |  | 98 | | 11 | - | | - | |  | | 97 | 11 | | - | - |  |
| 50-199mg | 511 | 44 | 0.747 | | | 0.402,1.389 | |  | 368 | | 26 | 0.653 | | 0.302,1.413 | |  | | 367 | 26 | | 0.629 | 0.291,1.361 |  |
| 200-299mg | 647 | 61 | 0.826 | | | 0.454,1.502 | |  | 466 | | 39 | 0.683 | | 0.326,1.429 | |  | | 462 | 39 | | 0.656 | 0.314,1.374 |  |
| >300mg | 2973 | 306 | 0.910 | | | 0.525,1.577 | |  | 2147 | | 185 | 0.693 | | 0.353,1.361 | |  | | 2131 | 182 | | 0.641 | 0.325,1.265 |  |

Note: DAWBA – Development and Well-Being Assessment; SDQ – Strengths and Difficulties Questionnaire; N – total sample size; n – number of ADHD cases; OR – odds ratio; 95% CI – 95% confidence intervals; *adjusted for child’s gender, ethnicity, parity, parental age, marital status, education, financial difficulties, depression and anxiety symptoms, prenatal smoking and alcohol use; **additionally adjusted for partner’s prenatal caffeine use

### Supplementary Table S52. Associations of maternal and paternal daily prenatal caffeine consumption on high risk of maternal reported offspring ADHD symptoms in ALSPAC (complete cases)

|  |  |  | **Unadjusted model** | | | **Adjusted model*** | | | | | **Mutually adjusted model**** | | | | | |
| --- | --- | --- | --- | --- | --- | --- | --- | --- | --- | --- | --- | --- | --- | --- | --- | --- |
|  | **N** | **n** | **OR** | **95% CI** | **p-value** | **OR** | **95% CI** | | | **p-value** | **OR** | | | **95% CI** | **p-value** | |
| **Maternal** | | | | | | | | | | | | | | | | |
| **ADHD (DAWBA)** | 5447 | 745 |  |  | 0.002 |  |  | | | 0.131 |  | | |  | 0.191 | |
| 0-49mg (ref) | 735 | 84 | - | - |  | - | - | | |  | - | | | - |  | |
| 50-199mg | 2284 | 304 | 1.190 | 0.920,1.539 |  | 1.109 | 0.851,1.446 | | |  | 1.096 | | | 0.840,1.429 |  | |
| 200-299mg | 1354 | 175 | 1.150 | 0.872,1.518 |  | 1.044 | 0.784,1.390 | | |  | 1.023 | | | 0.767,1.365 |  | |
| >300mg | 1074 | 182 | 1.581 | 1.198,2.087 |  | 1.293 | 0.965,1.732 | | |  | 1.260 | | | 0.937,1.695 |  | |
| **Hyperactive** | 5458 | 653 |  |  | 0.021 |  |  | | | 0.299 |  | | |  | 0.381 | |
| 0-49mg (ref) | 735 | 74 | - | - |  | - | - | | |  | - | | | - |  | |
| 50-199mg | 2284 | 273 | 1.213 | 0.924,1.591 |  | 1.139 | 0.863,1.504 | | |  | 1.127 | | | 0.853,1.489 |  | |
| 200-299mg | 1359 | 152 | 1.125 | 0.839,1.509 |  | 1.040 | 0.769,1.405 | | |  | 1.022 | | | 0.755,1.384 |  | |
| >300mg | 1080 | 154 | 1.486 | 1.106,1.995 |  | 1.251 | 0.919,1.702 | | |  | 1.224 | | | 0.896,1.673 |  | |
| **Inattentive** | 5455 | 722 |  |  | 0.167 |  |  | | | 0.850 |  | | |  | 0.953 | |
| 0-49mg (ref) | 735 | 90 | - | - |  | - | - | | |  | -- | | | - |  | |
| 50-199mg | 2292 | 302 | 1.088 | 0.846,1.399 |  | 1.012 | 0.781,1.310 | | |  | 0.997 | | | 0.769,1.293 |  | |
| 200-299mg | 1356 | 171 | 1.034 | 0.787,1.358 |  | 0.948 | 0.716,1.256 | | |  | 0.931 | | | 0.701,1.236 |  | |
| >300mg | 1072 | 159 | 1.248 | 0.946,1.647 |  | 1.056 | 0.789,1.415 | | |  | 1.034 | | | 0.769,1.391 |  | |
| **ADHD (SDQ)** | 5662 | 586 |  |  | 0.014 |  |  | | | 0.499 |  | | |  | 0.558 | |
| 0-49mg (ref) | 766 | 77 | - | - |  | - | - | | |  | - | | | - |  | |
| 50-199mg | 2349 | 229 | 0.967 | 0.736,1.269 |  | 0.914 | 0.693,1.206 | | |  | 0.913 | | | 0.691,1.206 |  | |
| 200-299mg | 1411 | 127 | 0.885 | 0.657,1.192 |  | 0.814 | 0.601,1.104 | | |  | 0.810 | | | 0.596,1.100 |  | |
| >300mg | 1136 | 153 | 1.393 | 1.041,1.862 |  | 1.104 | 0.814,1.497 | | |  | 1.091 | | | 0.801,1.486 |  | |
| **Paternal** | | | | | | | | | | | | | | | | |
| **ADHD (DAWBA)** | 4625 | 617 |  |  | 0.473 |  | |  | 0.874 | | |  |  | | | 0.786 |
| 0-49mg (ref) | 151 | 18 | - | - |  | - | | - |  | | | - | - | | |  |
| 50-199mg | 593 | 77 | 1.103 | 0.638,1.906 |  | 1.135 | | 0.648,1.986 |  | | | 1.110 | 0.634,1.945 | | |  |
| 200-299mg | 729 | 94 | 1.094 | 0.639,1.873 |  | 1.069 | | 0.616,1.853 |  | | | 1.021 | 0.588,1.773 | | |  |
| >300mg | 3152 | 428 | 1.161 | 0.702,1.919 |  | 1.107 | | 0.661,1.853 |  | | | 1.033 | 0.615,1.734 | | |  |
| **Hyperactive** | 4634 | 533 |  |  | 0.568 |  | |  | 0.750 | | |  |  | | | 0.976 |
| 0-49mg (ref) | 150 | 18 | - | - |  | - | | - |  | | | - | - | | |  |
| 50-199mg | 593 | 66 | 0.918 | 0.527,1.600 |  | 0.958 | | 0.543,1.688 |  | | | 0.943 | 0.535,1.663 | | |  |
| 200-299mg | 732 | 76 | 0.850 | 0.492,1.468 |  | 0.848 | | 0.485,1.482 |  | | | 0.821 | 0.469,1.438 | | |  |
| >300mg | 3159 | 373 | 0.982 | 0.593,1.626 |  | 0.978 | | 0.583,1.640 |  | | | 0.932 | 0.554,1.567 | | |  |
| **Inattentive** | 4633 | 609 |  |  | 0.366 |  | |  | 0.530 | | |  |  | | | 0.726 |
| 0-49mg (ref) | 151 | 14 | - | - |  | - | | - |  | | | - | - | | |  |
| 50-199mg | 597 | 78 | 1.471 | 0.808,2.678 |  | 1.545 | | 0.839,2.845 |  | | | 1.523 | 0.827,2.805 | | |  |
| 200-299mg | 730 | 97 | 1.500 | 0.831,2.705 |  | 1.503 | | 0.824,2.742 |  | | | 1.461 | 0.800,2.667 | | |  |
| >300mg | 3155 | 420 | 1.503 | 0.859,2.629 |  | 1.502 | | 0.849,2.658 |  | | | 1.437 | 0.810,2.550 | | |  |
| **ADHD (SDQ)** | 4772 | 481 |  |  | 0.170 |  | |  | 0.376 | | |  |  | | | 0.633 |
| 0-49mg (ref) | 147 | 12 | - | - |  | - | | - |  | | | - | - | | |  |
| 50-199mg | 612 | 58 | 1.178 | 0.615,2.255 |  | 1.284 | | 0.664,2.481 |  | | | 1.258 | 0.650,2.432 | | |  |
| 200-299mg | 753 | 68 | 1.117 | 0.588,2.120 |  | 1.137 | | 0.594,2.179 |  | | | 1.089 | 0.568,2.090 | | |  |
| >300mg | 3260 | 343 | 1.323 | 0.725,2.413 |  | 1.318 | | 0.716,2.429 |  | | | 1.229 | 0.665,2.271 | | |  |

Note: DAWBA – Development and Well-Being Assessment; SDQ – Strengths and Difficulties Questionnaire; N – total sample size; n – number of ADHD cases; OR – odds ratio; 95% CI – 95% confidence intervals; *adjusted for child’s gender, ethnicity, parity, parental age, marital status, education, financial difficulties, depression and anxiety symptoms, prenatal smoking and alcohol use; **additionally adjusted for partner’s prenatal caffeine use

### Supplementary Table S53. Associations of maternal daily prenatal caffeine consumption on high risk of maternal and teacher

### reported offspring ADHD symptoms in GenR (complete cases)

|  |  |  | **Unadjusted model** | | | **Adjusted model*** | | |
| --- | --- | --- | --- | --- | --- | --- | --- | --- |
|  | **N** | **n** | **OR** | **95% CI** | **p-value** | **OR** | **95% CI** | **p-value** |
| **ADHD (CPRS-R)** | 2053 | 282 |  |  | 0.733 |  |  | 0.438 |
| 0-49mg (ref) | 383 | 62 | - | - |  | - | - |  |
| 50-199mg | 972 | 125 | 0.764 | 0.549,1.063 |  | 0.735 | 0.520,1.039 |  |
| 200-299mg | 357 | 43 | 0.709 | 0.466,1.078 |  | 0.711 | 0.456,1.106 |  |
| >300mg | 341 | 52 | 0.932 | 0.624,1.392 |  | 0.832 | 0.534,1.297 |  |
| **Hyperactive** | 2057 | 201 |  |  | 0536 |  |  | 0.387 |
| 0-49mg (ref) | 383 | 43 | - | - |  | - | - |  |
| 50-199mg | 976 | 94 | 0.843 | 0.575,1.235 |  | 0.851 | 0.572,1.266 |  |
| 200-299mg | 358 | 29 | 0.697 | 0.425,1.143 |  | 0.735 | 0.438,1.235 |  |
| >300mg | 340 | 35 | 0.907 | 0.566,1.455 |  | 0.841 | 0.500,1.414 |  |
| **Inattentive** | 2058 | 240 |  |  | 0.978 |  |  | 0.701 |
| 0-49mg (ref) | 383 | 53 | - | - |  | - | - |  |
| 50-199mg | 976 | 102 | 0.727 | 0.509,1.037 |  | 0.685 | 0.473,0.992 |  |
| 200-299mg | 358 | 40 | 0.783 | 0.505,1.214 |  | 0.766 | 0.483,1.214 |  |
| >300mg | 341 | 45 | 0.947 | 0.618,1.451 |  | 0.840 | 0.526,1.342 |  |
| **ADHD (CBCL)** | 2565 | 331 |  |  | 0.760 |  |  | 0.285 |
| 0-49mg (ref) | 490 | 64 | - | - |  | - | - |  |
| 50-199mg | 1216 | 155 | 0.972 | 0.712,1.329 |  | 1.093 | 0.788,1.516 |  |
| 200-299mg | 445 | 54 | 0.919 | 0.624,1.354 |  | 1.120 | 0.742,1.690 |  |
| >300mg | 414 | 58 | 1.084 | 0.740,1.589 |  | 1.259 | 0.824,1.923 |  |
| **ADHD (TRF)** | 1671 | 218 |  |  | 0.063 |  |  | 0.128 |
| 0-49mg (ref) | 340 | 48 | - | - |  | - | - |  |
| 50-199mg | 777 | 112 | 1.025 | 0.711,1.476 |  | 1.171 | 0.791,1.734 |  |
| 200-299mg | 296 | 31 | 0.712 | 0.440,1.151 |  | 0.869 | 0.513,1.472 |  |
| >300mg | 258 | 27 | 0.711 | 0.430,1.175 |  | 0.678 | 0.383,1.200 |  |

Note: CPRS-R – The revised Conners’ Parent Rating Scale; CBCL – Child Behaviour Checklist; TRF – Teacher’s Report Form;

N – sample size; n – number of cases; OR – odds ratio; 95% CI – 95% confidence intervals; *adjusted for child’s gender, parity,

maternal ethnicity, age, education, anxiety and depression problems, financial difficulties and prenatal smoking and alcohol use

### Supplementary Table S54. Associations of maternal and paternal daily prenatal caffeine consumption on high risk of maternal reported offspring ADHD symptoms in MoBa (complete cases)

|  |  | |  | | **Unadjusted model** | | | | | | **Adjusted model*** | | | | | **Mutually adjusted model**** | | | | | |
| --- | --- | --- | --- | --- | --- | --- | --- | --- | --- | --- | --- | --- | --- | --- | --- | --- | --- | --- | --- | --- | --- |
|  | **N** | | **n** | | **OR** | | **95% CI** | | **p-value** | | **OR** | | **95% CI** | | **p-value** | **OR** | | **95% CI** | | **p-value** | |
| **Maternal** | | | | | | | | | | | | | | | | | | | | | |
| **ADHD (RS-DBD)** | 12621 | | 1686 | |  | |  | | 0.030 | |  | |  | | 0.053 |  | |  | | 0.045 | |
| 0-49mg (ref) | 7844 | | 1021 | | - | | - | |  | | - | | - | |  | - | | - | |  | |
| 50-199mg | 4133 | | 564 | | 1.056 | | 0.946,1.179 | |  | | 1.064 | | 0.950,1.192 | |  | 1.073 | | 0.957,1.203 | |  | |
| 200-299mg | 473 | | 66 | | 1.084 | | 0.829,1.417 | |  | | 1.078 | | 0.815,1.425 | |  | 1.085 | | 0.820,1.435 | |  | |
| >300 mg | 171 | | 35 | | 1.720 | | 1.172,2.524 | |  | | 1.595 | | 1.066,2.387 | |  | 1.607 | | 1.073,2.409 | |  | |
| **Hyperactive** | 12620 | | 1630 | |  | |  | | <0.001 | |  | |  | | <0.001 |  | |  | | <0.001 | |
| 0-49mg (ref) | 7845 | | 956 | | - | | - | |  | | - | | - | |  | - | | - | |  | |
| 50-199mg | 4130 | | 573 | | 1.161 | | 1.039,1.298 | |  | | 1.177 | | 1.050,1.320 | |  | 1.187 | | 1.058,1.332 | |  | |
| 200-299mg | 474 | | 65 | | 1.145 | | 0.871,1.506 | |  | | 1.153 | | 0.867,1.532 | |  | 1.166 | | 0.878,1.550 | |  | |
| >300 mg | 171 | | 36 | | 1.922 | | 1.314,2.809 | |  | | 1.842 | | 1.240,2.735 | |  | 1.857 | | 1.250,2.758 | |  | |
| **Inattentive** | 12620 | | 1448 | |  | |  | | 0.156 | |  | |  | | 0.228 |  | |  | | 0.249 | |
| 0-49mg (ref) | 7845 | | 888 | | - | | - | |  | | - | | - | |  | - | | - | |  | |
| 50-199mg | 4132 | | 474 | | 1.015 | | 0.902,1.143 | |  | | 1.021 | | 0.904,1.154 | |  | 1.025 | | 0.906,1.159 | |  | |
| 200-299mg | 473 | | 58 | | 1.095 | | 0.824,1.454 | |  | | 1.089 | | 0.810,1.463 | |  | 1.085 | | 0.807,1.459 | |  | |
| >300 mg | 170 | | 28 | | 1.545 | | 1.029,2.320 | |  | | 1.430 | | 0.934,2.188 | |  | 1.435 | | 0.936,2.198 | |  | |
| **Paternal** | | | | | | | | | | | | | | | | | | | | | |
| **ADHD (RS-DBD)** | | 10804 | | 1462 | |  | |  | | 0.968 | |  | |  | 0.535 | |  | |  | | 0.710 |
| 0-49mg (ref) | | 2353 | | 344 | | - | | - | |  | | - | | - |  | | - | | - | |  |
| 50-199mg | | 3861 | | 478 | | 0.825 | | 0.711,0.958 | |  | | 0.854 | | 0.731,0.999 |  | | 0.842 | | 0.720,0.985 | |  |
| 200-299mg | | 3231 | | 448 | | 0.940 | | 0.808,1.094 | |  | | 0.985 | | 0.838,1.157 |  | | 0.965 | | 0.820,1.136 | |  |
| >300 mg | | 1359 | | 192 | | 0.961 | | 0.794,1.162 | |  | | 1.006 | | 0.824,1.227 |  | | 0.982 | | 0.804,1.199 | |  |
| **Hyperactive** | | 10802 | | 1432 | |  | |  | | 0.672 | |  | |  | 0.929 | |  | |  | | 0.729 |
| 0-49mg (ref) | | 2352 | | 332 | | - | | - | |  | | - | | - |  | | - | | - | |  |
| 50-199mg | | 3862 | | 489 | | 0.882 | | 0.759,1.025 | |  | | 0.910 | | 0.778,1.065 |  | | 0.892 | | 0.762,1.045 | |  |
| 200-299mg | | 3230 | | 428 | | 0.929 | | 0.796,1.084 | |  | | 0.960 | | 0.815,1.132 |  | | 0.936 | | 0.794,1.104 | |  |
| >300 mg | | 1358 | | 183 | | 0.948 | | 0.780,1.152 | |  | | 0.977 | | 0.798,1.197 |  | | 0.949 | | 0.775,1.163 | |  |
| **Inattentive** | | 10806 | | 1254 | |  | |  | | 0.888 | |  | |  | 0.492 | |  | |  | | 0.569 |
| 0-49mg (ref) | | 2351 | | 288 | | - | | - | |  | | - | | - |  | | - | | - | |  |
| 50-199mg | | 3866 | | 427 | | 0.889 | | 0.758,1.043 | |  | | 0.927 | | 0.785,1.096 |  | | 0.919 | | 0.777,1.086 | |  |
| 200-299mg | | 3230 | | 368 | | 0.921 | | 0.781,1.086 | |  | | 0.972 | | 0.816,1.158 |  | | 0.959 | | 0.804,1.143 | |  |
| >300 mg | | 1359 | | 171 | | 1.031 | | 0.842,1.262 | |  | | 1.085 | | 0.877,1.342 |  | | 1.067 | | 0.862,1.322 | |  |

Note: RS-DBD – the Disruptive Behaviour Disorders scale; N – sample size; n – number of cases; OR – odds ratio; 95% CI – 95% confidence intervals; *adjusted for child’s gender, birth year, parity, parental age, education, marital status, financial difficulties, depression and anxiety symptoms, prenatal smoking and alcohol consumption.;**additionally adjusted for partner’s caffeine consumption

### Supplementary Table S55. Associations of maternal caffeine PRS on confounders in ALSPAC

| **Confounder** | **Effect estimate** | **Effect size** | **95% CI** | **P-value** | **Sample size** |
| --- | --- | --- | --- | --- | --- |
| Maternal age | beta | 0.291 | -0.406, 0.988 | 0.412 | 7421 |
| Maternal education | beta | -0.021 | -0.211, 0.170 | 0.832 | 6860 |
| Financial difficulties | beta | 0.302 | -0.226, 0.829 | 0.263 | 6691 |
| Marital status | OR | 0.815 | 0.569, 1.169 | 0.267 | 7124 |
| Depression symptoms | OR | 0.866 | 0.540, 1.388 | 0.550 | 6706 |
| Anxiety symptoms | OR | 1.141 | 0.741, 1.756 | 0.550 | 6669 |
| Parity | beta | -0.023 | -0.153, 0.107 | 0.725 | 7040 |

Note: adjusted for principal components; OR – odds ratio; 95% CI – 95% confidence intervals.

### Supplementary Table S56. Associations of maternal caffeine PRS on confounders in MoBa

| **Confounder** | **Effect estimate** | **Effect size** | **95% CI** | **P-value** | **Sample size** |
| --- | --- | --- | --- | --- | --- |
| Maternal age | beta | -1.427 | -3.101, 0.247 | 0.095 | 14,584 |
| Maternal education | beta | -0.036 | -0.094, 0.021 | 0.213 | 13,836 |
| Financial difficulties | OR | 0.815 | 0.593, 1.120 | 0.207 | 13,484 |
| Marital status | beta | 0.014 | -0.009, 0.036 | 0.236 | 14,519 |
| Depr/Anxiety symptoms | OR | 0.798 | 0.515, 1.238 | 0.314 | 14,464 |
| Maternal ADHD | OR | 0.749 | 0.319, 1.757 | 0.506 | 8841 |
| Parity | beta | -0.082 | -0.174, 0.011 | 0.083 | 14,584 |

Note: *adjusted for birth year, genotyping batch and principal components; OR -odds ratio; 95% CI – 95% confidence intervals.

### Supplementary Table S57. Associations of maternal caffeine consumption PRS on high risk of maternal reported offspring ADHD symptoms in ALSPAC

|  | **Maternal PRS** | | | **Maternal PRS adj. for PC** | | |  |
| --- | --- | --- | --- | --- | --- | --- | --- |
| **Outcome** | **OR** | **95% CI** | **P-value** | **OR** | **95% CI** | **P-value** | **Sample size** |
| ADHD (DAWBA) | 0.859 | 0.513, 1.439 | 0.564 | 0.871 | 0.520, 1.460 | 0.601 | 5005 |
| Hyperactive | 0.928 | 0.540, 1.596 | 0.787 | 0.945 | 0.549, 1.626 | 0.837 | 5016 |
| Inattentive | 0.735 | 0.436, 1.241 | 0.250 | 0.743 | 0.440, 1.255 | 0.266 | 5013 |
| ADHD (SDQ) | 1.089 | 0.615, 1.930 | 0.769 | 1.111 | 0.627, 1.969 | 0.718 | 5103 |

Note: DAWBA – Development and Well-Being Assessment; SDQ – Strengths and Difficulties Questionnaire; OR – odds ratio; 95% CI – 95% confidence intervals; PC – principal components

### Supplementary Table S58. Associations of maternal caffeine consumption PRS on high risk of teacher reported offspring ADHD symptoms in ALSPAC

|  | **Maternal PRS** | | | **Maternal PRS adj. for PC** | | |  |
| --- | --- | --- | --- | --- | --- | --- | --- |
| **Outcome** | **OR** | **95% CI** | **P-value** | **OR** | **95% CI** | **P-value** | **Sample size** |
| ADHD (DAWBA) | 0.678 | 0.355, 1.295 | 0.239 | 0.655 | 0.342, 1.253 | 0.201 | 3486 |
| Hyperactive | 0.542 | 0.277, 1.060 | 0.074 | 0.524 | 0.268, 1.027 | 0.060 | 3485 |
| Inattentive | 0.980 | 0.492, 1.951 | 0.954 | 0.948 | 0.476, 1.890 | 0.880 | 3487 |
| ADHD (SDQ) | 0.586 | 0.287, 1.195 | 0.141 | 0.565 | 0.277, 1.155 | 0.118 | 3486 |

Note: DAWBA – Development and Well-Being Assessment; SDQ – Strengths and Difficulties Questionnaire; OR – odds ratio; 95% CI – 95% confidence intervals; PC – principal components

### Supplementary Table S59. Associations of maternal caffeine consumption PRS on high risk of maternal reported offspring ADHD symptoms in MoBa

|  | **Maternal PRS** | | | **Maternal PRS adj. for PC** | | |  |
| --- | --- | --- | --- | --- | --- | --- | --- |
| **Outcome** | **OR** | **95% CI** | **P-value** | **OR** | **95% CI** | **P-value** | **Sample size** |
| ADHD (RS-DBD) | 1.160 | 0.713, 1887 | 0.549 | 1.178 | 0.762, 1.917 | 0.510 | 7017 |
| Hyperactive | 1.397 | 0.863, 2.262 | 0.174 | 1.410 | 0.870, 2.284 | 0.163 | 7012 |
| Inattentive | 0.907 | 0.545, 1.511 | 0.709 | 0.907 | 0.545, 1.512 | 0.709 | 7017 |

Note: RS-DBD – The Disruptive Behavior Disorders Scale; OR – odds ratio; 95% CI – 95% confidence intervals; *adjusted for principal components (PC), birth year and genotyping batch

### Supplementary Table S60. Meta-analysis of PRSs of maternal smoking, alcohol and caffeine consumption in ALSPAC and MoBa

**a)**

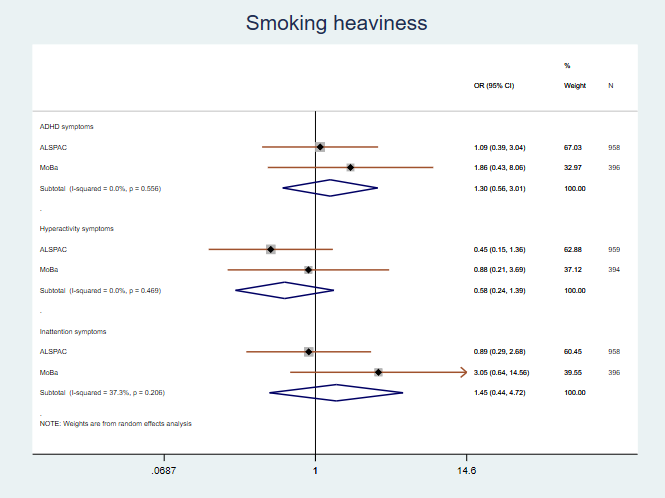

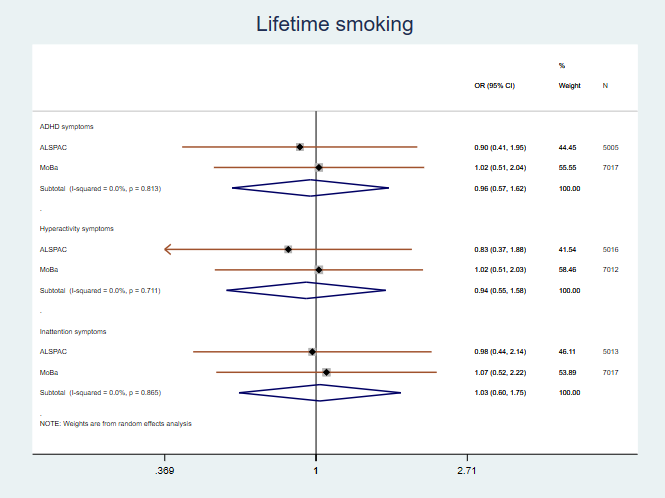

**b)**

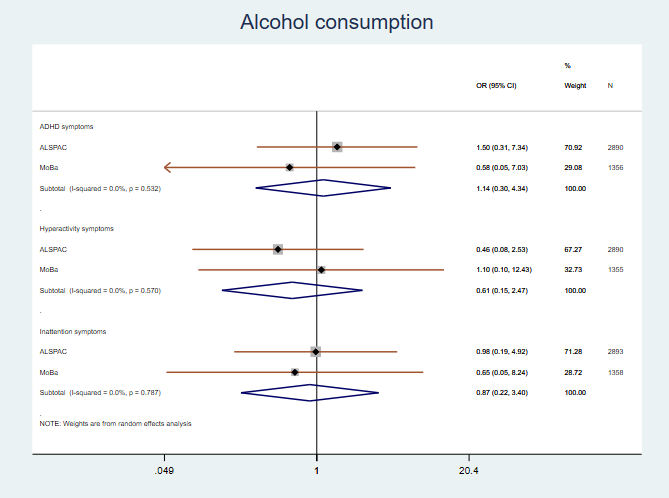

**c)**

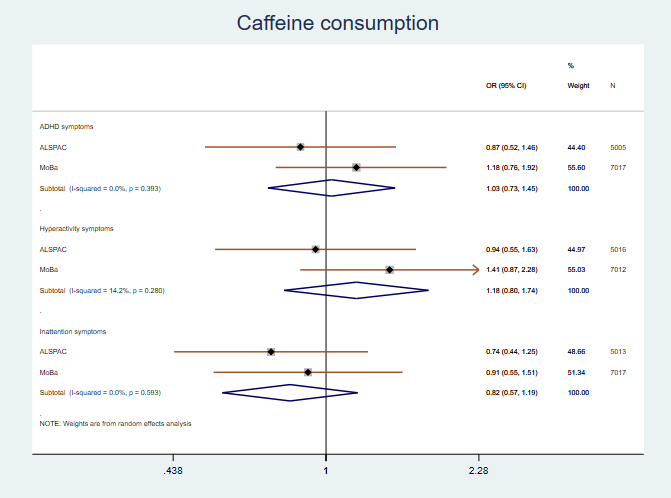

**d)**

*Note:* *Meta-analysis of PRS for smoking heaviness (a), lifetime smoking (b), alcohol consumption (c) and caffeine consumption (d).*

### Supplementary Table S61. Associations of maternal PRS for substance use on participation in ALSPAC

| **PRS** | **OR** | **95%CI** | **P-value** | **Sample size** |
| --- | --- | --- | --- | --- |
| Smoking heaviness | 0.950 | 0.561, 1.607 | 0.848 | 2345 |
| Lifetime smoking | 0.485 | 0.311, 0.757 | 0.001 | 7915 |
| Alcohol consumption | 0.978 | 0.398, 2.403 | 0.961 | 4789 |
| Caffeine consumption | 1.276 | 0.951, 1.714 | 0.105 | 7915 |

Note: OR – odds ratio; 95% CI – 95% confidence intervals; adjusted for principal components

### Supplementary Table S62. Associations of maternal PRS for substance use on participation in MoBA

| **PRS** | **OR** | **95%CI** | **P-value** | **Sample size** |
| --- | --- | --- | --- | --- |
| Smoking heaviness | 2.098 | 1.01, 4.359 | 0.047 | 1125 |
| Lifetime smoking | 0.585 | 0.427, 0.801 | 0.001 | 14,584 |
| Alcohol consumption | 0.740 | 0.237, 2.314 | 0.605 | 2861 |
| Caffeine consumption | 1.065 | 0.856, 1.324 | 0.575 | 14,584 |

Note: OR – odds ratio; 95% CI – 95% confidence intervals; adjusted for principal components, birth year and genotyping batch
